# Drug-target Mendelian randomization analysis supports lowering plasma ANGPTL3, ANGPTL4, and APOC3 levels as strategies for reducing cardiovascular disease risk

**DOI:** 10.1101/2024.01.23.24301541

**Authors:** Fredrik Landfors, Peter Henneman, Elin Chorell, Stefan K. Nilsson, Sander Kersten

**Author notes:** To whom correspondence should be addressed. **Contact information for the corresponding author:** Fredrik Landfors, Department of Public Health and Clinical Medicine, Umea University, S-901 87 Umea, Sweden.

## Abstract

**Background and Aims:** APOC3, ANGPTL3, and ANGPTL4 are circulating proteins that are actively pursued as pharmacological targets to treat dyslipidemia and reduce the risk of atherosclerotic cardiovascular disease. Here, we used human genetic data to compare the predicted therapeutic and adverse effects of APOC3, ANGPTL3, and ANGPTL4 inactivation.

**Methods:** We conducted drug-target Mendelian randomization analyses using variants in proximity to the genes associated with circulating protein levels to compare APOC3, ANGPTL3, and ANGPTL4 as drug targets. We obtained exposure and outcome data from large-scale genome-wide association studies and used generalized least squares to correct for linkage disequilibrium-related correlation. We evaluated five primary cardiometabolic endpoints and screened for potential side effects across 694 disease-related endpoints, 43 clinical laboratory tests, and 11 internal organ MRI measurements.

**Results:** Genetically lowering circulating ANGPTL4 levels reduced the odds of coronary artery disease (CAD) (odds ratio, 0.57 per s.d. protein [95%CI,0.47–0.70]) and type 2 diabetes (T2D) (odds ratio, 0.73 per s.d. protein [95%CI,0.57–0.94]). Genetically lowering circulating APOC3 levels also reduced the odds of CAD (odds ratio, 0.90 per s.d. protein [95%CI,0.82–0.99]). Genetically lowered ANGPTL3 levels via common variants were not associated with CAD. However, meta-analysis of deleterious variants revealed that *ANGPTL3* inactivation protected against CAD (odds ratio, 0.79 per allele [95%CI, 0.69–0.90]). Analysis of lowered ANGPTL3, ANGPTL4, and APOC3 levels did not identify important safety concerns.

**Conclusion:** Human genetic evidence suggests that therapies aimed at reducing circulating levels of ANGPTL3, ANGPTL4, and APOC3 reduce the risk of CAD. ANGPTL4 lowering may also reduce the risk of T2D.

**STRUCTURED GRAPHICAL ABSTRACT:** *Key Question:* Does human genetics support that triglyceride-lowering drugs targeting ANGPTL3, ANGPTL4, and APOC3 will reduce the risk of cardiometabolic disease without causing side effects?

*Key Finding:* Genetically lowered circulating ANGPTL4 reduced coronary artery disease and type 2 diabetes risk. Genetically lowered ANGPTL3 and APOC3 also reduced coronary artery disease risk, but no impact on type 2 diabetes risk was observed.

*Take-home Message:* Human genetics suggest that ANGPTL3, ANGPTL4, and APOC3-lowering medications may prevent CAD. Medicines targeting ANGPTL4 may have added benefits for patients with type 2 diabetes. Graphical abstract
summarizing the study’s methods and findings.
Graphical abstract summarizing the overall study design. The ‘Key Findings’ figure provides a summary of the results categorized into three groups. The term ‘improves’ denotes a statistically significant association with a clinically relevant effect magnitude. The term ‘weak’ refers to a statistically significant association with no clinically significant effect. ‘ASCVD’ denotes atherosclerotic cardiovascular disease. ‘T2D’ denotes type 2 diabetes.

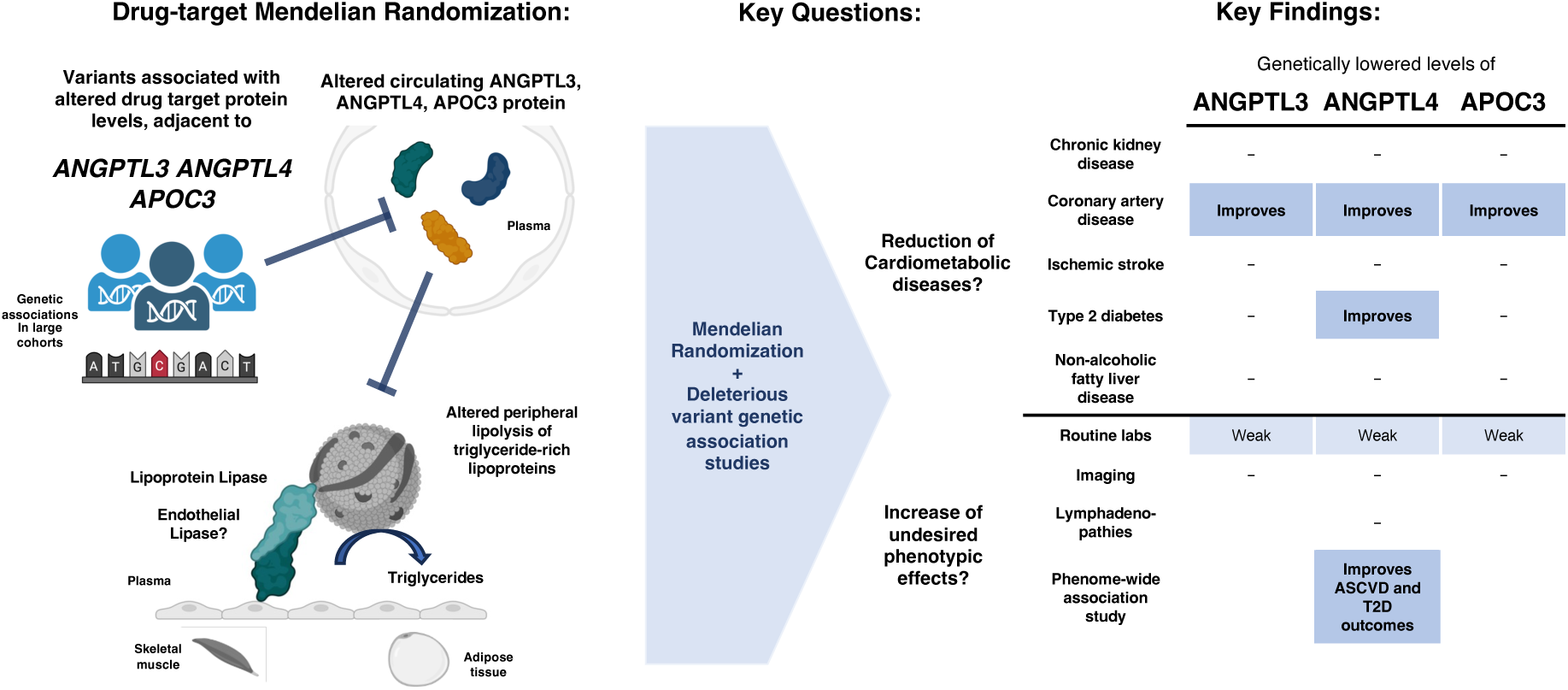

## INTRODUCTION

APOC3, ANGPTL3, and ANGPTL4 are circulating proteins that regulate plasma cholesterol and triglyceride (TG) levels. They mainly act by inhibiting the enzyme lipoprotein lipase. All three proteins are actively pursued as pharmacological targets to treat dyslipidemia and reduce the risk of atherosclerotic cardiovascular disease. The inactivation of APOC3 using ASOs (Volanesorsen, Olezarsen) has been shown to substantially reduce plasma TG levels in different patient groups with severe hypertriglyceridemia (1). Volanesorsen is a second-generation ASO that was approved in Europe for treating familial chylomicronemia syndrome. Olezarsen is a third-generation ASO that very recently received fast-track designation from the FDA. Currently, several human trials are ongoing with an RNAi against APOC3 called ARO-APOC3.

Similar to APOC3, the inactivation of ANGPTL3 using monoclonal antibodies (Evinacumab) (2–6), antisense oligonucleotides (ASOs) (Vupanorsen) (7, 8), and RNAi (ARO-ANG3) has been shown to significantly lower plasma LDL-C and TG levels in various dyslipidemic patients groups (9). Evinacumab was approved in 2021 as a treatment for homozygous familial hypercholesterolemia (HoFH), while Vupanorsen was discontinued in 2021 due to the limited reduction in non-HDL-C and TG and increases in liver fat and enzymes (10). Recent case reports suggest that Evinacumab may promote the regression of atherosclerotic plaques in HoFH patients (11, 12).

Whereas the clinical development of anti-APOC3 and -ANGPTL3 treatments have progressed well, therapies targeting ANGPTL4 have faced delay because mice deficient in ANGPTL4 develop lethal mesenteric lymphadenopathy and chylous ascites when fed a diet high in saturated fatty acids (13–15). Whether whole-body inactivation of ANGPTL4 might trigger similar pathological features in humans is unclear. As an alternative pharmacological strategy, inactivating ANGPTL4 specifically in the liver holds considerable promise (16). Despite these challenges, targeting ANGPTL4 presents a promising opportunity, as it may not only lower triglycerides and remnant cholesterol but also redirect lipids away from ectopic sites and towards adipose tissue, potentially protecting against type 2 diabetes (17).

Human genetic data can be leveraged to predict the clinical effect of the pharmacological inactivation of genes or proteins. Here, we aimed to compare the predicted therapeutic effects of APOC3, ANGPTL3, and ANGPTL4 inactivation by investigating the biological and clinical impact of inactivation variants in the respective genes. In addition, to address safety concerns, we compared the predicted detrimental effects of APOC3, ANGPTL3, and ANGPTL4 inactivation on relevant disease outcomes. We conclude that therapies specifically aimed at decreasing plasma ANGPTL3, ANGPTL4, and APOC3 levels are expected to reduce the risk of coronary artery disease without raising safety concerns. Therapies targeting ANGPTL4 levels are expected to favorably impact the risk of type 2 diabetes. This suggests that reducing ANGPTL4 could offer therapeutic advantages to a wider group of patients with dyslipidemia and type 2 diabetes.

## METHODS

### Study design

The study was performed in four sequential steps as summarized in **Figure 1**. First, a two-sample Mendelian randomization (MR) study was conducted to measure the association between ANGPTL3, ANGPTL4, and APOC3 lowering with cardiometabolic diseases and risk factors. Second, two-sample MR was conducted to measure the target proteins’ association with phenotypes related to potential adverse effects. Third, validation analyses were conducted to further assess the plausibility of the findings obtained from steps 1-2. Lastly, to measure the association between profound genetic inactivation of the target proteins and CAD, deleterious variant analyses in the UK Biobank were performed and meta-analysed with previous studies.

**Figure 1.**
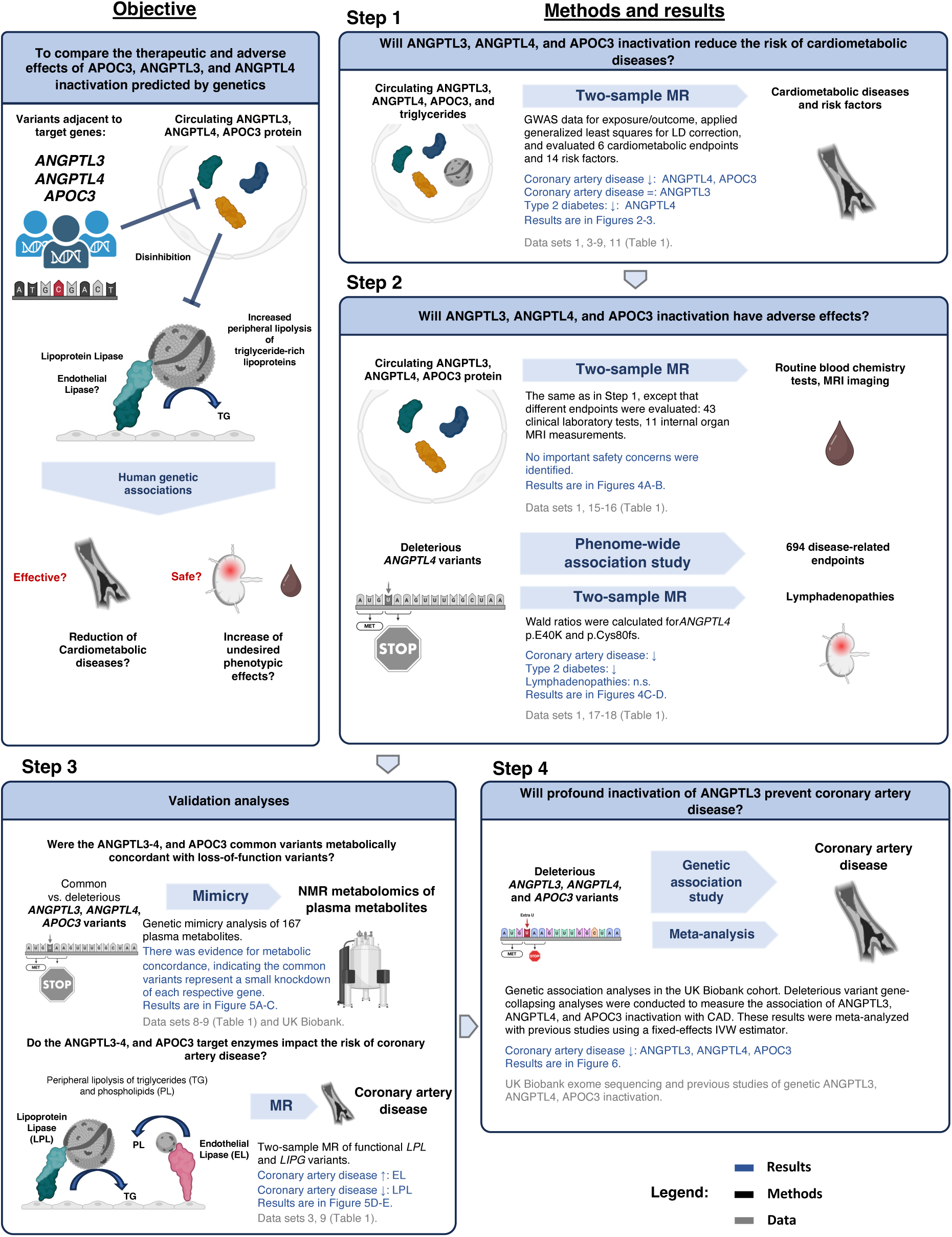
Study design flow chart summarizing the objective, methods, and results. ‘LD’ indicates linkage disequilibrium. ’n.s.’ denotes not significant. ‘LPL’ indicates lipoprotein lipase. ‘MR’ indicates Mendelian Randomization. ‘EL’ indicates endothelial lipase. ‘CAD‘ indicates coronary artery disease. ‘IVW’ indicates inverse-variance weighted.

### Steps 1-2

#### Genetic instruments

To estimate the causal effects of pharmacologically inactivating the *ANGPTL3*, *ANGPTL4*, and *APOC3* genes, we performed two-sample drug-target MR. We used, as instrumental variables (IV), genetic variants within 2.5 kilobase pairs (Kb) of the target gene that had genome-wide significant associations (P-value ≤ 5×10^-8^) with protein abundance (called *cis* protein quantitative trait loci, *cis-* pQTLs) or plasma TG, as determined by genome-wide association studies (GWAS). Variants adjacent to the target genes were clumped at an LD threshold of r^2^ ≥ 0.10 to avoid GLS-related multicollinearity issues. Residual LD was accounted for using the generalized least squares (GLS) IVW estimator described below.

#### Drug-target MR

The precision of the inverse-variance weighted (IVW) estimator can be influenced by LD-related correlation between the genetic IV in the drug target genes *cis’* position. Therefore, we used a GLS IVW MR estimator to correct for this potential source of bias (18, 19). The GLS-corrected MR approach can be conceptualized as combining the independent information of variants near a target gene while maintaining robust standard errors through weighting for their LD-related correlation. Further information regarding Drug-target MR methodology, GLS, LD matrix sensitivity, and sample overlap bias are found in the **Supplemental Methods**.

Due to the complex structure of the *APOA1*-*APOA5*-*APOC3* locus, we supplemented the original analyses with a second model of APOC3 lowering. In this model, APOC3 lowering was instrumented through the *APOC3 c.55+1G>A* splice donor loss variant solely, as this variant is a high-confidence predicted loss-of-function variant (gnomAD Genome Aggregation Database v.4.0.0, https://gnomad.broadinstitute.org) independent of other common variants in this genomic region (20). The *APOC3 c.55+1G>A* MRs used a Wald ratio estimator. Furthermore, we used *LPL*-adjacent and genome-wide TG-associated variants as positive controls. LPL was analyzed using drug-target MR. For the genome-wide TG-associated variant MR, we tested the causal effect of TG using variants in chromosomes 1-22 associated with TG at P-value ≤ 5×10^-8^. An LD clumping window of 500 Kb and a threshold of r^2^ ≥ 0.001 was applied before analysis using an IVW estimator.

#### Data sources

Plasma protein abundance was measured in GWAS using the SomaScan and Olink platforms (21, 22). GWAS data on plasma TG, LDL cholesterol, HDL cholesterol, apolipoprotein B, apolipoprotein A1, and lipoprotein(a) were retrieved from the 2018 Neale Lab UK biobank analysis (http://www.nealelab.is/uk-biobank/). For the functional variant analyses, genetic association data on TG, LDL cholesterol, and HDL cholesterol were retrieved from the AstraZeneca UK biobank exome sequencing-based phenome-wide association study (PheWAS) portal (23). We obtained outcome summary data from GWAS of six cardiometabolic disease endpoints, 16 cardiometabolic risk markers, 43 routine clinical chemistry tests, 11 internal organ MRI measurements, and five abdominal lymphadenopathy-related phenotypes (see **Table 1**, **Supplemental Table 1,** and **Supplemental Methods**). Phenome-wide MR analyses were conducted in FinnGen and the UK biobank. FinnGen integrates genotype data from Finnish biobanks with longitudinal health registry data (24). The UK Biobank is a large-scale research resource containing genetic, blood chemistry, imaging, and health record data from half a million UK participants (25). The FinnGen data freeze 10 and UK biobank meta-analysis (https://public-metaresults-fg-ukbb.finngen.fi) stores genetic association statistics on 694 disease-related outcomes from 301,552–882,347 individuals. Further details on the selection of GWAS and the definition of exposures and outcomes are given in the **Supplemental Methods**.

**Table 1.**
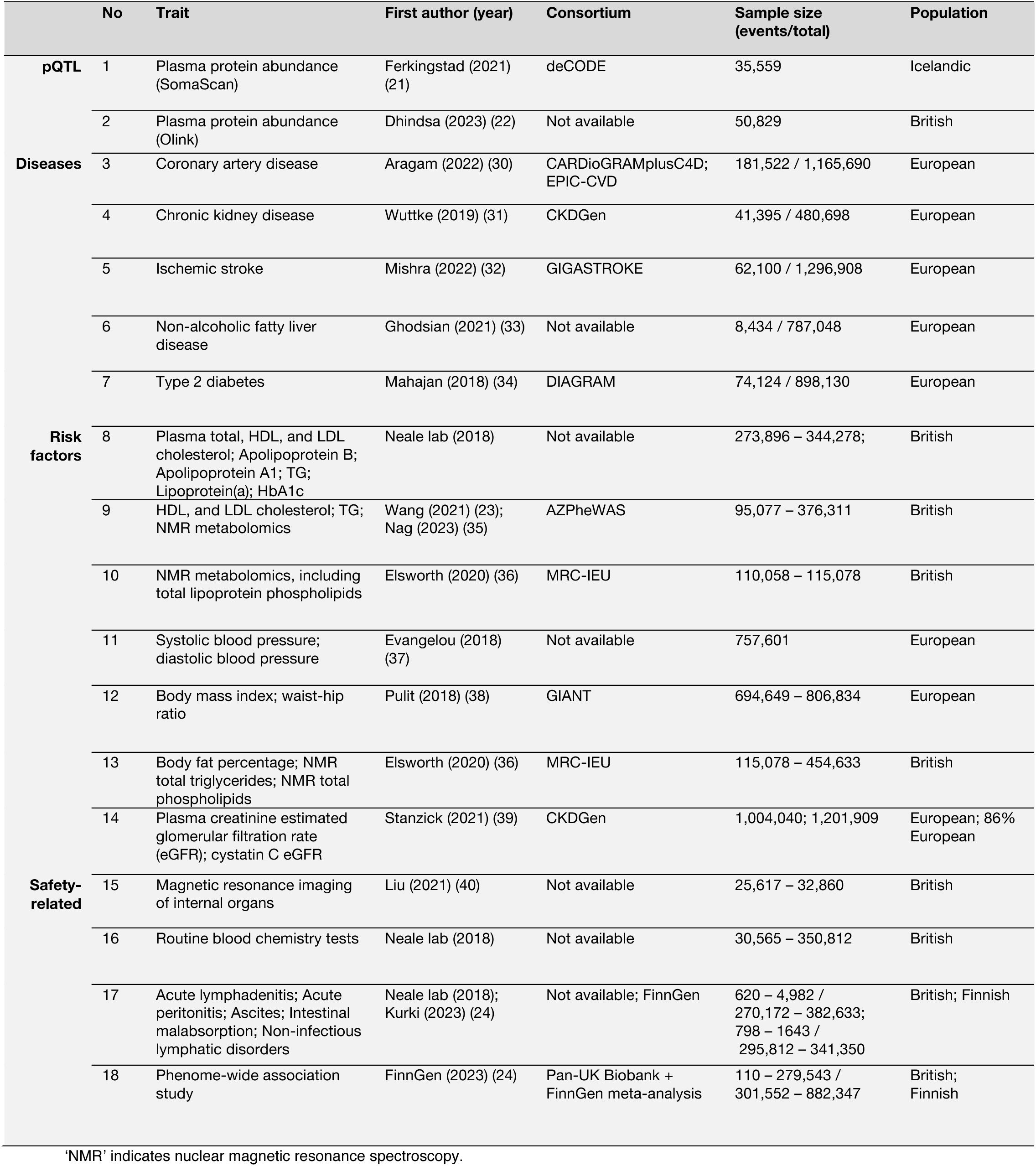
Description of GWAS data sets.

#### Colocalization analyses

Drug-target MR substantially relies on the assumption that LD (a phenomenon in which neighboring genetic variants are inherited together more frequently than anticipated by chance (26)) does not confound the association between variant and outcome. In cases where there are distinct genetic variants affecting both the exposure and the outcome, and they are connected through LD, there is a risk of making incorrect conclusions (27). To limit this issue, we performed colocalization analyses, which test whether two independent association signals in the same gene region are consistent with having a single shared causal variant (that is, testing if the association signals are ‘colocalized’) (28). To assess possible confounding from LD, all drug target MR analyses were complemented by colocalization analysis of the 500 Kb (±250 Kb) region surrounding each target gene (28). Further details regarding the colocalization analyses are provided in the **Supplemental Methods**.

#### Lymphadenopathy and phenome-wide MR analyses

A Wald ratio estimator was used for the single-variant MR of lymphadenopathy-related phenotypes and the phenome-wide MR conducted in FinnGen (29). The variants were selected based on being within 2.5 Kb of the drug target gene, their strength of association with target protein plasma abundance (P ≤ 5×10^-8^), their strength of association with triglyceride levels (P ≤ 5×10^-8^), availability, and their functional consequence. For further details regarding genetic instrument justification for the phenome-wide MRs, see the **Supplemental Methods**.

### Step 3

#### Genetic mimicry analyses

Genetic mimicry analysis was used to compare the metabolic concordance between common and deleterious variants adjacent to the *ANGPTL3*, *ANGPTL4*, and *APOC3* genes. This method uses linear regression to determine the extent of similarity between different variants’ genetic associations in high-dimensional data sets (41, 42). The degree of concordance was reported as the coefficient of determination (R^2^). Genetic associations between the common variants and 167 plasma metabolites were measured by drug-target MR with plasma TGs as the exposure using data sets 8 and 10 (see **Table 1**). Deleterious variants were defined as any protein-truncating variant with an allele frequency <0.05 or missense variants with a REVEL pathogenicity prediction score >0.25 and allele frequency <0.00025 (see **Supplemental methods**). The effects of deleterious variants were determined by regressing plasma concentration of metabolites on deleterious variant carrier status in 181,672 UK Biobank participants (see **Supplemental Methods** for details).

#### Robustness checks and sensitivity analyses

We performed sensitivity MR analyses of *ANGPTL3*, *ANGPTL4*, *APOC3*, *LPL*, and *LIPG* on CAD by restricting the genetic instrument selection to variants within these target genes predicted to have functional impacts. This strategy aimed to mitigate potential biases arising from common non-coding small-effect variants outside the target genes, which could be confounded due to linkage disequilibrium with other genes in the same genomic region. Ensembl Variant Effect Predictor (VEP) version 109 (43) was used to annotate variants within 2.5 Kb of the target gene associated (P ≤ 0.01) with target protein levels and plasma triglycerides. Non-coding variants outside of the 5’ untranslated region (UTR), 3’ UTR, or splice site regions were filtered out and excluded from further analysis, as were missense variants lacking SIFT deleterious or PolyPhen likely or probably damaging annotations. MR was conducted for single variants using the Wald ratio estimator, and meta-analysis was performed using a random-effects IVW estimator.

### Step 4

#### Meta-analysis of the impact of deleterious variants on CAD

We conducted genetic association analyses in the UK Biobank (see **Supplemental Methods**) and meta-analyzed the results with previous studies to assess how deleterious variants in *ANGPTL3*, *ANGPTL4*, and *APOC3* impact CAD risk. To minimize the influence of incorrect genotype calls for rare variants, the meta-analysis was limited to studies where genotypes were determined by DNA sequencing. When multiple papers reported on individuals from overlapping cohorts or case-control studies, we selected the substudy with the largest sample size for inclusion in the meta-analysis. The meta-analyses were restricted to European ancestries. We determined the impact of the inactivating variants on CAD risk per mmol/L reduction of TG and per inactivating allele using fixed-effect IVW estimators. If no within-sample association of inactivating variants with TG concentrations (in mmol/L) was available, the combined IVW meta-analysis TG estimate was used as the denominator to determine the CAD odds per mmol/L TG effect. Statistical heterogeneity across studies was estimated by calculating the Cochran Q statistic.

### Statistics

#### Multiple testing

P-values and 95% confidence intervals (CI) are reported using analysis-type Bonferroni multiple comparisons correction. In the primary MR analyses, we corrected for the five cardiometabolic disease outcomes that were run across three different drug-target gene exposures (*ANGPTL3, ANGPTL4, APOC3*) for protein abundance, and four genes for the TG exposure (*ANGPTL3, ANGPTL4, APOC3, LPL*). Additionally, we included five genome-wide triglyceride MR models, totaling 40 comparisons for the cardiometabolic disease outcomes. In the cardiometabolic risk factor MR analyses of *cis*-pQTLs, we made corrections for 45 multiple comparisons (15×3). Similarly, imaging and blood chemistry MR analyses were corrected for 33 (11×3), and 129 (43×3) multiple comparisons, respectively. We did not perform multiple comparison corrections for the *ANGPTL4-*targeted MR analyses of the lymphadenopathy-related phenotypes. This was because identifying potential safety concerns that needed to be addressed was considered more critical than stringent multiplicity correction for these specific outcomes. Similarly, the primary motivation for performing the functional variant-limited CAD MR analyses and deleterious variant meta-analysis was to reduce the risk of false-negative findings. Additionally, we wanted to ensure that these CIs and P-values remained comparable across different studies. These CIs and P-values were, therefore, not corrected for multiple comparisons. The significance threshold in the phenome-wide *cis*-pQTL MR analyses was set at 2082 multiple comparisons (694 phenotypes in the FinnGen R10 and UK biobank meta-analysis, times three genes).

## RESULTS

The results of the drug-target MR analyses of cardiometabolic diseases, cardiometabolic risk factors, and the safety-related endpoints are presented in **Figure 2**, **Figure 3**, and **Figure 4**, respectively. MR scatter, colocalization plots, and results tables with greater detail are provided in **Supplemental Figures 1-2** and **Supplemental Table 2**. Detailed PheWAS results are provided in **Supplemental Tables 6-9**. The genetic variants selected for inclusion as instrumental variables in one or more of the MR analyses are shown in **Table 2**.

**Figure 2.**
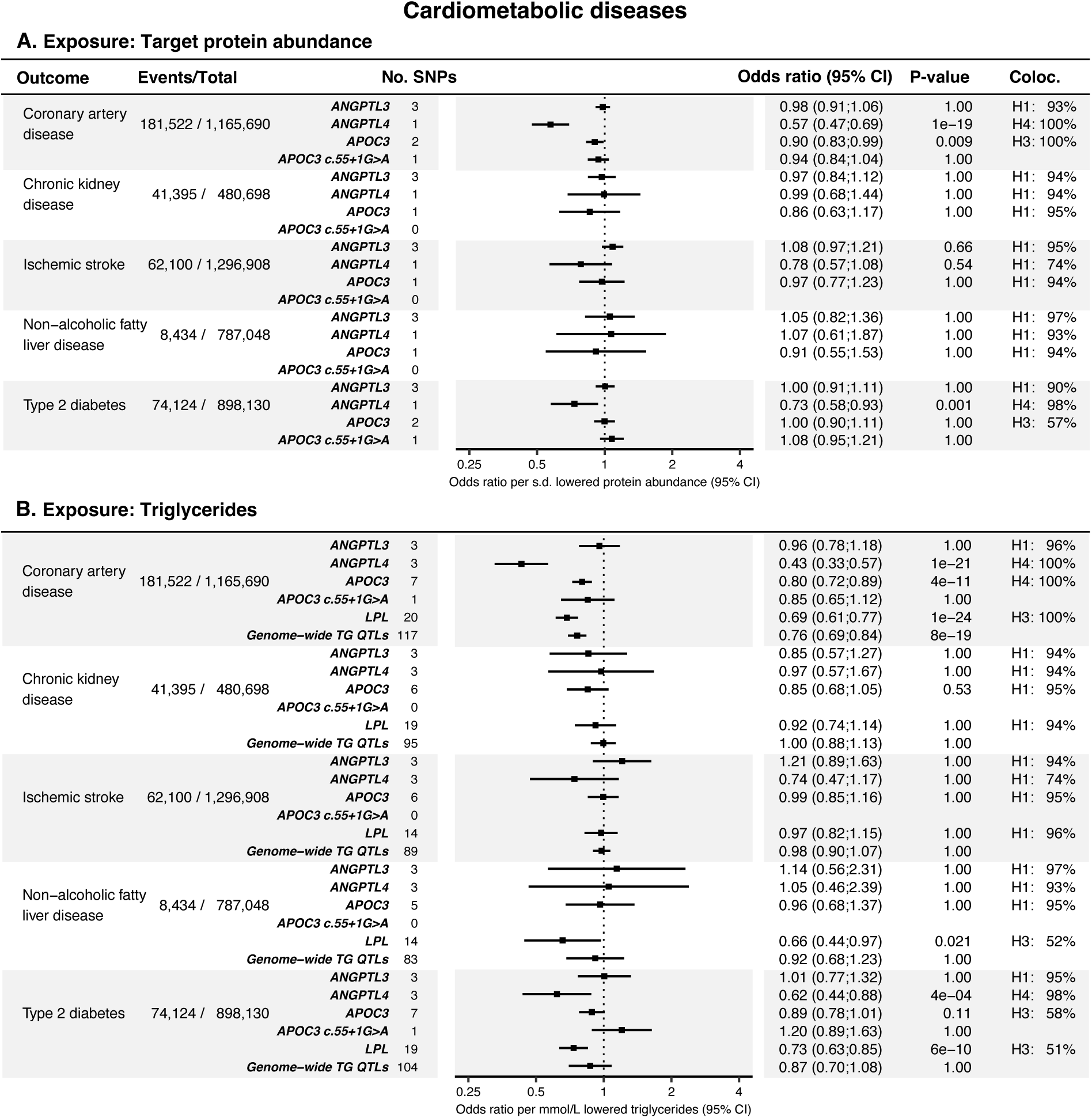
Results of MR analyses of cardiometabolic disease outcomes. **A:** Forest plot and table of the *cis-*pQTL-based MR analyses. ‘Events/total’ indicates the outcome study’s case count and total sample size. ‘No. SNPs’ specifies the number of variants included in the MR model. Zero SNPs indicate that none of the genetic instruments were detected in the outcome data set. ‘Coloc.’ shows the colocalization hypothesis (H0-4) with the highest posterior probability (see the ’Methods’ section for details about their interpretation). **B:** Results of the MR analysis using TG levels as the exposure. The ‘Genome-wide TG QTLs’ and ‘LPL’ models were positive controls. ‘Genome-wide TG QTLs’ indicate the MR model that included independent (r^2^ < 0.001, 500Kb clumping window) variants associated with TG levels (P≤5×10^-8^) across chromosomes 1-22. ‘LPL’ denotes lipoprotein lipase.

**Figure 3.**
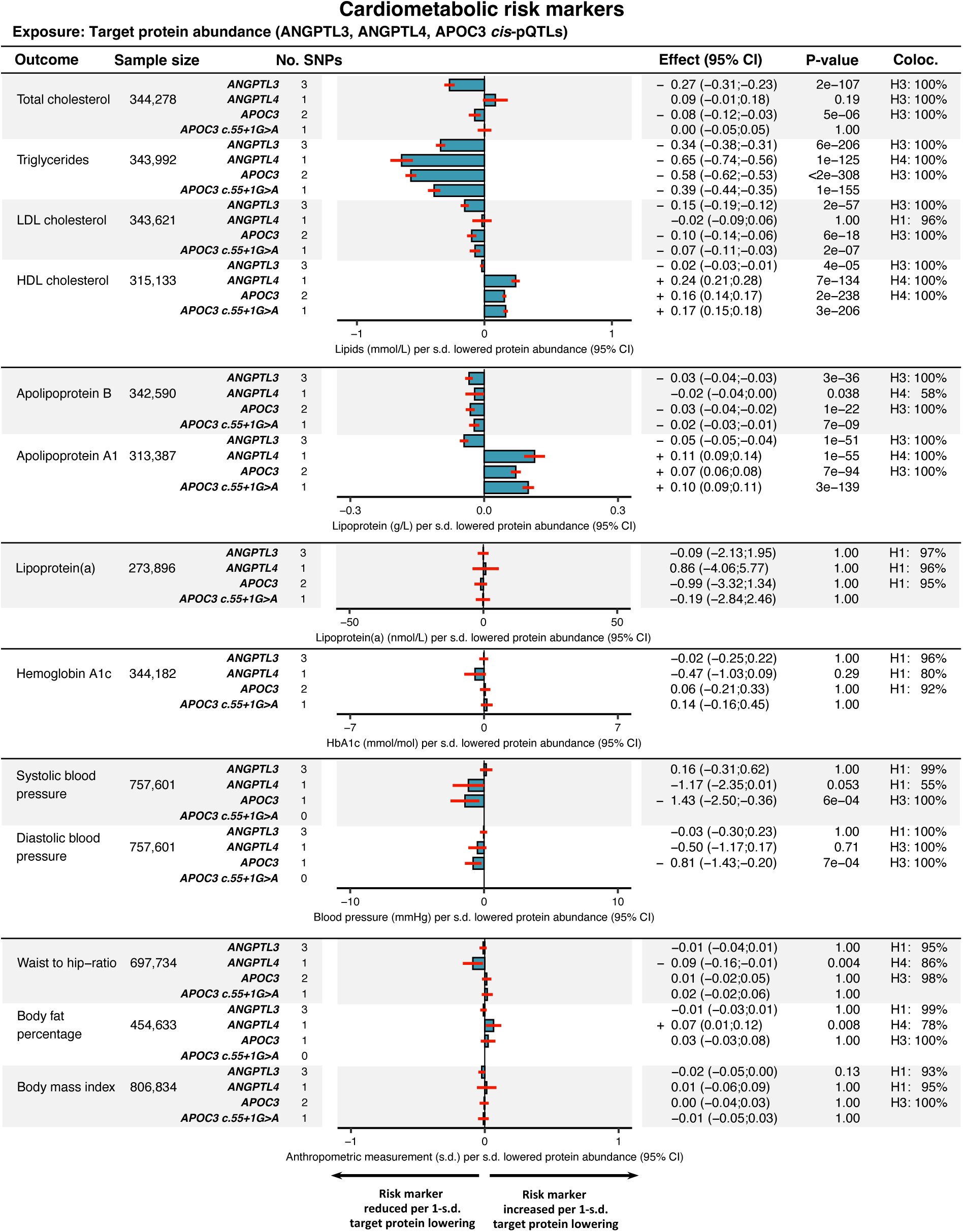
Results of MR analyses of cardiometabolic disease risk factors. The results are presented as bar plots, showing the magnitude of the effect per s.d. lowered protein abundance. The red lines indicate the 95 % CI. The results from cis-pQTL MR of the estimated glomerular filtration rate (eGFR) by Cystatin C and plasma Creatinine, respectively, are given in **Supplemental Figure 3**.

**Figure 4.**
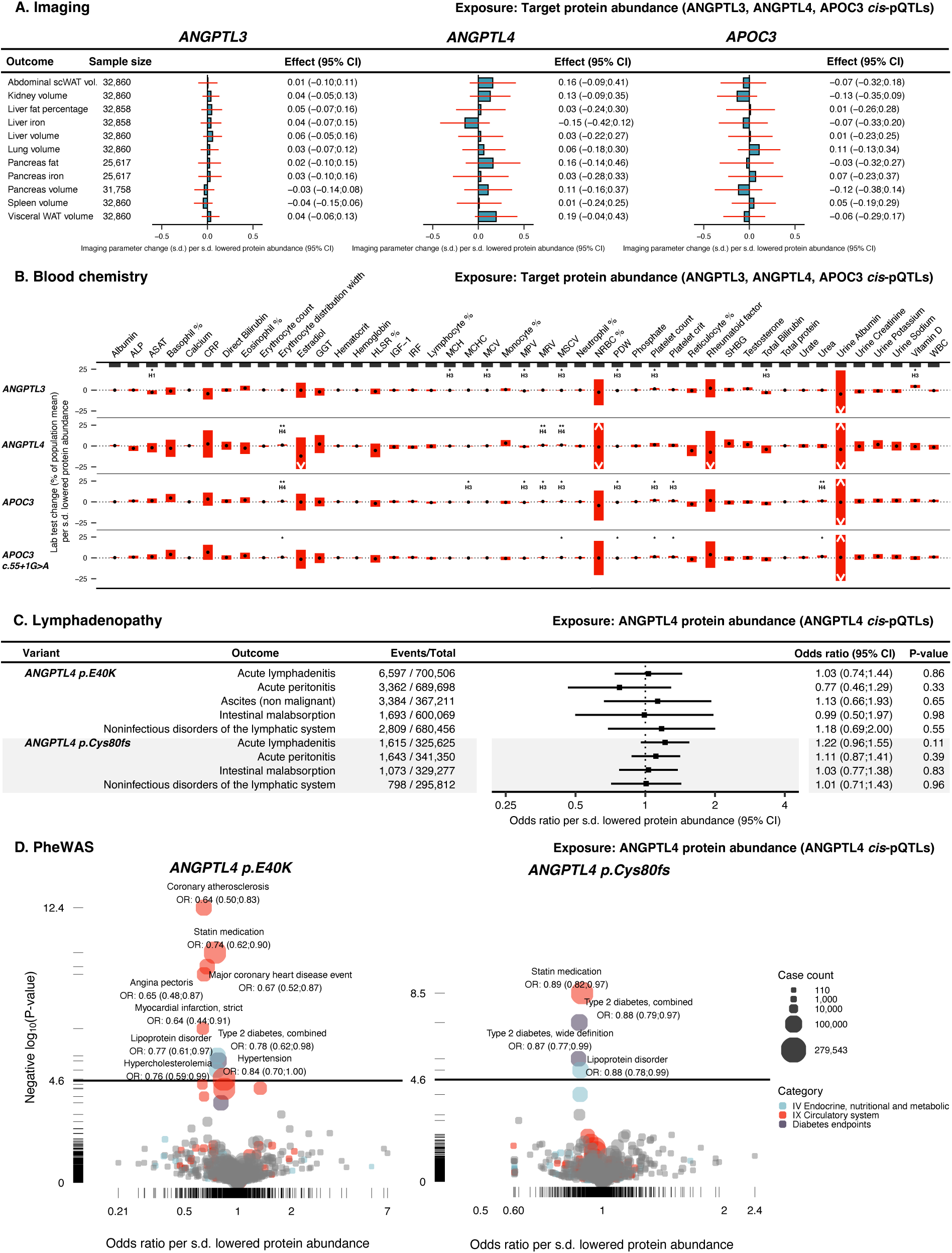
Results of MR analysis of potential adverse effects. **A:** *Cis*-pQTL MR results on the imaging outcomes. Bar plots and red lines indicate the effect and 95% CI:s. ‘scWAT’ indicates subcutaneous white adipose tissue. ‘vol.’ indicates volume. **B:** *Cis*-pQTL MR of the clinical laboratory outcomes. The red bars indicate the 95 % CI. The black dots indicate the effect point estimate. * indicate P < 0.05. ** indicates P < 0.05 with a shared causal variant (H4). A list explaining the abbreviations is provided in the supplemental material (**Table S1**). **Supplemental Figure 5** shows the results on a 1-s.d. scale. **C:** Results of ANGPTL4 *cis*-pQTL MR of mesenteric lymphadenopathy and malabsorption-related phenotypes. **D:** Volcano plot displaying the results of ANGPTL4 *cis*-pQTL phenome-wide MR scans on 694 outcomes in the FinnGen and UK Biobank meta-analysis (see **Table 2** and **Supplemental Table 1** for the reference and link to data, respectively). The *y*-axis solid straight lines indicate the phenome-wide significance threshold.

**Table 2.** Genetic instruments.

Effect (P-value)
|  | rsID | Variant | HGVS | Consequence | Frequency | Protein<br>(SomaScan) | Protein<br>(Olink) | Protein<br>(ELISA)<sup>‡</sup> | TG |
| --- | --- | --- | --- | --- | --- | --- | --- | --- | --- |
| **ANGPTL3** | rs79151558 | 1:62595131:A>G |  | Upstream variant | 0.028 | -0.32<br>(8.1e-39) | -0.27<br>(1.4e-36) | - | -0.06<br>(5.4e-13) |
|  | rs11207997* | 1:62596235:C>T |  | Upstream variant | 0.338 | -0.22<br>(1.2e-144)* | -0.31<br>(<2e-308)* | - | -0.08<br>(2.7e-221)* |
|  | rs17123728 | 1:62602628:G>A | c.931+248G>A | Intron variant | 0.04 | n.s. | - | - | 0.03<br>(2.5e-08) |
|  | rs35285100 | 1:62603486:T>C | c.932-483T>C | Intron variant | 0.071 | 0.10<br>(2.3e-09) | 0.08<br>(6.3e-10) | - | n.s. |
|  | rs10789117* | 1:62606594:A>C |  | Downstream variant | 0.354 | -0.22<br>(1.7e-144)* | -0.31<br>(<2e-308)* | - | -0.08<br>(1.6e-221)* |
|  | rs6678483* | 1:62608771:C>A |  | Downstream variant | 0.354 | -0.22<br>(1.7e-144)* | -0.31<br>(<2e-308)* | - | -0.08<br>(2e-221)* |
| **ANGPTL4** | rs116843064<sup>‡</sup> | 19:8364439:G>A | p.Glu40Lys<sup>‡</sup> | Missense variant<sup>‡</sup> | 0.024 | -0.32<br>(6.6e-35) | -0.35<br>(6.8e-48) | -0.45<br>(4.8e-05)<sup>‡</sup> | -0.21<br>(3.2e-127) |
|  | rs149480839 | 19:8368237:C>T | c.548-982C>T | Intron variant | 0.03 | n.s. | - | - | -0.05<br>(2.6e-14) |
|  | rs139469000 | 19:8376253:C>T |  | Downstream variant | 0.28 | n.s. | - | - | -0.02<br>(1.7e-09) |
| **APOC3** | rs2727788 | 11:116828247:C>A |  | Upstream variant | 0.262 | n.s. | - | - | -0.05<br>(7.3e-83) |
|  | rs138326449 | 11:116830638:G>A | c.55+1G>A | Splice donor variant | 0.002 | -2.19<br>(3.2e-142) | - | - | -0.86<br>(3.4e-157) |
|  | rs5141<sup>†</sup> | 11:116831407:T>C | c.179+511T>C | Intron variant | 0.916 | -0.17<br>(2.1e-38)<sup>†</sup> | - | - | -0.19<br>(<2e-308)<sup>†</sup> |
|  | rs5132 | 11:116832062:C>T | c.180-702C>A | Intron variant | 0.015 | n.s. | - | - | 0.06<br>(3.1e-08) |
|  | rs12721031 | 11:116833789:C>T |  | Downstream variant | 0.023 | n.s. | - | - | -0.06<br>(2.6e-12) |
|  | rs10750098<sup>†</sup> | 11:116834852:G>T |  | Downstream variant | 0.115 | -0.17<br>(2.1e-38)<sup>†</sup> | - | - | -0.19<br>(<2e-308)<sup>†</sup> |
|  | rs12721028 | 11:116834874:A>G |  | Downstream variant | 0.159 | n.s. | - | - | 0.04<br>(2e-35) |
|  | rs12718462 | 11:116835003:T>C |  | Downstream variant | 0.066 | n.s. | - | - | -0.05<br>(1.8e-22) |

\*† These variants were in strong linkage disequilibrium (LD) and, therefore, showed the same associations with TG levels and plasma ANGPTL3/APOC3 abundance. Because they were in strong LD, they were never included in the same MR model (see **Methods** and **Supplemental Table 2**).
<sup>‡</sup> The association between *ANGPTL4* p.E40K coding variant carrier status and plasma ANGPTL4 protein was confirmed in a separate study by ELISA using antibodies that were shown by Western blotting to similarly detect wildtype ANGPTL4 and ANGPTL4 containing the E40K substitution, indicating that the association was not attributable to epitope-binding artifacts (see **Supplemental Methods**).

### Step 1

#### Drug-target MR of cardiometabolic diseases

Genetically mediated changes in plasma ANGPTL3 protein abundance were not associated with a reduced risk of any cardiometabolic outcome (**Figure 2A**), nor were *ANGPTL3*-mediated changes in plasma TG (**Figure 2B**).

The p.E40K coding variant was the only variant that qualified as a *cis-*pQTL in the *ANGPTL4* region. *ANGPTL4* p.E40K is a common missense variant (allele freq. ∼2% in Europeans) that destabilizes ANGPTL4 after secretion and prevents ANGPTL4 from inhibiting LPL (44). The association between the *ANGPTL4* p.E40K coding variant and plasma ANGPTL4 protein was validated via ELISA in a separate cohort. The association was -0.45 s.d. protein per allele, P=4.8×10^-5^, comparable to the associations detected with the Olink and Somascan platforms (see **Table 2**). The ELISA antibodies detected wild-type and E40K ANGPTL4 proteins to a comparable degree, as determined by Western blot analysis (see **Supplemental Methods** for details). This suggests that the observed association was not attributable to epitope-binding artifacts.

Changes in ANGPTL4 protein levels via *ANGPTL4* p.E40K were associated with a decreased risk of CAD (OR: 0.57, P=1×10^-19^), and T2D (OR: 0.73, P=0.001) (**Figure 2A**). Similarly, changes in plasma TG levels via three *ANGPTL4*-adjacent variants were associated with a decreased risk of CAD (OR: 0.43, P=1×10^-21^), and T2D (OR: 0.62, P=4×10^-4^) (**Figure 2B**). In addition, colocalization analyses indicated a high probability of *ANGPTL4* p.E40K being a shared causal variant for ANGPTL4 levels and TG levels with CAD and T2D (pp.H_4_: 98–100%) (**Figures 2A-B**).

Changes in APOC3 levels caused by *APOC3*-adjacent variants were associated with a reduced risk of CAD (OR: 0.90, P=0.009) (**Figure 2A**), as were changes in TG levels through *APOC3*-adjacent variants (OR: 0.80, P=4×10^-11^). The *APOC3* c.55+1G>A splice donor loss variant had a substantial impact on plasma APOC3 levels (-2.19 s.d. protein, P=3.2×10^-142^) and plasma TG (-0.86 mmol/L, P=3.4×10^-157^) (**Table 2**). When compared to the model allowing for multiple variants in the *APOC3* region, APOC3 lowering modeled through the *APOC3* c.55+1G>A variant demonstrated a comparable correlation with CAD in terms of the direction of its effect. However, the association was non-significant (**Figure 2**).

Similar to APOC3 and ANGPTL4, changes in plasma TG levels through *LPL*-adjacent variants were associated with a reduced risk of CAD (OR: 0.69, P=1×10^-24^), NAFLD (OR: 0.66, P=0.021), and T2D (OR: 0.73, P=6×10^-10^) (**Figure 2B**).

#### Drug-target *cis*-pQTL MR of cardiometabolic risk factors

Genetically lowered plasma ANGPTL3 levels were associated with reduced total cholesterol (-0.27 mmol/L, P=2×10^-107^), TG (-0.34 mmol/L, P=6×10^-206^), LDL-C (-0.15 mmol/L, P=2×10^-57^), ApoB (-0.03 g/, P=3×10^-36^), and ApoA-I levels (-0.05 g/L, P=1×10^-51^), while the effect on HDL-C was comparatively weak (-0.02 mmol/L, P=4×10^-5^) (**Figure 3**).

Genetically lowered plasma ANGPTL4 levels instrumented through the p.E40K variant were associated with reduced plasma TG (-0.65 mmol/L, P=1×10^-125^) and weakly reduced ApoB levels (-0.02 g/L, P=0.038), as well as increased ApoA1 (0.11 g/L, P=1×10^-55^) and HDL-C levels (0.24 mmol/L, P=7×10^-134^) (**Figure 3**). Genetically lowered plasma ANGPTL4 levels were also associated with modest reductions in the waist-hip ratio (-0.09 s.d., P=0.004), and a small increase in body fat percentage (0.07 s.d., P=0.008) (**Figure 3**).

Genetically lowered plasma APOC3 levels were associated with reduced TG levels (-0.58 mmol/L, P < 2×10^-308^) (**Figure 3**). APOC3 levels were also associated with ApoB (-0.03 g/L, P=1×10^-22^), LDL-C (- 0.10 mmol/L, P=6×10^-18^), HDL-C (0.16 mmol/L, P = 2×10^-238^), and total cholesterol (-0.08 mmol/L, P=5×10^-6^) (**Figure 3**). In terms of association and effect directionality, these results closely resembled those of the *APOC3 c.55+1G>A* model (**Figure 3**).

### Step 2

#### Drug-target MR of potential adverse effects

Genetic lowering of plasma protein levels of the target genes was not associated with any of the MRI imaging endpoints (**Figure 4A**). 9, 3, 9, and 6 out of the 43 routine clinical laboratory tests showed statistically significant associations by drug-target cis-pQTL MR of the ANGPTL3, ANGPTL4, APOC3, and c.55+1G>A models, respectively (**Figure 4B**). The effect magnitudes were weak. For example, genetically lowered ANGPTL3 and APOC3 levels were significantly associated with increased platelet count. However, the effect was estimated to be 4–5×10^9^ cells/L (equalling 0.06-0.08 s.d.) per s.d. lowered plasma protein levels, which was minimal compared to the population mean value of 252×10^9^ cells/L.

Given that safety concerns have arisen from preclinical models of ANGPTL4 deficiency, we conducted targeted cis-pQTL MR analyses of ANGPTL4 on disease phenotypes that may be associated with abdominal lymphadenopathy. The mechanism behind the fatal chylous lymphadenopathy observed in mice was purportedly the loss of inhibition of LPL in macrophages, which caused them to take up excess lipids, leading to massive inflammation in the mesenteric lymph system (14). Exposure to ANGPTL4 inactivation was instrumented using two different models: by the *ANGPTL4* p.E40K coding variant, and by the *ANGPTL4* p.Cys80frameshift (fs) variant. *ANGPTL4* p.Cys80fs is a high-confidence predicted loss-of-function variant (gnomAD v.4.0.0). It is enriched in Finns compared to non-Finnish Europeans (allele frequency: 0.63% vs. 0.05%). *Cis*-pQTL MR via the relatively common *ANGPTL4* p.E40K variant was conducted at five different phenotypes that may be related to lymphadenopathy and malabsorptive states. Four had overlapping phenotype codes between the UK biobank and FinnGen and were meta-analyzed using IVW meta-analysis. ANGPTL4 levels via p.E40K were not associated with any of the five phenotypes (**Figure 4C**). However, since the confidence intervals were wide, we cannot fully exclude an association of p.E40K within this interval. Genetically lowered plasma ANGPTL4 levels via the *ANGPTL4* p.Cys80fs variant were not associated with any of the four FinnGen phenotypes that may be related to lymphadenopathy and malabsorptive states (**Figure 4C**).

To investigate if there was any genetic evidence for unknown ANGPTL4-mediated side effects, we performed cis-pQTL MR on 694 disease-related phenotypes in FinnGen and the UK Biobank via the *ANGPTL4* p.E40K and p.Cys80fs variants. Using a phenome-wide significance threshold of 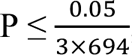, we found no evidence for increased risk of any endpoint via p.E40K- or p.Cys80fs-lowered ANGPTL4 levels (**Figure 4D**). Instead, we found phenome-wide evidence that p.E40K reduced the risk of four CAD-related phenotypes, including myocardial infarction and one T2D-related phenotype, while also being associated with a lowered probability of statin prescription, lipoprotein disorders, and hypercholesterolemia (**Figure 4D**). Additionally, *ANGPTL4* p.Cys80fs was associated with a decreased risk of two T2D-related outcomes and a lowered probability of statin prescription and hypercholesterolemia diagnosis (**Figure 4D**).

The phenome-wide MR results of lowered plasma ANGPTL4 levels were compared with ANGPTL3 and APOC3 by cis-pQTL MR of the 694 FinnGen and UK Biobank endpoints using the *ANGPTL3* c.*52_*60del and *APOC3* c.55+1G>A. Genetically lowered plasma ANGPTL3 levels were associated with a reduced risk of being prescribed statin medication, two lipid-related diagnosis codes but not any other patient-related outcome (**Supplemental Figure 4A**). *APOC3 c.55+1G>A* was associated with a reduced risk of statin prescription but not any other endpoint (**Supplemental Figure 4B**).

### Step 3

#### Common variants in *ANGPTL3*, *ANGPTL4*, and *APOC3* share their metabolic fingerprint with deleterious variants

In line with a previous investigation (45), we found no significant association between ANGPTL3 inactivation via common variants and CAD. Previously, however, evidence was presented that deleterious variants in *ANGPTL3* are associated with a decreased risk of CAD (46, 47). As the common variants adjacent to *ANGPTL3* only modestly impacted plasma lipids, it could be argued that they do not accurately reflect the effects of more profound *ANGPTL3* inactivation. Therefore, we examined whether common variants adjacent to *ANGPTL3*, *ANGPTL4*, and *APOC3* mimicked the effects (that is, showed the same effect directionality) of deleterious variants.

All common variants adjacent to *ANGPTL3*, *ANGPTL4*, and *APOC3* were highly concordant with deleterious variants within the same gene (**Figure 5A-C**). One hundred sixty-seven metabolite associations near *ANGPTL3* showed a high concordance metric (R^2^) of 84% between the common variants and deleterious models. *ANGPTL4* common variants were also highly concordant with *ANGPTL4* deleterious variants, having an R^2^ of 86%. *APOC3* showed a concordance metric R^2^ of 87%. These results demonstrate that the common genetic variations adjacent to *ANGPTL3*, *ANGPTL4,* and *APOC3* would be valid genetic instruments reflecting a modest “knock-down” of each respective gene.

**Figure 5.**
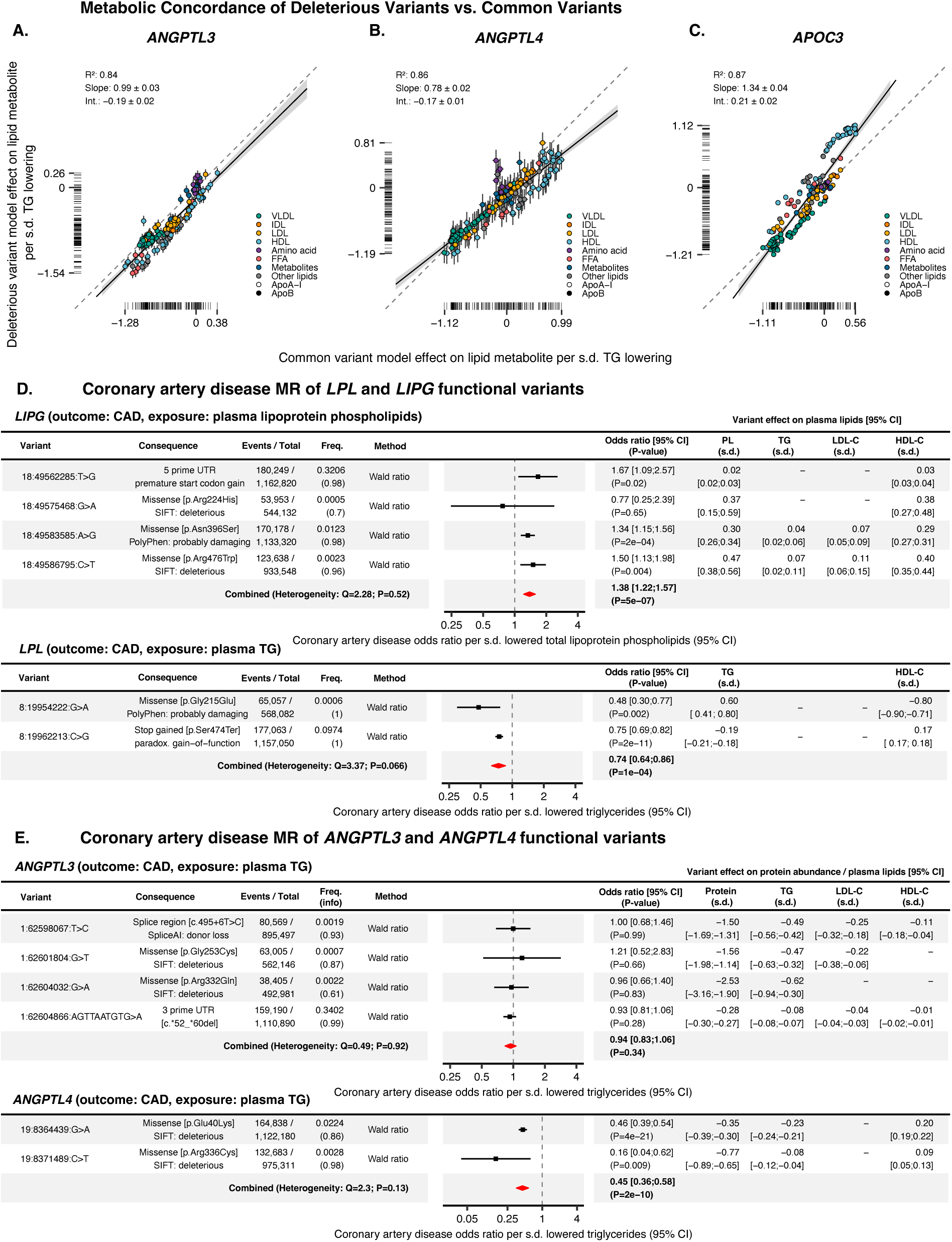
Results of validation analyses. The concordance between the effect directionality of CVs and PTVs is displayed using scatter plots with a regression line. **A:** Comparison of the effect directionality between *ANGPTL3* CVs and PTVs. **B:** *ANGPTL4* CVs vs. PTVs. **C:** *APOC3* CVs vs. PTVs. ’R^2^’ represents the coefficient of determination. ‘Int.’ indicates the regression line intercept. The color of the scattered dots indicates the lipid class of the NMR parameter. The collapsing model estimates were scaled by their 1-s.d. effect on plasma TGs to improve interpretability. **D:** Forest plots and tables showing the results of the CAD MR analysis focusing on functional variants in *LPL* and *LIPG*. Genetic association summary statistics of *LIPG* with the exposure were extracted from the UK biobank NMR study of 115,078 individuals retrieved from (36). *LPL* variant associations were retrieved from the same data set. CAD data was from the Aragam et al. meta-analysis (30). **E:** Forest plots and tables showing the results of the CAD MR analysis that limited the selection of genetic instruments to functional variants in *ANGPTL3* and *ANGPTL4*. “Freq.” represents the alternative allele frequency. “info” represents the imputation quality metric derived from the outcome GWAS. Variant effects on plasma lipids were retrieved from (23). The *ANGPTL4* p.Glu40Lys (p.E40K) estimates differ slightly from Figure 2 because a slightly different estimator and UK Biobank subcohort were used to measure the association between the functional variants and plasma TGs.

#### Comparative drug-target MR of LPL and endothelial lipase (EL) reveals that in order to achieve CAD benefits, ANGPTL3 inhibition should primarily target LPL rather than EL

ANGPTL3 targets both EL and LPL and may thus influence CAD via two independent pathways (48). To compare the effects of these two target enzymes, we analyzed the effects of genetically instrumented EL and LPL activity on CAD by performing functional-variant limited MR of the *LIPG* (encoding EL) and *LPL* genes. We used the preferred enzyme substrate as the exposure, as EL prefers lipoprotein phospholipids, whereas LPL primarily hydrolyzes lipoprotein TGs (49). We detected two functional *LPL* variants and four functional *LIPG* variants with small to large effects on plasma TG/lipoprotein phospholipids (range: 0.02 – 0.6 s.d. per allele).

MR analysis of *LPL* and *LIPG* found opposing significant associations with CAD for LPL (IVW meta-analysis: OR: 0.74, P=1×10^-4^) and EL (IVW meta-analysis OR: 1.38, P=5×10^-7^) (**Figure 5D**). These findings suggest that increased activity of LPL protects against the development of atherosclerosis, whereas heightened activity of EL may contribute to the acceleration of atherosclerosis. The contrasting impact of genetic EL and LPL activity on CAD risk suggests that for ANGPTL3 inactivation to lower CAD risk, it may need to have a greater impact on LPL activity compared to EL activity.

### Step 4

#### Deleterious variants in *ANGPTL3*, *ANGPTL4*, and *APOC3*, and the risk of CAD

Two previous studies found that deleterious variants in *ANGPTL3* protected against CAD (3, 46). In an effort to reproduce these findings, we performed a sensitivity MR analysis of CAD and limited the selection of genetic instruments to functional variants. Functional annotations were detected for four *ANGPTL3*, two *ANGPTL4*, and one *APOC3* variant. The detected *APOC3* variant was the *c.55+1G>A* splice donor loss variant, which was already reported in **Figures 2-4**. The other variants associated with lowered protein levels and triglycerides, with effect sizes ranging from profound to modest (protein range: -2.53– -0.28; TG range -0.28– -0.62) (**Figure 5E**) (22). The variants were analyzed individually and together using random-effects IVW meta-analysis. *Cis*-pQTL MR of the *ANGPTL3* variants indicated that ANGPTL3 protein levels were not significantly associated with CAD, individually or together (meta-analysis IVW OR per s.d. TG: 0.94, P=0.34) (**Figure 5E**). By contrast, reduced *ANGPTL4* protein levels were associated with a decreased risk of CAD (meta-analysis IVW OR: 0.45 per s.d. TG, P=2×10^-10^) (**Figure 5E**).

Considering the beneficial effects of ANGPTL3, ANGPTL4, and APOC3 on plasma lipids, it was expected that genetic inactivation of these proteins would confer protection against CAD. However, the ANGPTL3 MR analyses focusing on common variants and MR of functional variants (identified through DNA microarrays) did not support this hypothesis. Therefore, we pursued a meta-analysis of DNA sequencing-based studies that studied the effect of *ANGPTL3*, *ANGPTL4*, and *APOC3* deleterious variants on CAD. The rationale for excluding DNA microarray and exome bead chip-based studies was the potential risk of introducing measurement error for rare variants (50, 51), leading to bias towards the null hypothesis. DNA-sequencing-based substudies from previous papers (3, 46, 52, 53), were extracted and analyzed together with genetic association analyses conducted in the UK Biobank. Loss-of-function variant genetic association effect sizes typically range from -1 to -3 s.d. for their affected protein (22). The carrier status of deleterious variants was associated with substantial decreases in protein levels for both *ANGPTL3* (-1.29 s.d. protein, P=9×10^-32^) and *ANGPTL4* (-1.33 s.d. protein, P=1.4×10^-43^). APOC3 protein levels were not measured in the UK Biobank. However, *APOC3* deleterious variants were associated with a significant reduction in TG (-0.68 mmol/L TG, P=1×10^-82^). .

The results of the meta-analysis are presented in **Figure 6**. The presence of *ANGPTL3* deleterious variants was associated with reduced CAD risk (meta-analysis IVW OR: 0.41 per TG, P=3×10^-5^). *ANGPTL4* deleterious variant carrier status, excluding p.E40K, was also associated with a reduced risk of CAD (meta-analysis IVW OR: 0.32 per TG, P=0.016), as was *APOC3* deleterious variant carriers status (meta-analysis IVW OR: 0.70 per TG, P=0.005). The key finding was the robust association of *ANGPTL3* deleterious carrier status with a reduced risk of CAD. This association was not detected with the other approaches and implies that ANGPTL3 lowering might offer atheroprotective benefits similar to ANGPTL4 or APOC3 lowering.

**Figure 6.**
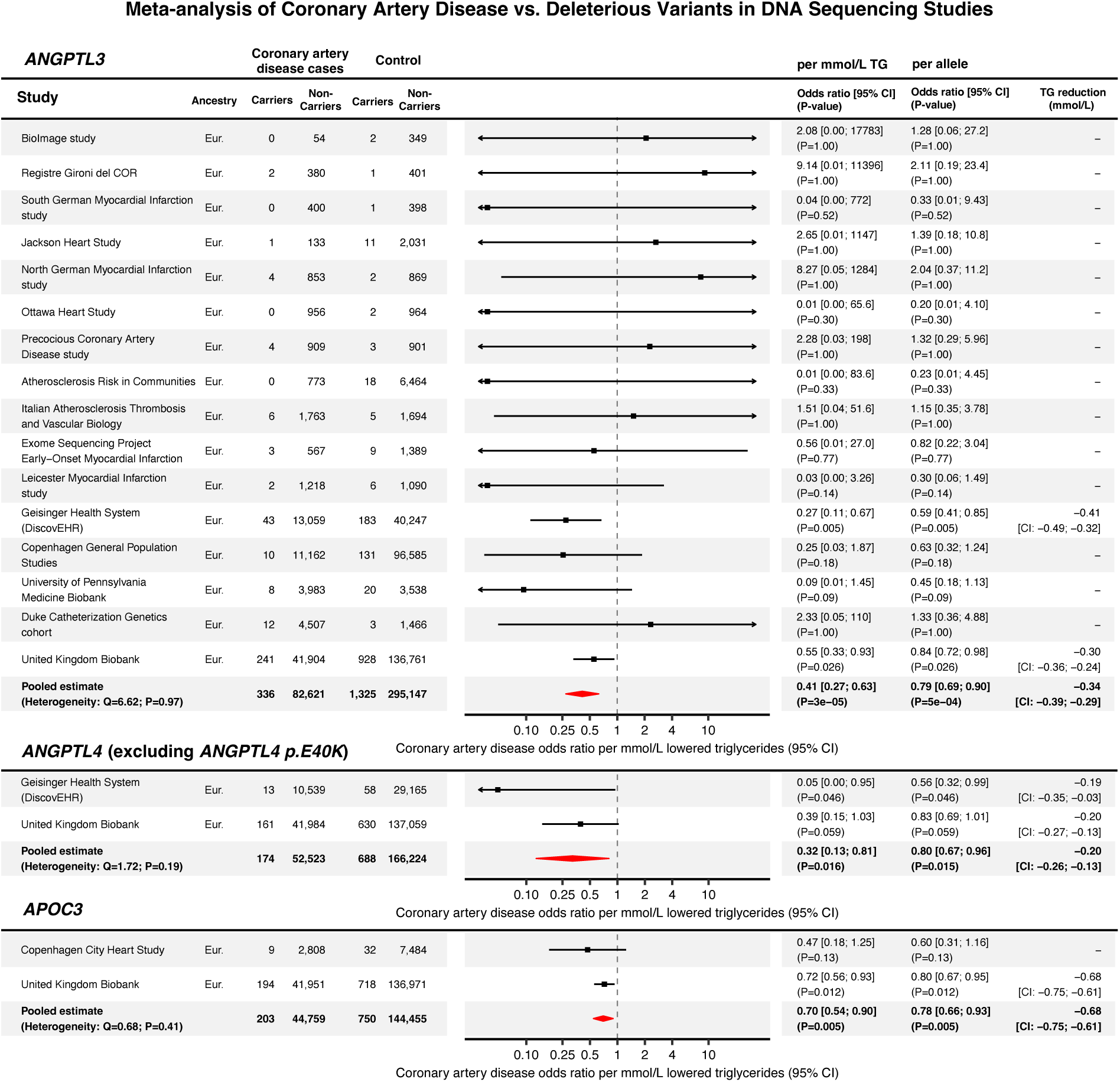
Meta-analysis of deleterious variants in ANGPTL3, ANGPTL4, and APOC3, and the risk of CAD. Forest plots and tables indicating the effect on CAD per mmol/L TG, and per allele. The deleterious variant effect estimates for each substudy were retrieved from (3, 23, 46, 52, 53). The case definition used in the Copenhagen City Heart Study was not exclusively restricted to CAD. 21% of the ischemic vascular disease cases were diagnosed with atherosclerotic cerebrovascular disease, rather than CAD (with CAD encompassing 79 % of the cases) (53). ‘Eur.’ denotes European ancestry.

## DISCUSSION

We find that targeted inactivation and associated lowering of plasma APOC3 levels is predicted to decrease plasma TG and LDL and raise HDL levels. Targeted lowering of plasma ANGPTL3 is expected to reduce plasma TG, LDL, and HDL levels, while lowering of plasma ANGPTL4 is predicted to decrease plasma TG and increase HDL levels. Based on these findings, it is expected that genetic inactivation of APOC3, ANGPTL3, and ANGPTL4 levels is associated with protection against CAD. Through MR and a meta-analysis of rare variant genetic association studies, we confirmed that targeted inactivation and lowering of ANGPTL3, ANGPTL4, and APOC3 is associated with a lowered risk of CAD. In addition, lifetime genetic lowering of ANGPTL4 was observed to reduce the risk of T2D, indicating that ANGPTL4 inhibition might provide additional benefits to patients with T2D and dyslipidemia.

The inactivation of ANGPTL4 in mice and monkeys was shown to lead to mesenteric lymphadenopathy and other severe complications. Naturally, these observations raised serious concerns about the safety of pharmacological targeting of ANGPTL4. Here, we did not find an association between genetic ANGPTL4 inactivation and several disease codes related to lymphatic disorders. While these data do not entirely exclude any harmful effects of ANGPTL4 inactivation, they do mitigate safety concerns about the impact of whole-body inactivation of ANGPTL4 in humans. Recently, it was shown that silencing of ANGPTL4 in the liver and adipose tissue using ASO markedly reduces plasma TG levels in mice yet does not lead to mesenteric lymphadenopathy or other complications (16). These data suggest that liver- and adipose tissue-specific inactivation of ANGPTL4 may confer similar cardiovascular benefits as whole-body ANGPTL4 inactivation without any particular safety risks.

The association of ANGPTL4 with T2D was distinct from the other proteins that inhibit LPL. In preclinical studies, mice overexpressing LPL in muscle were more insulin resistant, while mice lacking LPL in muscle were more insulin sensitive. In contrast, mice overexpressing LPL in adipocytes were more insulin sensitive (54, 55). The protective effect of enhanced LPL action in adipose tissue may be related to increased lipid partitioning into the adipose tissue and reduced ectopic fat. While ANGPTL3, ANGPTL4, and APOC3 all act through LPL, only ANGPTL4 acts exclusively via LPL, which may explain why only genetic variation in ANGPTL4 is associated with T2D risk.

Previous studies reported conflicting findings regarding the association between ANGPTL3 and CAD. Dewey et al. (3), and Stitziel et al. (46) found that rare deleterious *ANGPTL3* variants were associated with decreased odds of ASCVD, whereas MR studies of common ANGPTL3-lowering variants reported negative findings (45). By meta-analysis of deleterious variant genetic association studies, we found clear, statistically robust evidence that lifetime genetic inactivation of ANGPTL3 confers protection against CAD. These findings align with recent case reports indicating that ANGPTL3 lowering with Evinacumab protects against atherosclerosis progression in HoFH patients (11, 12). A recent and similar UK Biobank study examining the impact of protein-truncating *ANGPTL3* variants on CAD found no association (56). Compared to their analysis, key differences were a broader case definition, a stricter definition of controls, the inclusion of deleterious missense variants, and the meta-analysis, which incorporated evidence from previous studies. These methodological differences strengthened statistical power in our study, making a false negative finding less probable.

The discrepancy between the deleterious and common ANGPTL3 variants in terms of their association with CAD could be due to a range of different factors. One possible explanation is the pleiotropic effects of ANGPTL3. Besides inhibiting LPL, ANGPTL3 inhibits endothelial lipase (EL) (57). In a recent paper, we showed that the LPL-independent effects of ANGPTL3 inactivation on plasma metabolic parameters showed a striking inverse resemblance with EL inactivation, suggesting that ANGPTL3 modulates plasma lipid levels by inhibiting LPL and EL (41). Here, using MR, we compared the effects of genetically instrumented EL and LPL activity on CAD. Whereas increased LPL activity reduced the odds of CAD, increased EL activity increased the odds of CAD. The observed link between EL and CAD is consistent with previous human genetic studies showing the possible harmful effects of a genetically predicted increase in EL activity (58, 59). This suggests that ANGPTL3’s interaction with EL might counteract its cardiovascular benefits achieved through LPL inhibition under certain physiological conditions. While our research demonstrated metabolic concordance between ANGPTL3 common variants and deleterious variants, it remains possible that more profound ANGPTL3 inactivation by deleterious variants could tip the balance in favor of LPL inhibition over EL. This shift could potentially enhance the anti-atherosclerotic benefits of ANGPTL3 lowering.

Interestingly, the association of ANGPTL3 inactivation with CAD was only present for rare deleterious variants when the carrier status was determined by DNA sequencing. This exposes the limitations of drug-target MR studies using DNA micro-array-based GWAS. When rare variants are incorrectly imputed, this typically introduces a one-sided loss of information that biases toward the null hypothesis, leading to falsely negative findings (51). Even though the imputation quality score (e.g., ‘INFO’) reports an imputation quality metric, this metric does not really measure the true imputation accuracy (60). The imputation accuracy can only truly be determined if variant carrier status is called by genotyping (e.g., via DNA sequencing). However, studying rare variants in genetic association studies is not without drawbacks. An important limitation of rare variants is statistical imprecision simply due to their rarity (61). Rare variants also often emerged relatively recently and consequently are more susceptible to confounding by enrichment in specific geographical regions, families, or socioeconomic strata (62). Even if appropriate model adjustments are applied, subtle differences in population structure could cause a small number of extra alleles to be present in the control (or case) group. This can lead to biased estimates when the rare alternative allele is present in ten, or hundred individuals in total, which is often the case for rare variant studies even when the total sample size is above hundreds of thousands. Overall, our findings underscore the importance of combining evidence from rare deleterious and benign common variants in genetic association studies of complex disease phenotypes.

The complexity of the *APOA1*-*APOA5*-*APOC3* locus and the potential confounding due to LD poses significant challenges in separating the genetic association signals. The use of *APOC3* c.55+1G>A as a genetic instrument was justified because of its independence from common variants within this region, making it ideal for studying APOC3 inactivation specifically. On the other hand, the analyses of APOC3 inactivation that did not include the c.55+1G>A variant should be interpreted with caution. Compared to clinical trials, MR analysis can exaggerate the magnitude of the effect of inactivating a gene/protein (63). Cis-pQTL MR utilizing protein-coding variants warrants extra carefulness due to the possibility of epitope-binding artifacts, which may complicate the precise interpretation of effect sizes. The p.E40K coding variant was the only variant qualifying as a *cis*-pQTL in the *ANGPTL4* region in the Step 1-2 analyses. While our validation analyses suggested this specific association was not attributable to epitope-binding artifacts, we still advise caution when extrapolating effect sizes from the analyses. Additionally, MR and other genetic association studies estimate lifelong exposure to changed gene function, while drug trials typically last 2–5 years in late adulthood. If the treatment effect multiplicatively interacts with time, MR may exaggerate it. This constraint should be considered when translating MR findings to predict the results of clinical trials. For ANGPTL4, Dewey et al. (52), found that the TG levels of p.E40K homozygotes were reduced by 0.58 mmol/L (0.92 mmol/L for p.E40K homozygotes vs. 1.49 mmol/L in non-carriers; relative change -39%) in a normotriglyceridemic population. When translating these findings (TG reduction of 0.58 mmol/L) onto the effect size on CAD found in this study, one would expect that lifetime *ANGPTL4* inactivation—in a population of normotriglyceridemic individuals—results in a risk reduction corresponding to a CAD odds ratio of 0.61 (95% CI: 0.52–0.72).

In conclusion, our genetic analysis predicts that in a broader dyslipidemic patient population, therapies aimed at decreasing plasma ANGPTL3, ANGPTL4, or APOC3 levels will be effective in preventing CAD without raising specific safety concerns. In addition, therapies aimed at reducing plasma levels of ANGPTL4 may provide additional benefits to patients with dyslipidemia and T2D.

## ETHICAL REVIEW

This study analyzed scientific data that is available to the public, as detailed in **Table 1** and **Supplemental Table 1**, where references to the specific datasets can be found. Analyses using individual-level access to the UK Biobank Resource were conducted under Application Number #148828. The Erasmus Rucphen Family study was approved by the medical ethics board of the Erasmus MC Rotterdam, the Netherlands (64). All studies complied with the ethical standards outlined in the Helsinki Declaration.

## ACKNOWLEDGEMENTS

We would like to thank Prof. Cornelia van Duijn and Prof. Ko Willems van Dijk for providing access to samples from the Erasmus Rucphen Family study. We would like to thank the participants and researchers of the CARDIoGRAMplusC4D, CKDGen, deCODE, DIAGRAM, EPIC-CVD, FinnGen, GIGASTROKE, GLGC, and UK biobank studies, as well as the other non-consortium studies.

## DATA AVAILABILITY STATEMENT

Database identifiers and links to the public data sets are provided in **Supplemental Table 1**. Access to UK Biobank data is limited to authorized researchers who comply with data use policies to safeguard participant confidentiality. Interested researchers must apply to UK Biobank, adhering to an application process that ensures ethical and legal compliance in data handling. UK Biobank data set identifiers used for the analyses under Application Number #148828 are provided in the **Supplemental Methods**. The data from the Erasmus Rucphen Family study (ERF) cannot be shared publicly due to data protection laws. Access to this data is restricted to ensure participant confidentiality, aligning with legal and ethical obligations. Any request for access to the data for legitimate scientific purposes can be directed to the principal investigators of the ERF study, subject to a rigorous review process ensuring that all legal and ethical standards are met. The analyses in this manuscript were performed using the R programming language (v.4.2.1) with the packages coloc, cowplot, data.table, ggplot2, ggthemes, mungegwas, phewas, twosamplemr, wesanderson, writexl, and the Python programming language (v.3.8.16) using the packages numpy, pandas, and scipy. The LD matrix estimates were calculated using PLINK (v1.90b6.24).

## APPENDICES

### Supplemental Note

This document contains the **Supplemental Methods**. The supplemental note *.pdf* also contains **Supplemental Figures *1–6***.

### Supplemental Figures

*The Supplemental Figures 1–6 are contained within the supplemental note .pdf file*.

**Supplemental Figure 1. Scatter plots showing the results of the drug-target MR analyses.** Each subplot represents the results of the analyses displayed in **Figures 2-4** of the main manuscript that used more than one genetic instrument.

**Supplemental Figure 2. Regional genetic association plots showing the results of the colocalization analyses.** Each subplot represents the results of the analyses displayed in **Figure 2-4** of the main manuscript.

**Supplemental Figure 3. Results of cis-pQTL MR analyses of the estimated glomerular filtration rate by (eGFR) by Cystatin C and plasma Creatinine.** The findings are displayed in bar graphs, illustrating the level of the effect per s.d. decrease in protein abundance. The red lines represent the 95% CI.

**Supplemental Figure 4. Results of phenome-wide MR analysis on FinnGen outcomes using ANGPTL3 rs34483103-1:62604866:AGTTAATGTG>A [3 prime UTR, c.*52_*60del] and APOC3 *rs138326449-11:116830638:G>A* [donor loss, c.55+1G>A] variants. A**: Volcano plot displaying the results of ANGPTL3 cis-pQTL phenome-wide MR. **B**: APOC3 cis-pQTL phenome-wide MR volcano plot. The y-axis solid straight lines indicate the phenome-wide significance threshold. ‘OR’ indicates the odds ratio with 95% confidence intervals with Bonferroni correction for the 694 FinnGen outcomes.

**Supplemental Figure 5. Cis-pQTL MR of the clinical laboratory outcomes, showing the results on a 1-s.d. scale.** The red bars indicate the 95 % CI. The black dots indicate the effect point estimate. ‘*’ indicate P < 0.05. ‘**’ indicates P < 0.05 with a shared causal variant (H_4_). A list explaining the abbreviations is provided in the supplemental material (**Supplemental Table 1**).

**Supplemental Figure 6. Wildtype human ANGPTL4 protein and the ANGPTL4 E40K variant protein are detected to a similar extent by the antibody used in the human ANGPTL4 ELISA.** Western blot of the precipitated medium of HEK293 cells transfected with expression vectors for different mutant forms of human ANGPTL4 fused to a V5-tag (65). CC/AA is an oligomerization defective ANGPTL4 variant. SDS-PAGE was performed using a loading buffer without DTT or other disulfide-reducing agent. Equal amounts of medium were loaded. Membranes were blotted with an antibody from R&D Systems that is used for the human ANGPTL4 Elisa (AF3485, 1:2500). Secondary antibody was goat anti-rabbit IgG conjugated to HRP (1:5000).

### Supplemental Tables

*The Supplemental Tables 1-10 are located within the supplemental_tables_1_10.xlsx file. The .xlsx file is available via a public repository and can be accessed via the following link:* https://doi.org/10.5281/zenodo.10880711

**Supplemental Table 1. Description of GWAS data sets**

List of the GWAS data sets used. ’database.ID’ denotes the EBI GWAS catalog identifiers if starting by “GCST”, or the UK biobank showcase identifier if starting by a number.

**Supplemental Table 2. MR results with 1000Genomes LD matrix**

Results of the MR analyses that are presented in the paper. ’rsids.dbsnp.ver144’ denote the rsids that were used in each MR model.

**Supplemental Table 3. MR sensitivity analysis w. UKB 337K LD matrix vs. 1000Genomes.**

LD matrix sensitivity analysis using the 1000Genomes and UKB 337K LD matrices.’heterogeneity.statistic’ indicates the heterogeneity P-value, which is described in detail in the supplemental note.

**Supplemental Table 4. MR sample overlap risk of bias analysis**

Result of the risk of bias from sample overlap analysis reporting the F-statistics, bias (’Beta.bias.low.scenario’, ’ Beta.bias.medium.scenario’, and ’ Beta.bias.high.scenario’) and type 1 error inflation (Type1error.lowbias.scenario, Type1error.mediumbias.scenario, Type1error.highbias.scenario) for each scenario and model.

**Supplemental Table 5. ICD codes used for UKB lymphadenopathy-related phecodes**

Phecode and ICD-codes as described in the Methods section.

**Supplemental Table 6. ANGPTL4 p.40K pQTL MR PheWAS results Detailed results of the MR analyses presented in Figure 4D**.

**Supplemental Table 7. ANGPTL4 p.Cys80fs pQTL MR PheWAS results**

Detailed results of the MR analyses presented in **Figure 4D**.

**Supplemental Table 8. *ANGPTL3 rs34483103-1:62604866:AGTTAATGTG>A* [3 prime UTR, c.*52_*60del] pQTL MR PheWAS results**

Detailed results of the MR analyses presented in **Supplemental Figure 4A**.

**Supplemental Table 9. *APOC3 rs138326449-11:116830638:G>A* [donor loss, c.55+1G>A] pQTL MR PheWAS results**

Detailed results of the MR analyses presented in **Supplemental Figure 4B**.

**Supplemental Table 10. List of variants used in the UK Biobank genetic association analyses** ’rsID’ indicates the NCBI Reference SNP cluster ID. ’REVEL’ denotes the Rare Exome Variant Ensemble Learner pathogenicity prediction score for missense variants.

