## Supplemental Note for "Drug-target Mendelian randomization analysis supports lowering plasma ANGPTL3, ANGPTL4, and APOC3 levels as strategies for reducing cardiovascular disease risk"

### SUPPLEMENTAL METHODS

The supplemental methods section is presented below.

#### Data sources (Step 1-2)

***Selection of GWAS***

The GWAS were predominantly meta-analyses of case-control and population-based cohort studies, including males and females. Genotyping was conducted using different DNA microarrays and exome-sequencing platforms. Our quality control procedures included updating the genomic reference to GRCh38 coordinates, effect and reference allele harmonization, and filtering indels and variants with an alternative allele frequency < 0.1%. We restricted the analyses to GWAS of European ancestry populations to reduce potential bias from population stratification. The exception was the Cystatin C-based estimated glomerular filtration rate (eGFR) GWAS that included a small subset (~16 %) of non-European ancestry individuals (**Table 1**). We chose studies with the largest effective sample sizes for outcomes. Here, one exception was the type 2 diabetes GWAS, where an earlier study (1), was given priority over a later study (2), due to a higher degree of SNP overlap between the exposure and outcome GWAS. For cardiometabolic risk factors, we selected GWAS based on the sample size while also considering if the exposure was measured on a clinically interpretable scale (for example, mmol/L was prioritized over standard deviations [s.d.] when possible). Supplemental Table 1 provides trait definitions, EBI GWAS catalog, and UK biobank trait IDs.

***Assessment and diagnostic criteria for diseases***

The primary outcomes of this study were CAD, CKD, ischemic stroke, non-alcoholic fatty liver disease (NAFLD), and T2D. Prevalent or incident CAD was defined according to the international classification of diseases (ICD-9, ICD-10) codes, or hospital registry codes, relating to the diagnosis of, complications, and procedures related to myocardial infarction and chronic coronary syndrome (primarily ICD-10 I20-I25, see reference for details) (3). CKD was defined as hospital inpatient records connected to chronic kidney disease (ICD-10 N18.*) (4). NAFLD status was defined according to health registry or hospital record ICD-10 codes of fatty liver disease (K76.0), hepatic fibrosis (K74.0 and K74.2), non-alcoholic steatohepatitis (NASH; K75.9), or other specified liver disease (K76.9), while excluding cases with codes related to a wide range of secondary causes of liver disease, for example, alcoholism or viral hepatitis (5). Ascertainment of ischemic stroke in GIGASTROKE included analysis of health registry and hospital records corresponding to any cerebral infarction (ICD-10 codes I63.*), according to radiographic criteria, and other methods (see reference for further details) (6). T2D definitions included health record diagnoses corresponding to T2D (for example, ICD-9 250.*0), Hb1Ac or plasma glucose-based, WHO criteria, and physician- or patient-reported T2D (1). The UK Biobank outcomes that may be associated with abdominal lymphadenopathy were derived from hospital in-patient records. Related ICD-10 diagnoses (**Supplemental Table 5**) were grouped into outcomes using Phecode Map 1.2 (7). The FinnGen abdominal lymphadenopathy-associated phenotypes were ascertained through Finnish health registries of lymphadenitis (ICD-10: L04.*, ICD-8, 9: 683), acute peritonitis (ICD-10: K65.*, ICD-8, 9: 567), intestinal malabsorption (ICD-10: K90.*, ICD-9: 579, ICD-8: 269[1-2]), and other noninfective disorders of lymphatic vessels and lymph nodes (ICD-10: I89.*, ICD-8, 9: 457) (8).

***Assessment and definitions for continuous exposures and outcomes***

This study examined the following continuous exposures and outcomes: plasma protein abundance, routine clinical chemistry including plasma lipid concentrations (TGs, LDL-C, HDL-C, ApoB, ApoA1, Lipoprotein a), systolic and diastolic blood pressure (SBP/DBP), anthropometric traits (Body-mass-index [BMI], waist-to-hip ratio [WHR], body fat percentage [BFP]), kidney function (cystatin C and creatinine-based estimated glomerular filtration rate [eGFR]), magnetic resonance imaging (MRI) measurements, and high-throughput nuclear magnetic resonance (NMR) spectroscopy-determined lipid metabolite concentrations. Plasma protein abundance was measured using the SomaScan and Olink platforms (9, 10). These aptamer- and antibody-binding-based platforms report plasma protein abundance in standard deviations (s.d.). The UK biobank quality control and validation procedures of the routine clinical chemistry tests are summarized in (11, 12). SBP/DBP was measured using manual, automatic, and ambulatory methods, and reported as untransformed millimeters of mercury (mmHg) (13). WHR was defined as the waist measurement divided by hip measurement adjusted by various factors as defined by the GIANT consortium protocol (14). BMI was defined as the body weight divided by squared height ($kg/m^2$) (14). BFP was determined using bioelectrical impedance analysis (15). eGFR measurements were derived from natural log-transformed creatinine and cystatin C, using the CKD-EPI or Schwartz formula for creatinine and the Stevens or CKD-EPI formula for cystatin C (16). The Metabolomics had been performed using a commercial NMR spectroscopy-based metabolomics platform from Nightingale Health Ltd., Helsinki, Finland. Over one hundred plasma metabolite parameters are measured on this platform, such as lipoprotein size, subclass lipid contents, and composition, but also free fatty acids, and amino acids (17).

#### Drug-target Mendelian randomization analyses (Step 1-2)

##### *Drug-target MR methodology*

MR is a method of causal inference that uses genetic variants as instrumental variables (IVs) to test the effect of an exposure on an outcome. By exploiting the random assignment of alleles at conception, MR studies can limit the influence of confounding variables, assuming certain key assumptions are satisfied (18). These assumptions include: 1) the genetic variants are reliably associated with the exposure of interest, 2) they are not associated with the factors that may confound the relationship between the exposure and outcome, and 3) the genetic variants do not influence the outcome, independently of the exposure (18).

Drug-target MR analysis is a specific type of MR used to estimate the causal effect of pharmacological silencing of a target gene (19). It requires an emphasis on assumptions different from those of traditional MR. It is less likely to be influenced by horizontal pleiotropy or reverse causality (for example, when the genetic association with the risk factor is secondary to its effect on the outcome, through feedback mechanisms, or via cross-generational effects (20)). This is because, in drug-target MR, we restrict the selection of genetic IVs to a region surrounding the target gene (2.5 kilobase pairs [Kb] in this paper). Additionally, we assume that genetic information travels from DNA to RNA and protein and not the other way around (following Cricks' central dogma of molecular biology). This means that in drug-target MR, the genetic effect on the outcome is presumed to occur distally to the causal chain of events that starts with a variation (the IV) in the target gene that leads to drug-target exposure (the exposure being changed target gene expression, increased/decreased protein abundance, or altered protein function), making bias from horizontal pleiotropy less probable. Likewise, the influence of horizontal pleiotropy is limited because both the variant and exposure (changed target gene expression, increased/decreased protein abundance, or altered protein function) are located within the causal chain given by Cricks' central dogma ([DNA – RNA – protein] – mediators – outcome), making bias from reverse causality less probable.

##### *Generalized least squares estimator*

This study used a Generalized least squares (GLS) inverse-variance weighted (IVW) estimator to measure the association between ANGPTL3, ANGPTL4, and APOC3 protein abundance and health-related outcomes. This method can achieve a more accurate causal estimate using multiple genetic variations from a single genomic region, even if these variants are correlated via linkage disequilibrium (LD) (21). GLS adjusts the estimated effects and their standard errors to account for between-variant correlations caused by LD. To achieve this, the LD correlation matrix is used as weights. The code snippet (R programming language v.4.2.1) used for the analyses of this manuscript is provided below (modified from (21)).

```
> # run cis GLS-MR according to equations in DOI:10.1002/sim.6835
> bx = mr_harmonized$beta.exposure
> by = mr_harmonized$beta.outcome
> byse = mr_harmonized$standard_error.outcome
> rho = LD_correlation_matrix[mr_harmonized$variant_id, mr_harmonized$variant_id]
```

```
> Omega = byse %o% byse * rho
> results <- data.frame(
>           b = solve(t(bx) %*% solve(Omega) %*% bx) * t(bx) %*% solve(Omega) %*% by,
>           se = sqrt(solve(t(bx) %*% solve(Omega) %*% bx)),
>           pval = NA,
>           n.snps = nrow(mr_harmonized)
> )
> results$pval = with(results, 2 * pnorm(abs(b/se), lower.tail = FALSE))
```

**Code snippet legend: 'mr_harmonized' is the harmonized data holding the variant IDs ('variant_id'), the association of the variants with the exposure ('beta.exposure'), the association of the variants with the outcome ('beta.outcome') and its associated standard error ('standard_error.outcome').**

##### *Justification of Genetic IVs used in the phenome-wide MR*

*ANGPTL3 rs34483103-1:62604866:AGTTAATGTG>A* [, c.*52_*60del] is a 3 prime UTR variant associated with reduced circulating ANGPTL3 protein (effect: -0.28 s.d. per allele, 95% CI: -0.30 – -0.27, P: $< 2\times10^{-308}$) (10). *ANGPTL4* p.E40K is a missense variant (2% allele frequency in Europeans) that hinders ANGPTL4's ability to inhibit LPL by destabilizing it after secretion (22). ANGPTL4 p.Cys80fs rs746226153-19:8364556:GC>G [p.Cys80ValfsTer12] is predicted to be a loss-of-function variant with high confidence (gnomAD Genome Aggregation Database v.4.0.0, https://gnomad.broadinstitute.org). *APOC3* c.55+1G>A is a high-confidence predicted loss-of-function variant (gnomAD v.4.0.0). The APOC3 c.55+1G>A splice donor loss variant substantially impacted plasma APOC3 levels (-2.19 s.d. protein $=3.2\times10^{-142}$, **Table 2**). *APOC3* c.55+1G>A status as a loss-of-function variant is further supported by the fact that the variant's estimated effect on plasma triglycerides was considerable (-0.86 mmol/L, $P=3.4\times10^{-157}$; Table 2), and comparable to that of other loss-of-function variants in *APOC3* (23). In addition, the literature reports the *APOC3* c.55+1G>A splice donor loss variant as an *APOC3* loss-of-function variant (24, 25).

##### *Validation of the ANGPTL4 p.E40K coding variants' association with plasma ANGPTL4 protein levels*

The p.E40K variant was the only variant qualifying as a *cis*-pQTL in the *ANGPTL4* region. Including missense variants in MR studies focusing on protein levels warrants careful consideration due to the possibility of epitope-binding artifacts. The association between ANGPTL4 p.E40K coding variant carrier status and plasma ANGPTL4 protein levels was confirmed in a separate study by ELISA using antibodies that were shown by Western blotting to similarly detect wildtype ANGPTL4 and ANGPTL4 containing the E40K substitution, indicating that the association was not attributable to epitope-binding artifacts.

The Western blot showing that wild-type human ANGPTL4 protein and the ANGPTL4 E40K protein are detected to a similar extent by the antibody used in the ANGPTL4 Elisa (AF3485, R&D Systems) is shown in **Supplemental Figure 6.**

The association of the ANGPTL4 p.E40K coding variant with plasma ANGPTL4 protein levels as measured using the abovementioned ANGPTL4 Elisa was determined in the Erasmus Rucphen Family study (26). The study included 106 ANGPTL4 p.E40K carriers and 314 p.E40K non-carriers. The ERF study received approval from the medical ethics board of the Erasmus MC Rotterdam, the Netherlands. The investigations were conducted in compliance with the Declaration of Helsinki. Genotyping was performed using Sequenom iPLEX (MALDITOF, Sequenom Inc. San Diego, USA). To make the scale comparable to the Olink and SomaScan assays, the ANGPTL4 concentrations measured by the ELISA were log2 transformed and then standardized to have a mean of zero and a standard deviation of one. The association was measured using linear regression, including age and sex as covariates. Having one copy of the p.E40K coding variant was associated with -0.45 s.d. plasma ANGPTL4 protein (95% CI: -0.66 – -0.23, $P=4.83\times10^{-5}$). The association was comparable to those measured by the Olink and SomaScan GWAS and thus confirms that plasma ANGPTL4 levels are lower in p.E40K carriers, irrespective of the method (antibody or single-stranded DNA aptamers) used to measure the plasma ANGPTL4 level (**Table 2**).

#### Sensitivity analyses (Step 1-2)

##### *Colocalization analyses*

Drug-target MR substantially relies on the assumption that LD (a phenomenon in which neighboring genetic variants are inherited together more frequently than anticipated by chance (27)) does not confound the association between variant and outcome. In cases where there are distinct genetic variants affecting both the exposure and the outcome, but they are connected through LD, there is a risk of making incorrect conclusions (28). To limit this issue, we performed colocalization analyses, which test whether two independent association signals in the same gene region are consistent with having a single shared causal variant (that is, testing if the association signals are 'colocalized') (29). The results were presented as the posterior probability, expressed in percentages, of five different hypotheses: $H_0$) no genetic association signal for either trait, $H_1$) a signal for the exposure but not the outcome, $H_2$) a signal for the outcome but not the exposure, $H_3$) a signal for both the exposure and outcome, but they do not share a single causal variant, $H_4$) the exposure and outcome share the same single causal variant. When interpreting the results, it is important to bear in mind that the colocalization analysis we applied assumes a single causal variant. If there exist several causal variants in the same locus, the estimator will tend to favor $H_3$. Prior probabilities were set to $p_1 = 1 \times 10^{-4}$, $p_2 = 1 \times 10^{-4}$, and $p_{12} = 1 \times 10^{-5}$, meaning that we assumed that the prior probability that any SNP was causal for either trait was 1 in 1000 and that the prior probability of any SNP was causal for both traits was 1 in 10,000.

##### *LD matrix sensitivity*

Generalized least squared (GLS)-corrected inverse-variance weighted (IVW) MR can be a powerful tool to conduct MR with correlated genetic instruments. However, given how GLS works, the method could be sensitive to which LD matrix is used. Therefore, we conducted a sensitivity analysis using an LD matrix derived from 337-thousand British ancestry individuals in the UK biobank (30). We tested for substantial heterogeneity by 1) looking at if the effect directionality was discordant, and 2) calculating a heterogeneity statistic using the formula below (v), where the resulting *Z* value was used to obtain a P-value from the cumulative distribution function of the normal distribution.

$$\text{(i)} \qquad Z = \frac{Beta_{1000G} - Beta_{UKB\,LD}}{\sqrt{SE^2_{1000G} - SE^2_{UKB\,LD}}}$$

The analysis indicated that the MR results were not LD matrix sensitive (see **Supplemental Table 3**)

##### *Sample overlap bias*

There was a small-moderate sample overlap between the exposure and outcome GWAS' (Supplemental Table 4). Assuming weak MR instruments, there will be bias towards a possibly confounded risk factor-outcome relationship if the samples overlap, and bias towards the null if they do not (31). Even though it is less likely that variants positioned close to the protein-encoding gene would violate the relevance assumption by weak instrument bias, we conducted risk of bias analyses. We assumed a worst-case scenario in which all the participants of the exposure GWAS were also included in the outcome GWAS. F-statistics were calculated to determine instrument strength for each MR model.

Formula (i) was used for the single-variant MRs, where $Beta_{exposure}$ was the genetic association with the exposure, and $SE_{exposure}$ was the corresponding standard error.

$$\text{(ii)} \qquad F = \left(\frac{Beta_{exposure}}{SE_{exposure}}\right)^2$$

For multiple variant MRs, another formula was used (ii), where $N_{sample\ size}$ denotes the sample size and $N_{variants}$ denote the number of variants in the MR model.

$$\text{(iii)} \qquad F = \left(\frac{N_{sample\,size} - N_{variants} - 1}{N_{variants}}\right)\left(\frac{R^2}{1 - R^2}\right)$$

The $R^2$ in (ii) was approximated using formula (iii), where 'MAF' denotes the minor allele frequency.

(iv) $R^2 = 2\,Beta^2_{exposure} MAF(1 - MAF)$

Where the $R^2$ was summed together for uncorrelated instruments in (ii).

For correlated instruments, another formula (iv) was used to approximate the $R^2$, where $B_X^T$ was the transposed vector of the genetic associations with the exposure, and $C$ was the inverse of the LD correlation matrix after Cholesky decomposition, as suggested by Burgess and Thompson (32).

(v) $R^2 = \sum 2(B_X^T\, C)^2\, MAF(1 - MAF)$

The analyses were run using three scenarios of bias of the observational estimate (olsbias) using the code originally written by (31). These scenarios were 'low olsbias', 'medium olsbias', and 'high olsbias', where the effect magnitude of the biased observational estimate was 10%, 50%, and 100% of the MR estimate, respectively.

The analysis indicated that the risk of bias from sample overlap was minimal (see **Supplemental Table 4**).

#### Analyses of deleterious variants in the UK Biobank (Step 3-4)

##### *The UK Biobank resource*

The UK Biobank is a population-based study with a vast repository of research data comprising health record information, genetic information, blood chemistry, and imaging data from approximately 500,000 participants in the United Kingdom (33). Between 2006 and 2010, participants aged 40-69 were recruited, and their blood was drawn upon recruitment. The UK Biobank follows participants, and health information is continuously updated from health records, death certificates, and registries.

##### *Definition of deleterious variants*

Variants from the population variant call files were annotated with gnomAD v.4.0.0. Deleterious variants were defined as protein-truncating variants and UK Biobank allele frequency <0.05 and missense variants with REVEL pathogenicity prediction score ≥0.25 and UK Biobank allele frequency <0.00025. Protein-truncating variants included Variant Effect Predictor high impact consequences "transcript_ablation", "splice_acceptor_variant", "splice_donor_variant", "stop_gained", "frameshift_variant", "stop_lost", "start_lost", "transcript_amplification", "feature_elongation", and "feature_truncation". The resulting variants classified as deleterious are listed in the **Supplemental Table 10.**

##### *Quality control of genetic data*

We applied quality control filters on the UK Biobank exome sequencing population variant call files (UKB data field 23157). First, to avoid batch effects, we filtered variants that did not meet genotype depth of coverage (DP) ≥10 or were marked as missing for more than 90% of the population. For single nucleotide variants (SNVs), we then filtered variants with DP <10, genotype quality (GQ) <20, and binomial test of alternate allele departure from the heterozygous expectation of 0.5 P >$1\times10^{-3}$. For insertions and deletions (indels), we applied filters to exclude variants with DP <10 and GQ <20.

##### *Genetic mimicry analyses in the United Kingdom Biobank (Step 3)*

We conducted genetic mimicry analyses to determine whether the metabolic effects of deleterious and common variants adjacent to ANGPTL3, ANGPTL4, and APOC3 were metabolically concordant. For the deleterious variant models, we selected unrelated individuals of British ancestry with nuclear magnetic resonance spectroscopy (NMR) data available (UKB category 220). Ratios derived from the NMR measurements were excluded. The total number of unique metabolites amounted to 167, and the total cohort encompassed 181,672 individuals. Left-censored NMR measurements were imputed using minimum value imputation. Measurements missing due to citrate peaks, degraded sample, high ethanol, isopropyl alcohol contamination, low glutamine or high glutamate, ethanol, polysaccharide, or unknown contamination were excluded from further analyses. Each metabolite was standardized to have a mean of zero and a standard deviation of one. The effect of each deleterious variant model on the metabolites was estimated using linear regression adjusting for age, sex, age * sex interaction, fasting time at sampling, and the five first genetic principal components (UKB data fields 21022, 22001, 74, and 22009). Each association was scaled for its 1-s.d. effect on plasma triglycerides to ease comparability with the common variant models.
For the common variant models, we included variants within 2.5 Kb of the target genes transcription start site and transcription termination sites, with an allele frequency > 0.01, significantly associated with plasma triglycerides at the genome-wide significance level ($P < 5\times10^{-8}$). Genetic associations between these variants and the NMR metabolites (category: 'met-d-…' .) were extracted through the MRC-IEU open GWAS database (https://gwas.mrcieu.ac.uk). The effect of the common variants on each metabolite per 1-s.d. change of plasma triglycerides was estimated through drug-target MR of the 167 NMR metabolites using triglyceride levels as the exposure, and single metabolites as the outcome. This means the effect on each metabolite is given per 1-s.d. triglyceride change. Finally, the metabolic concordance between the deleterious and common-variant models was estimated using linear regression on the association effect estimates ('betas'). The concordance metric was reported as the coefficient of determination ($R^2$).

***Association of deleterious variants with coronary artery disease in the United Kingdom Biobank (Step 4)***

We conducted gene-collapsing analyses in the UK Biobank to measure the association between *ANGPTL3*, *ANGPTL4*, and *APOC3* deleterious variants and coronary artery disease (CAD). Diagnosis codes were retrieved from the death register, hospital inpatient health records, algorithmically defined myocardial infarction outcomes, and first occurrences data sets (UKB data fields 131271–131423, 40001, 40002, 41202, 41270, 42001, 42003, 42005). We selected non-related British ancestry UK Biobank participants. Cases were defined as positive for ICD-10 diagnosis codes related to ischemic heart disease (I20-I25), only excluding subjects with evidence for non-obstructive ischemic heart disease. The excluded diagnoses were: I20.1: Angina pectoris with documented spasm, I24.8: Other forms of acute ischemic heart disease (which includes I24.81: Acute coronary microvascular dysfunction), and I25.4: Coronary artery aneurysm and dissection. Controls were defined as participants without a positive diagnosis for any phenotype listed in the ICD-10 I00-I99: Diseases of the circulatory system chapter. This strategy was implemented to minimize the risk of contamination resulting from correlated or genetically related diagnoses. After applying these filters, the total UK Biobank CAD cohort amounted to 42,145 cases and 137,689 controls. Of these 179,834 individuals, there were 1169 participants with deleterious *ANGPTL3* variants, 761 with deleterious *ANGPTL4* variants, and 912 with deleterious *APOC3* variants. The association of each deleterious variant model with CAD was measured using logistic regression, adjusting for age, sex, age * sex interaction, and the five first genetic principal components. The associations were reported as the odds ratio per 1-allele.

***Association of deleterious variants with plasma triglycerides, ANGPTL3 and ANGPTL4 protein levels in the United Kingdom Biobank (Step 4)***

We measured the genetic association of deleterious variants in *ANGPTL3*, *ANGPTL4*, and *APOC3* with plasma triglycerides (UKB data field 30870). We also measured the association of deleterious *ANGPTL3* variants with ANGPTL3 protein levels (UKB data field olink_instance_0.angptl3), and the association of deleterious *ANGPTL4* variants with ANGPTL4 protein levels (UKB data field olink_instance_0.angptl4). APOC3 plasma protein levels were not measured in the UK Biobank. The association analyses were conducted in the control group of the CAD analyses described in the section above. This was for two reasons. First, it restricts bias from post-event measurements affecting the gene-triglycerides or gene-protein associations. Second, the over-recruitment of cases can affect the distribution of confounders, which might distort the gene-triglycerides or gene-protein associations.

The ANGPTL3 and ANGPTL4 protein concentrations, measured as Olink log2 normalized protein expression (NPX), were scaled to have a mean of zero and a standard deviation of one. In the control group, there were 131,292 individuals with available plasma triglycerides measurements, of which 886 carried deleterious *ANGPTL3* variants, 601 carried deleterious *ANGPTL4* variants, and 676 carried deleterious *APOC3* variants. There were 13,662 in the control group with available plasma ANGPTL3 protein measurements, of which 93 had a deleterious *ANGPTL3* variant. There were 13,727 with available plasma ANGPTL4 protein measurements in the control group, of which 77 had a deleterious *ANGPTL4* variant. The associations between the deleterious variants and plasma triglycerides, ANGPTL3 or ANGPTL4 protein levels were determined using linear regression adjusting for age, sex, age * sex interaction, fasting time at sampling, and the five first genetic principal components. For triglycerides, the associations were reported as the 1-mmol/L change per 1-allele. For ANGPTL3 and ANGPTL4 plasma protein concentrations, the associations were reported as the 1-s.d. change per 1-allele.

### SUPPLEMENTAL FIGURE 1

**Figure S1. Scatter plots showing the results of the drug-target MR analyses.** Each subplot represents the results of the analyses displayed in **Figures 2-4** of the main manuscript that used more than one genetic instrument.

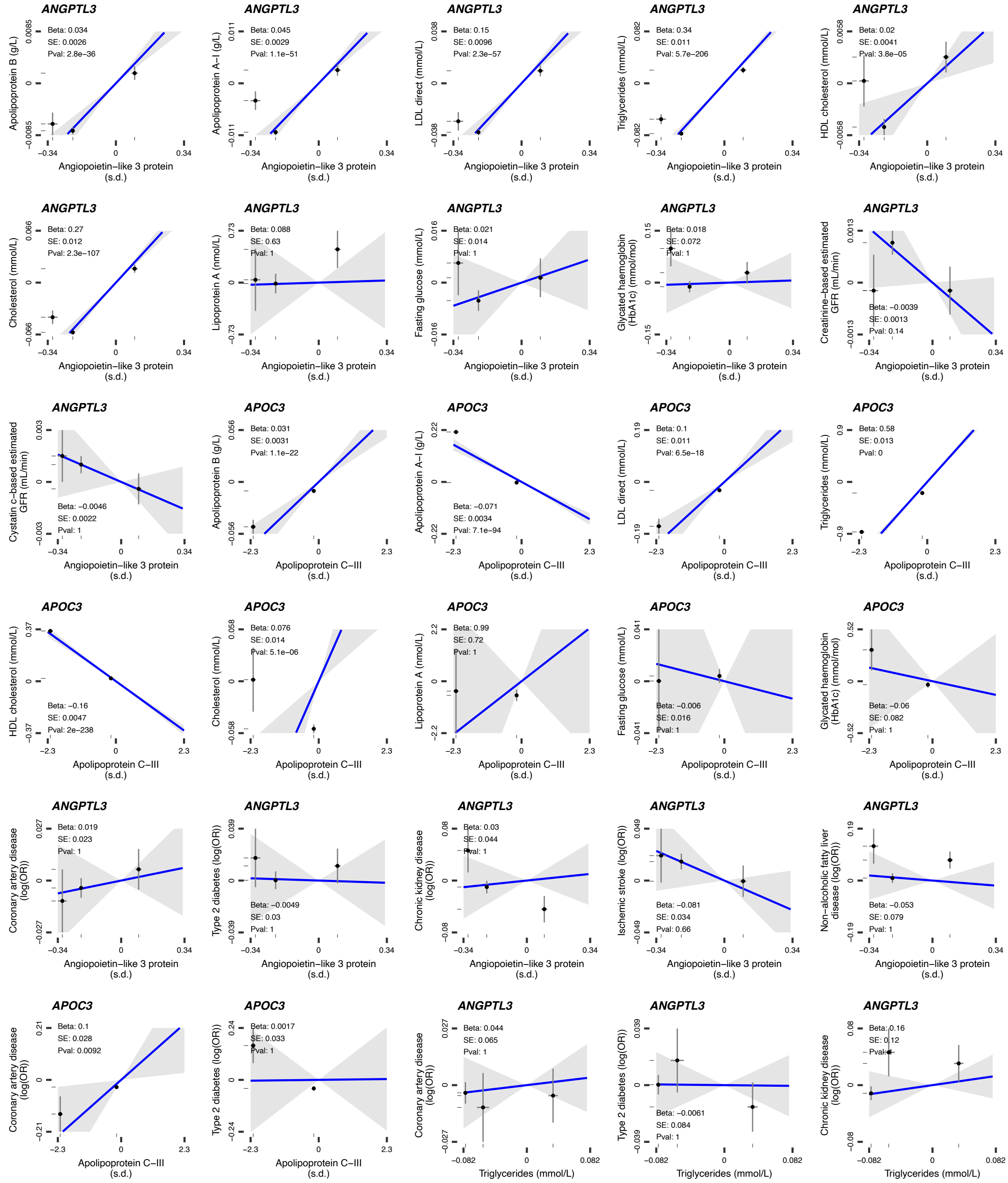

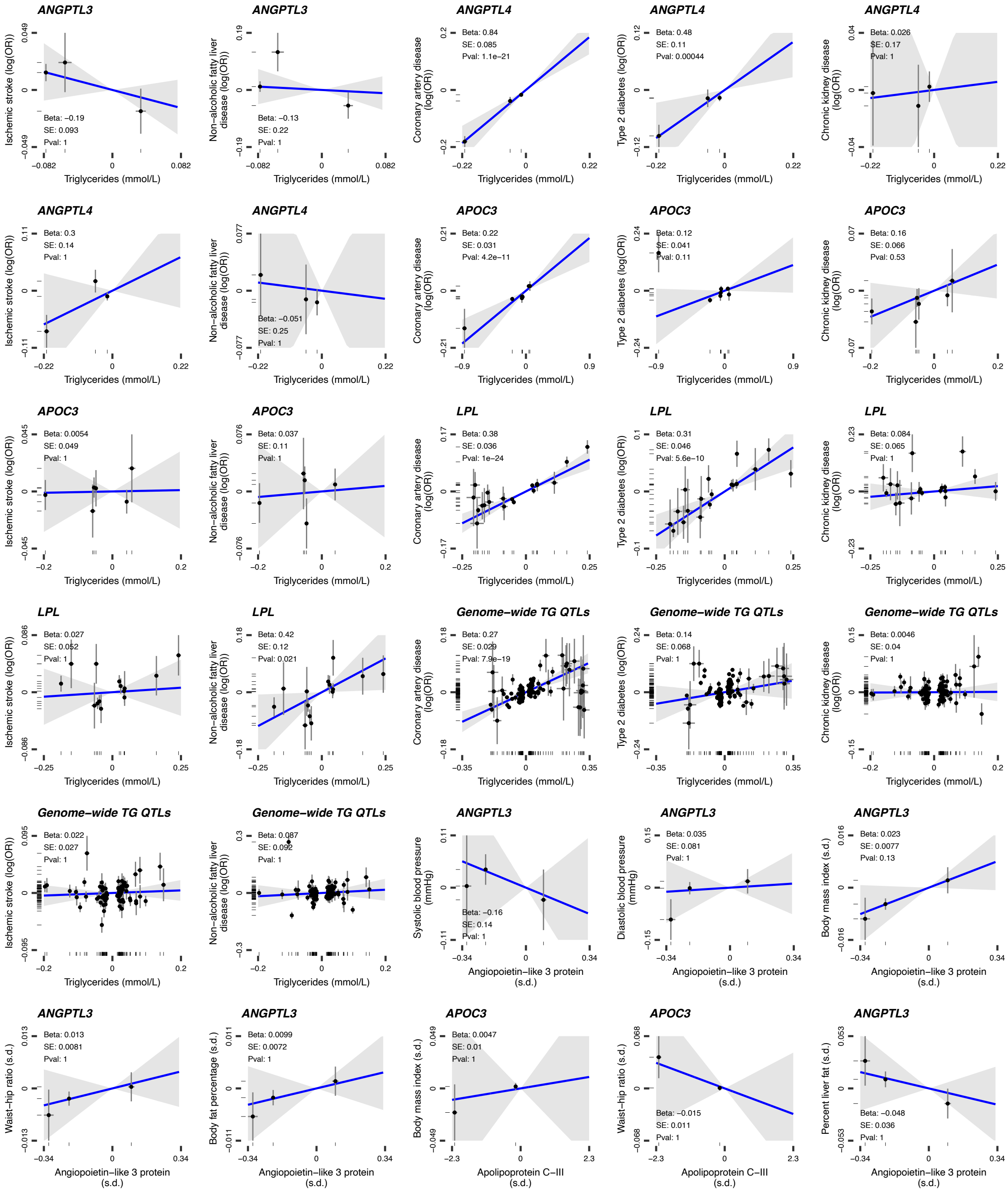
ANGPTL3
Ischemic stroke (log(OR))
Beta: −0.19
SE: 0.093
Pval: 1
Triglycerides (mmol/L)
ANGPTL3
Non−alcoholic fatty liver disease (log(OR))
Beta: −0.13
SE: 0.22
Pval: 1
Triglycerides (mmol/L)
ANGPTL4
Coronary artery disease (log(OR))
Beta: 0.84
SE: 0.085
Pval: 1.1e−21
Triglycerides (mmol/L)
ANGPTL4
Type 2 diabetes (log(OR))
Beta: 0.48
SE: 0.11
Pval: 0.00044
Triglycerides (mmol/L)
ANGPTL4
Chronic kidney disease (log(OR))
Beta: 0.026
SE: 0.17
Pval: 1
Triglycerides (mmol/L)
ANGPTL4
Ischemic stroke (log(OR))
Beta: 0.3
SE: 0.14
Pval: 1
Triglycerides (mmol/L)
ANGPTL4
Non−alcoholic fatty liver disease (log(OR))
Beta: −0.051
SE: 0.25
Pval: 1
Triglycerides (mmol/L)
APOC3
Coronary artery disease (log(OR))
Beta: 0.22
SE: 0.031
Pval: 4.2e−11
Triglycerides (mmol/L)
APOC3
Type 2 diabetes (log(OR))
Beta: 0.12
SE: 0.041
Pval: 0.11
Triglycerides (mmol/L)
APOC3
Chronic kidney disease (log(OR))
Beta: 0.16
SE: 0.066
Pval: 0.53
Triglycerides (mmol/L)
APOC3
Ischemic stroke (log(OR))
Beta: 0.0054
SE: 0.049
Pval: 1
Triglycerides (mmol/L)
APOC3
Non−alcoholic fatty liver disease (log(OR))
Beta: 0.037
SE: 0.11
Pval: 1
Triglycerides (mmol/L)
LPL
Coronary artery disease (log(OR))
Beta: 0.38
SE: 0.036
Pval: 1e−24
Triglycerides (mmol/L)
LPL
Type 2 diabetes (log(OR))
Beta: 0.31
SE: 0.046
Pval: 5.6e−10
Triglycerides (mmol/L)
LPL
Chronic kidney disease (log(OR))
Beta: 0.084
SE: 0.065
Pval: 1
Triglycerides (mmol/L)
LPL
Ischemic stroke (log(OR))
Beta: 0.027
SE: 0.052
Pval: 1
Triglycerides (mmol/L)
LPL
Non−alcoholic fatty liver disease (log(OR))
Beta: 0.42
SE: 0.12
Pval: 0.021
Triglycerides (mmol/L)
Genome−wide TG QTLs
Coronary artery disease (log(OR))
Beta: 0.27
SE: 0.029
Pval: 7.9e−19
Triglycerides (mmol/L)
Genome−wide TG QTLs
Type 2 diabetes (log(OR))
Beta: 0.14
SE: 0.068
Pval: 1
Triglycerides (mmol/L)
Genome−wide TG QTLs
Chronic kidney disease (log(OR))
Beta: 0.0046
SE: 0.04
Pval: 1
Triglycerides (mmol/L)
Genome−wide TG QTLs
Ischemic stroke (log(OR))
Beta: 0.022
SE: 0.027
Pval: 1
Triglycerides (mmol/L)
Genome−wide TG QTLs
Non−alcoholic fatty liver disease (log(OR))
Beta: 0.087
SE: 0.092
Pval: 1
Triglycerides (mmol/L)
ANGPTL3
Systolic blood pressure (mmHg)
Beta: −0.16
SE: 0.14
Pval: 1
Angiopoietin−like 3 protein (s.d.)
ANGPTL3
Diastolic blood pressure (mmHg)
Beta: 0.035
SE: 0.081
Pval: 1
Angiopoietin−like 3 protein (s.d.)
ANGPTL3
Body mass index (s.d.)
Beta: 0.023
SE: 0.0077
Pval: 0.13
Angiopoietin−like 3 protein (s.d.)
ANGPTL3
Waist−hip ratio (s.d.)
Beta: 0.013
SE: 0.0081
Pval: 1
Angiopoietin−like 3 protein (s.d.)
ANGPTL3
Body fat percentage (s.d.)
Beta: 0.0099
SE: 0.0072
Pval: 1
Angiopoietin−like 3 protein (s.d.)
APOC3
Body mass index (s.d.)
Beta: 0.0047
SE: 0.01
Pval: 1
Apolipoprotein C−III (s.d.)
APOC3
Waist−hip ratio (s.d.)
Beta: −0.015
SE: 0.011
Pval: 1
Apolipoprotein C−III (s.d.)
ANGPTL3
Percent liver fat (s.d.)
Beta: −0.048
SE: 0.036
Pval: 1
Angiopoietin−like 3 protein (s.d.)

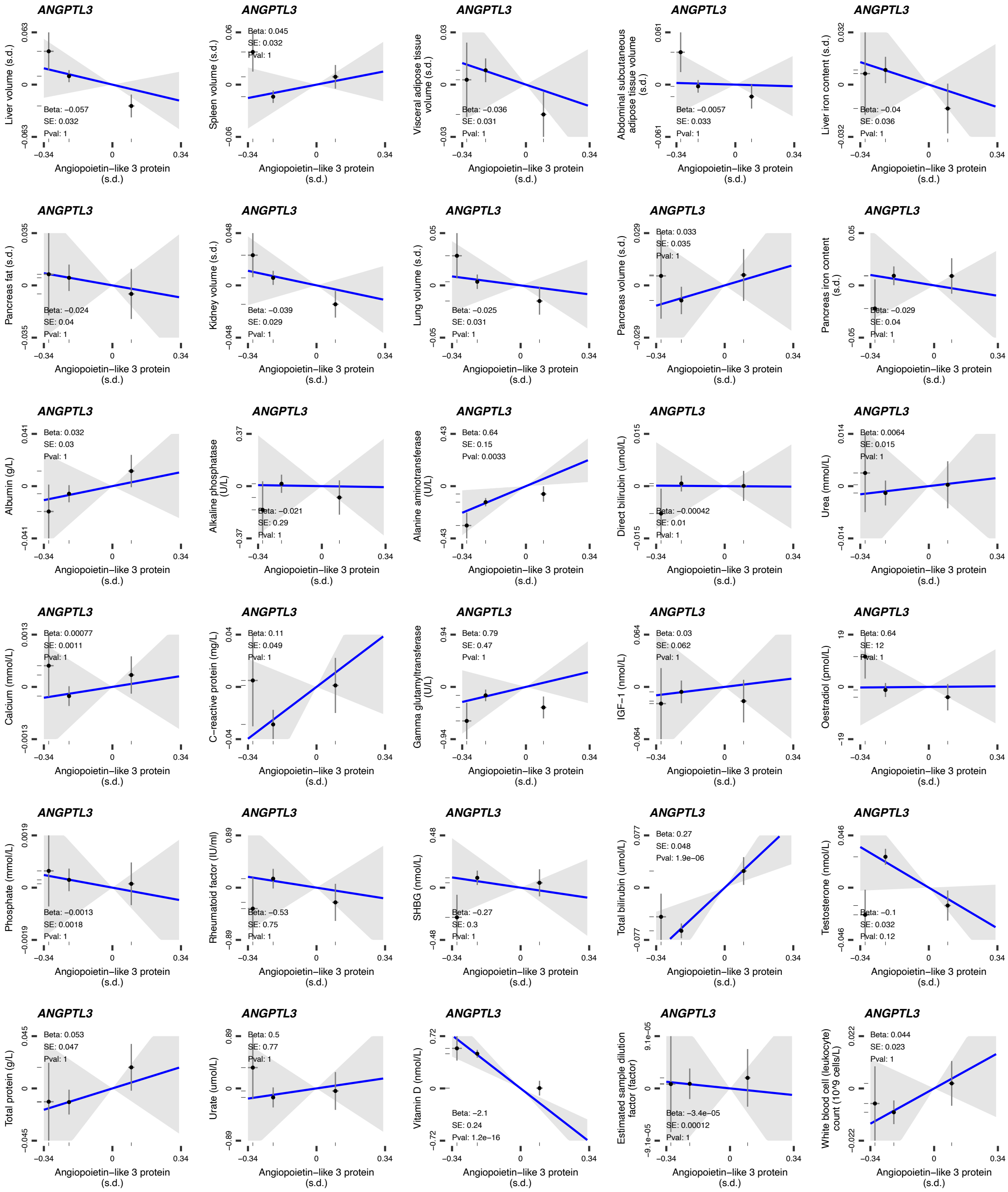
ANGPTL3
Liver volume (s.d.)
Beta: −0.057
SE: 0.032
Pval: 1
Angiopoietin−like 3 protein (s.d.)
ANGPTL3
Spleen volume (s.d.)
Beta: 0.045
SE: 0.032
Pval: 1
Angiopoietin−like 3 protein (s.d.)
ANGPTL3
Visceral adipose tissue volume (s.d.)
Beta: −0.036
SE: 0.031
Pval: 1
Angiopoietin−like 3 protein (s.d.)
ANGPTL3
Abdominal subcutaneous adipose tissue volume (s.d.)
Beta: −0.0057
SE: 0.033
Pval: 1
Angiopoietin−like 3 protein (s.d.)
ANGPTL3
Liver iron content (s.d.)
Beta: −0.04
SE: 0.036
Pval: 1
Angiopoietin−like 3 protein (s.d.)
ANGPTL3
Pancreas fat (s.d.)
Beta: −0.024
SE: 0.04
Pval: 1
Angiopoietin−like 3 protein (s.d.)
ANGPTL3
Kidney volume (s.d.)
Beta: −0.039
SE: 0.029
Pval: 1
Angiopoietin−like 3 protein (s.d.)
ANGPTL3
Lung volume (s.d.)
Beta: −0.025
SE: 0.031
Pval: 1
Angiopoietin−like 3 protein (s.d.)
ANGPTL3
Pancreas volume (s.d.)
Beta: 0.033
SE: 0.035
Pval: 1
Angiopoietin−like 3 protein (s.d.)
ANGPTL3
Pancreas iron content (s.d.)
Beta: −0.029
SE: 0.04
Pval: 1
Angiopoietin−like 3 protein (s.d.)
ANGPTL3
Albumin (g/L)
Beta: 0.032
SE: 0.03
Pval: 1
Angiopoietin−like 3 protein (s.d.)
ANGPTL3
Alkaline phosphatase (U/L)
Beta: −0.021
SE: 0.29
Pval: 1
Angiopoietin−like 3 protein (s.d.)
ANGPTL3
Alanine aminotransferase (U/L)
Beta: 0.64
SE: 0.15
Pval: 0.0033
Angiopoietin−like 3 protein (s.d.)
ANGPTL3
Direct bilirubin (umol/L)
Beta: −0.00042
SE: 0.01
Pval: 1
Angiopoietin−like 3 protein (s.d.)
ANGPTL3
Urea (mmol/L)
Beta: 0.0064
SE: 0.015
Pval: 1
Angiopoietin−like 3 protein (s.d.)
ANGPTL3
Calcium (mmol/L)
Beta: 0.00077
SE: 0.0011
Pval: 1
Angiopoietin−like 3 protein (s.d.)
ANGPTL3
C−reactive protein (mg/L)
Beta: 0.11
SE: 0.049
Pval: 1
Angiopoietin−like 3 protein (s.d.)
ANGPTL3
Gamma glutamyltransferase (U/L)
Beta: 0.79
SE: 0.47
Pval: 1
Angiopoietin−like 3 protein (s.d.)
ANGPTL3
IGF−1 (nmol/L)
Beta: 0.03
SE: 0.062
Pval: 1
Angiopoietin−like 3 protein (s.d.)
ANGPTL3
Oestradiol (pmol/L)
Beta: 0.64
SE: 12
Pval: 1
Angiopoietin−like 3 protein (s.d.)
ANGPTL3
Phosphate (mmol/L)
Beta: −0.0013
SE: 0.0018
Pval: 1
Angiopoietin−like 3 protein (s.d.)
ANGPTL3
Rheumatoid factor (IU/ml)
Beta: −0.53
SE: 0.75
Pval: 1
Angiopoietin−like 3 protein (s.d.)
ANGPTL3
SHBG (nmol/L)
Beta: −0.27
SE: 0.3
Pval: 1
Angiopoietin−like 3 protein (s.d.)
ANGPTL3
Total bilirubin (umol/L)
Beta: 0.27
SE: 0.048
Pval: 1.9e−06
Angiopoietin−like 3 protein (s.d.)
ANGPTL3
Testosterone (nmol/L)
Beta: −0.1
SE: 0.032
Pval: 0.12
Angiopoietin−like 3 protein (s.d.)
ANGPTL3
Total protein (g/L)
Beta: 0.053
SE: 0.047
Pval: 1
Angiopoietin−like 3 protein (s.d.)
ANGPTL3
Urate (umol/L)
Beta: 0.5
SE: 0.77
Pval: 1
Angiopoietin−like 3 protein (s.d.)
ANGPTL3
Vitamin D (nmol/L)
Beta: −2.1
SE: 0.24
Pval: 1.2e−16
Angiopoietin−like 3 protein (s.d.)
ANGPTL3
Estimated sample dilution factor (factor)
Beta: −3.4e−05
SE: 0.00012
Pval: 1
Angiopoietin−like 3 protein (s.d.)
ANGPTL3
White blood cell (leukocyte) count (10^9 cells/L)
Beta: 0.044
SE: 0.023
Pval: 1
Angiopoietin−like 3 protein (s.d.)

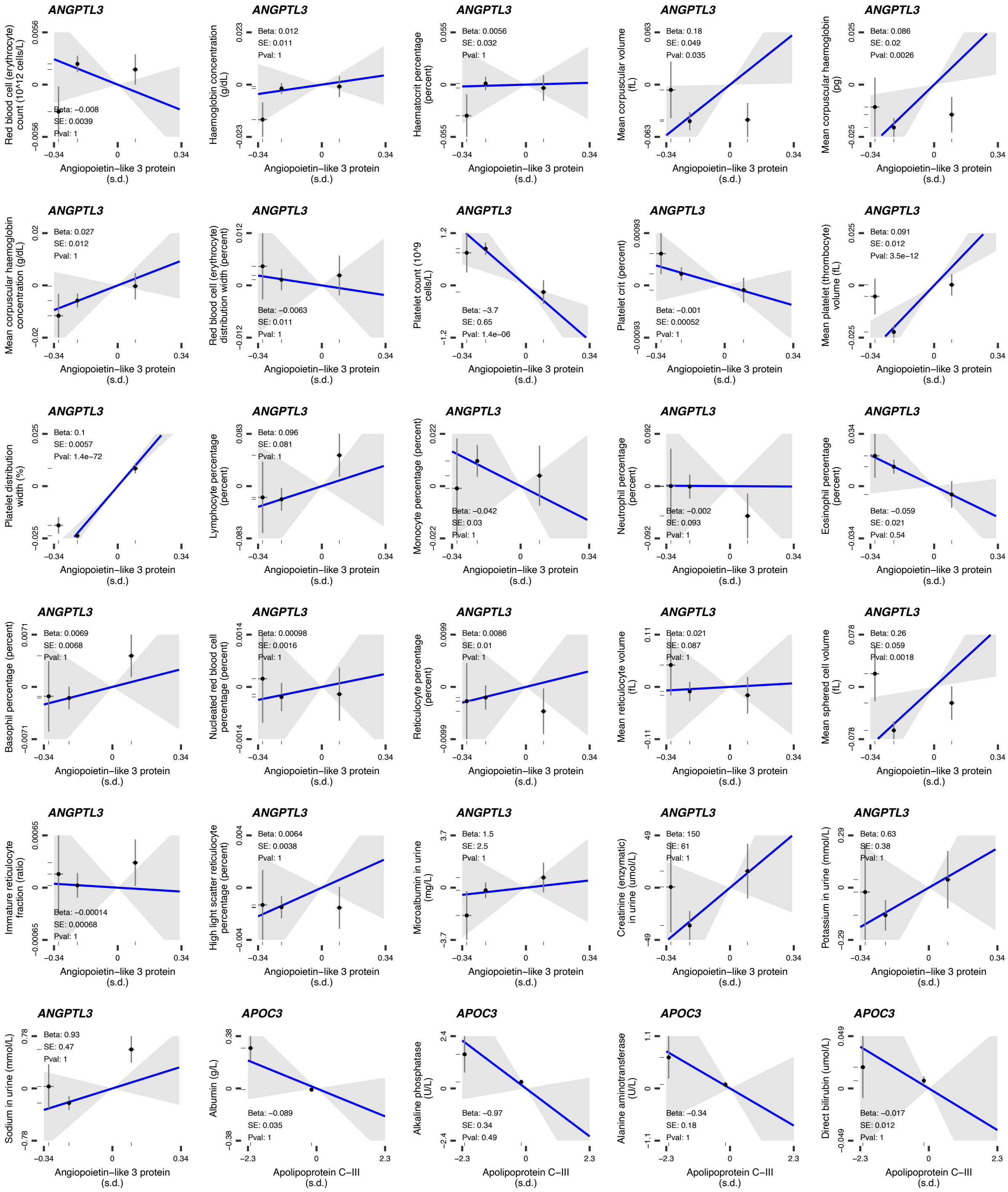
ANGPTL3
Red blood cell (erythrocyte) count (10^12 cells/L)
Beta: −0.008
SE: 0.0039
Pval: 1
Angiopoietin−like 3 protein (s.d.)
ANGPTL3
Haemoglobin concentration (g/dL)
Beta: 0.012
SE: 0.011
Pval: 1
Angiopoietin−like 3 protein (s.d.)
ANGPTL3
Haematocrit percentage (percent)
Beta: 0.0056
SE: 0.032
Pval: 1
Angiopoietin−like 3 protein (s.d.)
ANGPTL3
Mean corpuscular volume (fL)
Beta: 0.18
SE: 0.049
Pval: 0.035
Angiopoietin−like 3 protein (s.d.)
ANGPTL3
Mean corpuscular haemoglobin (pg)
Beta: 0.086
SE: 0.02
Pval: 0.0026
Angiopoietin−like 3 protein (s.d.)
ANGPTL3
Mean corpuscular haemoglobin concentration (g/dL)
Beta: 0.027
SE: 0.012
Pval: 1
Angiopoietin−like 3 protein (s.d.)
ANGPTL3
Red blood cell (erythrocyte) distribution width (percent)
Beta: −0.0063
SE: 0.011
Pval: 1
Angiopoietin−like 3 protein (s.d.)
ANGPTL3
Platelet count (10^9 cells/L)
Beta: −3.7
SE: 0.65
Pval: 1.4e−06
Angiopoietin−like 3 protein (s.d.)
ANGPTL3
Platelet crit (percent)
Beta: −0.001
SE: 0.00052
Pval: 1
Angiopoietin−like 3 protein (s.d.)
ANGPTL3
Mean platelet (thrombocyte) volume (fL)
Beta: 0.091
SE: 0.012
Pval: 3.5e−12
Angiopoietin−like 3 protein (s.d.)
ANGPTL3
Platelet distribution width (%)
Beta: 0.1
SE: 0.0057
Pval: 1.4e−72
Angiopoietin−like 3 protein (s.d.)
ANGPTL3
Lymphocyte percentage (percent)
Beta: 0.096
SE: 0.081
Pval: 1
Angiopoietin−like 3 protein (s.d.)
ANGPTL3
Monocyte percentage (percent)
Beta: −0.042
SE: 0.03
Pval: 1
Angiopoietin−like 3 protein (s.d.)
ANGPTL3
Neutrophil percentage (percent)
Beta: −0.002
SE: 0.093
Pval: 1
Angiopoietin−like 3 protein (s.d.)
ANGPTL3
Eosinophil percentage (percent)
Beta: −0.059
SE: 0.021
Pval: 0.54
Angiopoietin−like 3 protein (s.d.)
ANGPTL3
Basophil percentage (percent)
Beta: 0.0069
SE: 0.0068
Pval: 1
Angiopoietin−like 3 protein (s.d.)
ANGPTL3
Nucleated red blood cell percentage (percent)
Beta: 0.00098
SE: 0.0016
Pval: 1
Angiopoietin−like 3 protein (s.d.)
ANGPTL3
Reticulocyte percentage (percent)
Beta: 0.0086
SE: 0.01
Pval: 1
Angiopoietin−like 3 protein (s.d.)
ANGPTL3
Mean reticulocyte volume (fL)
Beta: 0.021
SE: 0.087
Pval: 1
Angiopoietin−like 3 protein (s.d.)
ANGPTL3
Mean sphered cell volume (fL)
Beta: 0.26
SE: 0.059
Pval: 0.0018
Angiopoietin−like 3 protein (s.d.)
ANGPTL3
Immature reticulocyte fraction (ratio)
Beta: −0.00014
SE: 0.00068
Pval: 1
Angiopoietin−like 3 protein (s.d.)
ANGPTL3
High light scatter reticulocyte percentage (percent)
Beta: 0.0064
SE: 0.0038
Pval: 1
Angiopoietin−like 3 protein (s.d.)
ANGPTL3
Microalbumin in urine (mg/L)
Beta: 1.5
SE: 2.5
Pval: 1
Angiopoietin−like 3 protein (s.d.)
ANGPTL3
Creatinine (enzymatic) in urine (umol/L)
Beta: 150
SE: 61
Pval: 1
Angiopoietin−like 3 protein (s.d.)
ANGPTL3
Potassium in urine (mmol/L)
Beta: 0.63
SE: 0.38
Pval: 1
Angiopoietin−like 3 protein (s.d.)
ANGPTL3
Sodium in urine (mmol/L)
Beta: 0.93
SE: 0.47
Pval: 1
Angiopoietin−like 3 protein (s.d.)
APOC3
Albumin (g/L)
Beta: −0.089
SE: 0.035
Pval: 1
Apolipoprotein C−III (s.d.)
APOC3
Alkaline phosphatase (U/L)
Beta: −0.97
SE: 0.34
Pval: 0.49
Apolipoprotein C−III (s.d.)
APOC3
Alanine aminotransferase (U/L)
Beta: −0.34
SE: 0.18
Pval: 1
Apolipoprotein C−III (s.d.)
APOC3
Direct bilirubin (umol/L)
Beta: −0.017
SE: 0.012
Pval: 1
Apolipoprotein C−III (s.d.)

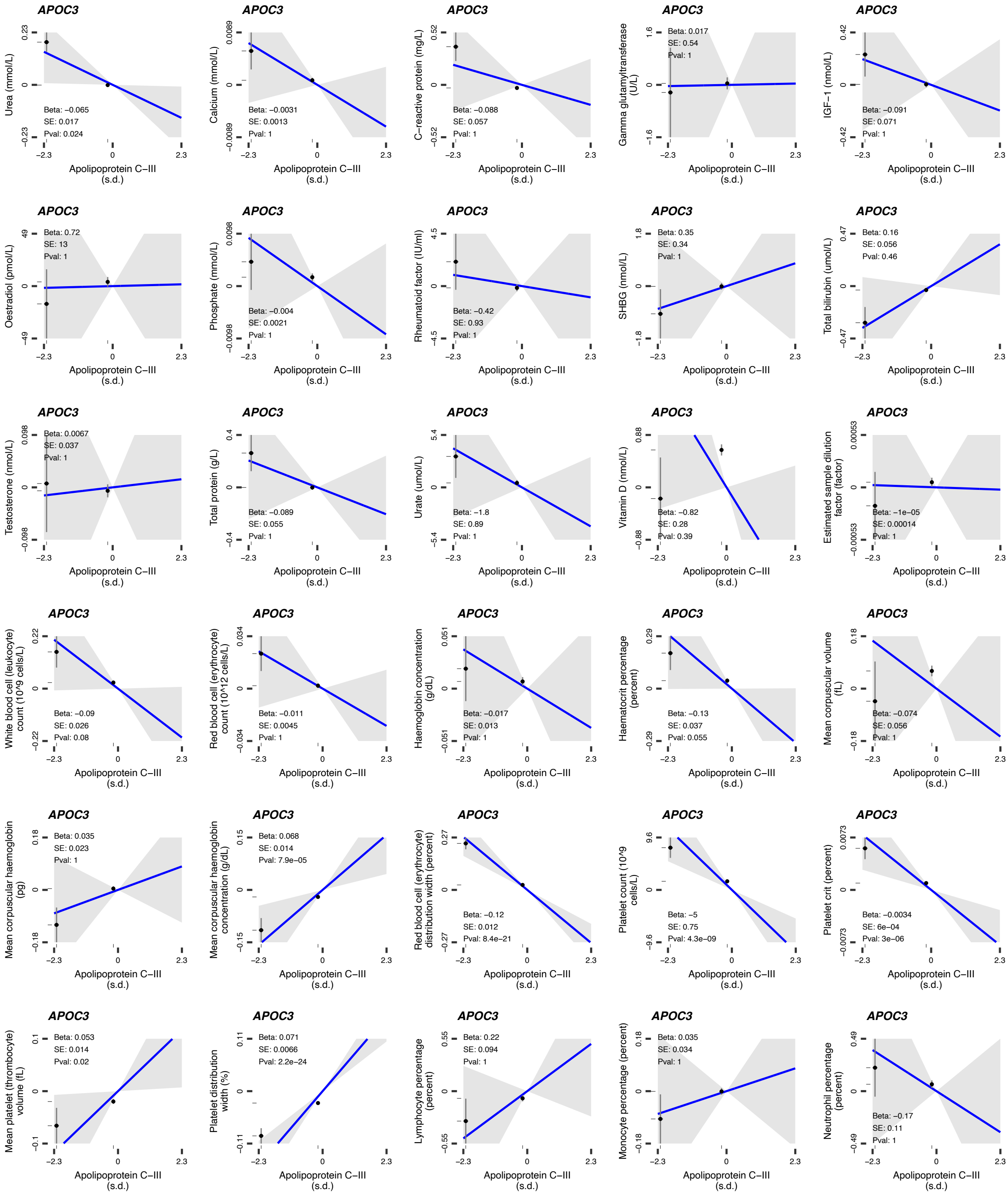
APOC3
Urea (mmol/L)
Beta: −0.065
SE: 0.017
Pval: 0.024
Apolipoprotein C−III (s.d.)
APOC3
Calcium (mmol/L)
Beta: −0.0031
SE: 0.0013
Pval: 1
Apolipoprotein C−III (s.d.)
APOC3
C−reactive protein (mg/L)
Beta: −0.088
SE: 0.057
Pval: 1
Apolipoprotein C−III (s.d.)
APOC3
Gamma glutamyltransferase (U/L)
Beta: 0.017
SE: 0.54
Pval: 1
Apolipoprotein C−III (s.d.)
APOC3
IGF−1 (nmol/L)
Beta: −0.091
SE: 0.071
Pval: 1
Apolipoprotein C−III (s.d.)
APOC3
Oestradiol (pmol/L)
Beta: 0.72
SE: 13
Pval: 1
Apolipoprotein C−III (s.d.)
APOC3
Phosphate (mmol/L)
Beta: −0.004
SE: 0.0021
Pval: 1
Apolipoprotein C−III (s.d.)
APOC3
Rheumatoid factor (IU/ml)
Beta: −0.42
SE: 0.93
Pval: 1
Apolipoprotein C−III (s.d.)
APOC3
SHBG (nmol/L)
Beta: 0.35
SE: 0.34
Pval: 1
Apolipoprotein C−III (s.d.)
APOC3
Total bilirubin (umol/L)
Beta: 0.16
SE: 0.056
Pval: 0.46
Apolipoprotein C−III (s.d.)
APOC3
Testosterone (nmol/L)
Beta: 0.0067
SE: 0.037
Pval: 1
Apolipoprotein C−III (s.d.)
APOC3
Total protein (g/L)
Beta: −0.089
SE: 0.055
Pval: 1
Apolipoprotein C−III (s.d.)
APOC3
Urate (umol/L)
Beta: −1.8
SE: 0.89
Pval: 1
Apolipoprotein C−III (s.d.)
APOC3
Vitamin D (nmol/L)
Beta: −0.82
SE: 0.28
Pval: 0.39
Apolipoprotein C−III (s.d.)
APOC3
Estimated sample dilution factor (factor)
Beta: −1e−05
SE: 0.00014
Pval: 1
Apolipoprotein C−III (s.d.)
APOC3
White blood cell (leukocyte) count (10^9 cells/L)
Beta: −0.09
SE: 0.026
Pval: 0.08
Apolipoprotein C−III (s.d.)
APOC3
Red blood cell (erythrocyte) count (10^12 cells/L)
Beta: −0.011
SE: 0.0045
Pval: 1
Apolipoprotein C−III (s.d.)
APOC3
Haemoglobin concentration (g/dL)
Beta: −0.017
SE: 0.013
Pval: 1
Apolipoprotein C−III (s.d.)
APOC3
Haematocrit percentage (percent)
Beta: −0.13
SE: 0.037
Pval: 0.055
Apolipoprotein C−III (s.d.)
APOC3
Mean corpuscular volume (fL)
Beta: −0.074
SE: 0.056
Pval: 1
Apolipoprotein C−III (s.d.)
APOC3
Mean corpuscular haemoglobin (pg)
Beta: 0.035
SE: 0.023
Pval: 1
Apolipoprotein C−III (s.d.)
APOC3
Mean corpuscular haemoglobin concentration (g/dL)
Beta: 0.068
SE: 0.014
Pval: 7.9e−05
Apolipoprotein C−III (s.d.)
APOC3
Red blood cell (erythrocyte) distribution width (percent)
Beta: −0.12
SE: 0.012
Pval: 8.4e−21
Apolipoprotein C−III (s.d.)
APOC3
Platelet count (10^9 cells/L)
Beta: −5
SE: 0.75
Pval: 4.3e−09
Apolipoprotein C−III (s.d.)
APOC3
Platelet crit (percent)
Beta: −0.0034
SE: 6e−04
Pval: 3e−06
Apolipoprotein C−III (s.d.)
APOC3
Mean platelet (thrombocyte) volume (fL)
Beta: 0.053
SE: 0.014
Pval: 0.02
Apolipoprotein C−III (s.d.)
APOC3
Platelet distribution width (%)
Beta: 0.071
SE: 0.0066
Pval: 2.2e−24
Apolipoprotein C−III (s.d.)
APOC3
Lymphocyte percentage (percent)
Beta: 0.22
SE: 0.094
Pval: 1
Apolipoprotein C−III (s.d.)
APOC3
Monocyte percentage (percent)
Beta: 0.035
SE: 0.034
Pval: 1
Apolipoprotein C−III (s.d.)
APOC3
Neutrophil percentage (percent)
Beta: −0.17
SE: 0.11
Pval: 1
Apolipoprotein C−III (s.d.)

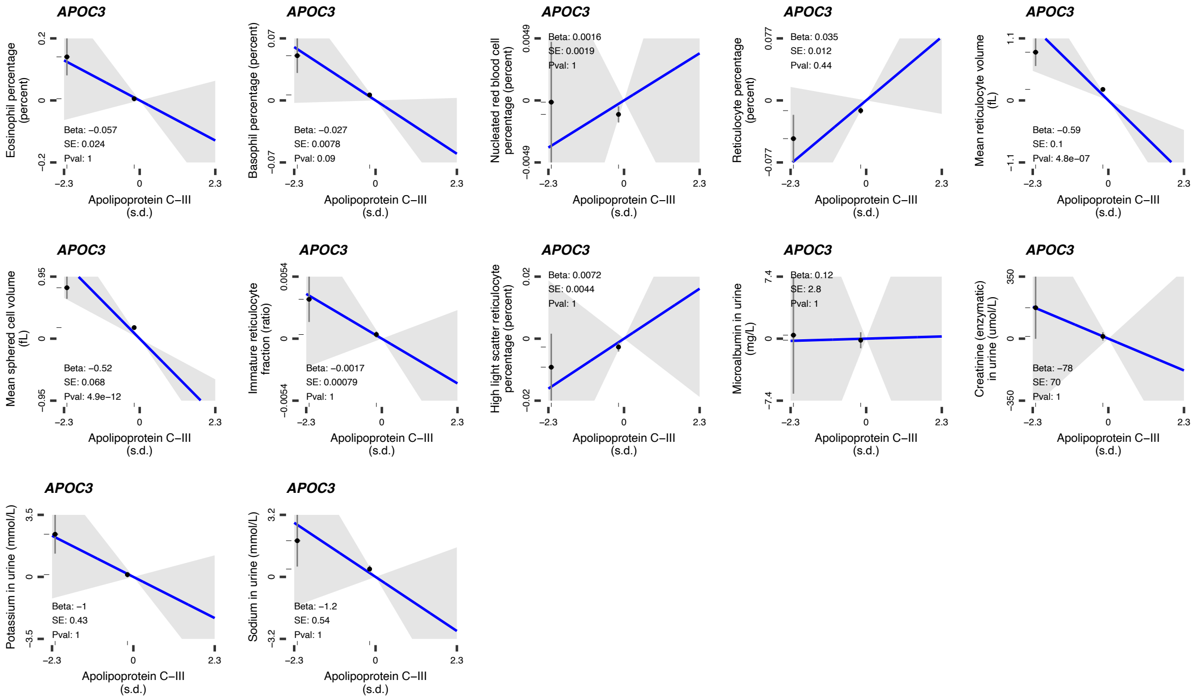
APOC3
Eosinophil percentage (percent)
Beta: −0.057
SE: 0.024
Pval: 1
APOC3
Basophil percentage (percent)
Beta: −0.027
SE: 0.0078
Pval: 0.09
APOC3
Nucleated red blood cell percentage (percent)
Beta: 0.0016
SE: 0.0019
Pval: 1
APOC3
Reticulocyte percentage (percent)
Beta: 0.035
SE: 0.012
Pval: 0.44
APOC3
Mean reticulocyte volume (fL)
Beta: −0.59
SE: 0.1
Pval: 4.8e−07
APOC3
Mean sphered cell volume (fL)
Beta: −0.52
SE: 0.068
Pval: 4.9e−12
APOC3
Immature reticulocyte fraction (ratio)
Beta: −0.0017
SE: 0.00079
Pval: 1
APOC3
High light scatter reticulocyte percentage (percent)
Beta: 0.0072
SE: 0.0044
Pval: 1
APOC3
Microalbumin in urine (mg/L)
Beta: 0.12
SE: 2.8
Pval: 1
APOC3
Creatinine (enzymatic) in urine (umol/L)
Beta: −78
SE: 70
Pval: 1
APOC3
Potassium in urine (mmol/L)
Beta: −1
SE: 0.43
Pval: 1
APOC3
Sodium in urine (mmol/L)
Beta: −1.2
SE: 0.54
Pval: 1
Apolipoprotein C−III (s.d.)

### SUPPLEMENTAL FIGURE 2

**Figure S2. Regional genetic association plots showing the results of the colocalization analyses.** Each subplot represents the results of the analyses displayed in **Figure 2-4** of the main manuscript.

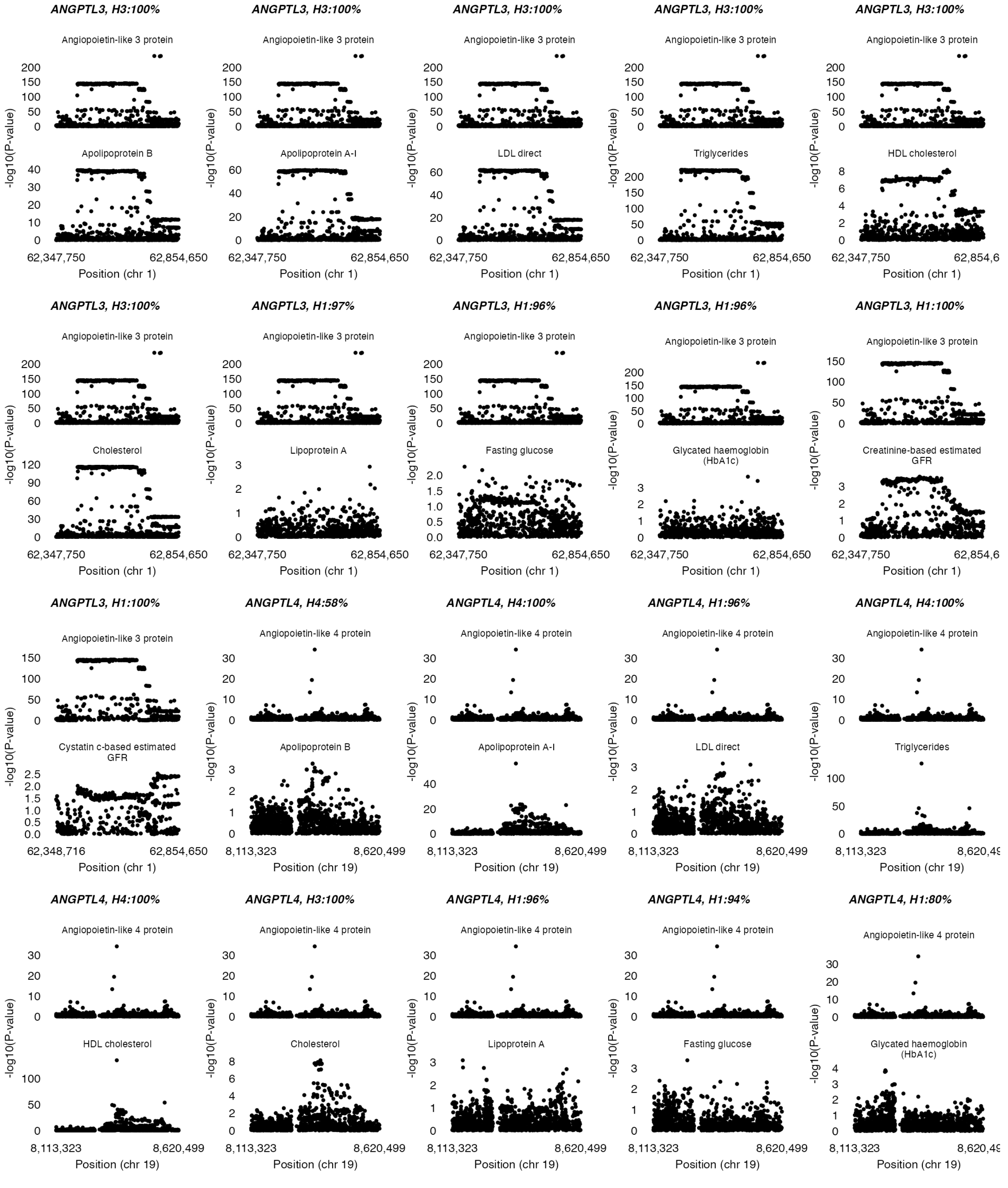

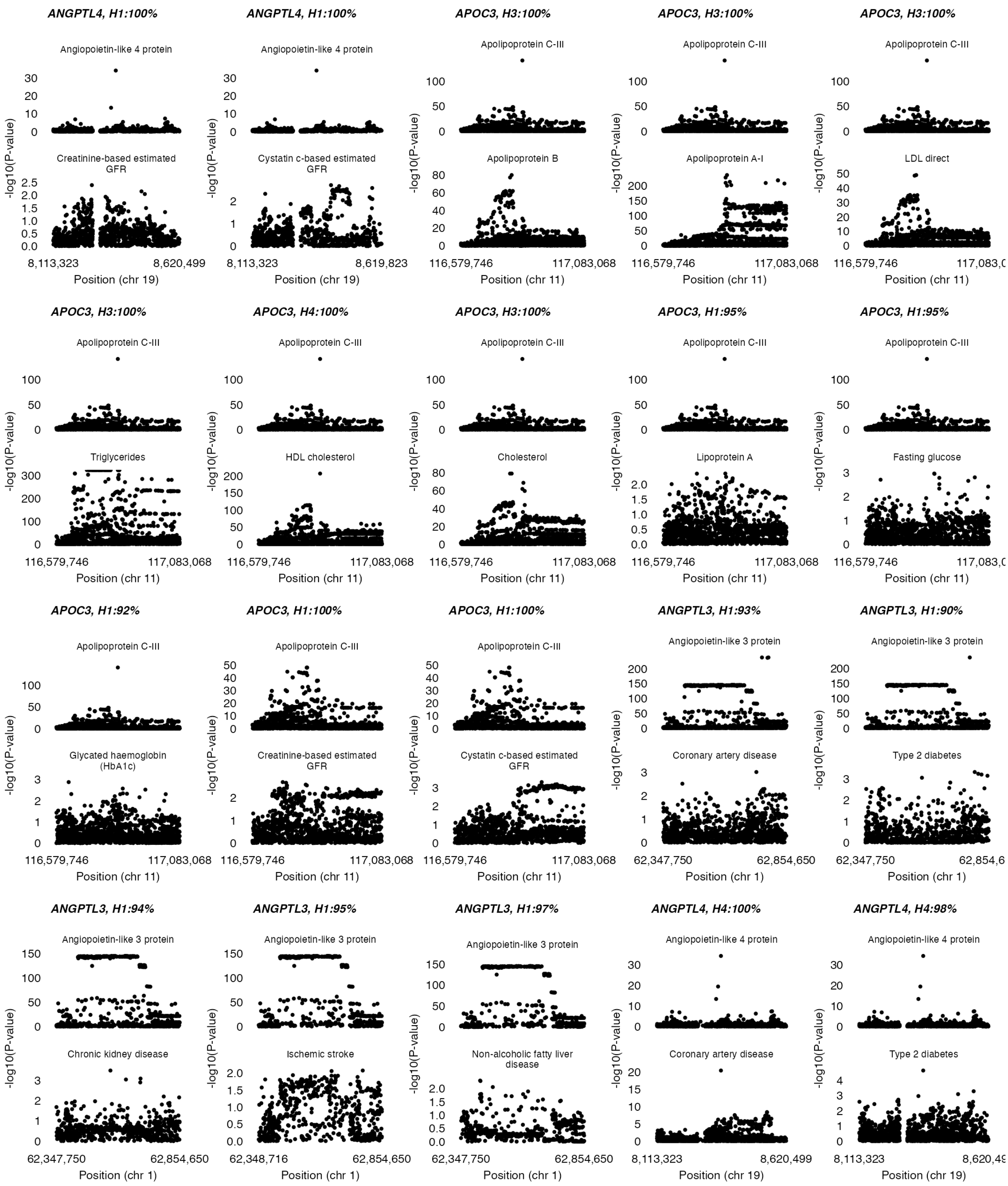
ANGPTL4, H1:100%
Angiopoietin-like 4 protein
Creatinine-based estimated GFR
8,113,323
8,620,499
Position (chr 19)
ANGPTL4, H1:100%
Angiopoietin-like 4 protein
Cystatin c-based estimated GFR
8,113,323
8,619,823
Position (chr 19)
APOC3, H3:100%
Apolipoprotein C-III
Apolipoprotein B
116,579,746
117,083,068
Position (chr 11)
APOC3, H3:100%
Apolipoprotein C-III
Apolipoprotein A-I
APOC3, H3:100%
Apolipoprotein C-III
LDL direct
APOC3, H3:100%
Triglycerides
APOC3, H4:100%
HDL cholesterol
APOC3, H3:100%
Cholesterol
APOC3, H1:95%
Lipoprotein A
APOC3, H1:95%
Fasting glucose
APOC3, H1:92%
Glycated haemoglobin (HbA1c)
APOC3, H1:100%
Creatinine-based estimated GFR
APOC3, H1:100%
Cystatin c-based estimated GFR
ANGPTL3, H1:93%
Angiopoietin-like 3 protein
Coronary artery disease
62,347,750
62,854,650
Position (chr 1)
ANGPTL3, H1:90%
Type 2 diabetes
ANGPTL3, H1:94%
Chronic kidney disease
ANGPTL3, H1:95%
Ischemic stroke
62,348,716
ANGPTL3, H1:97%
Non-alcoholic fatty liver disease
ANGPTL4, H4:100%
Coronary artery disease
ANGPTL4, H4:98%
Type 2 diabetes
-log10(P-value)

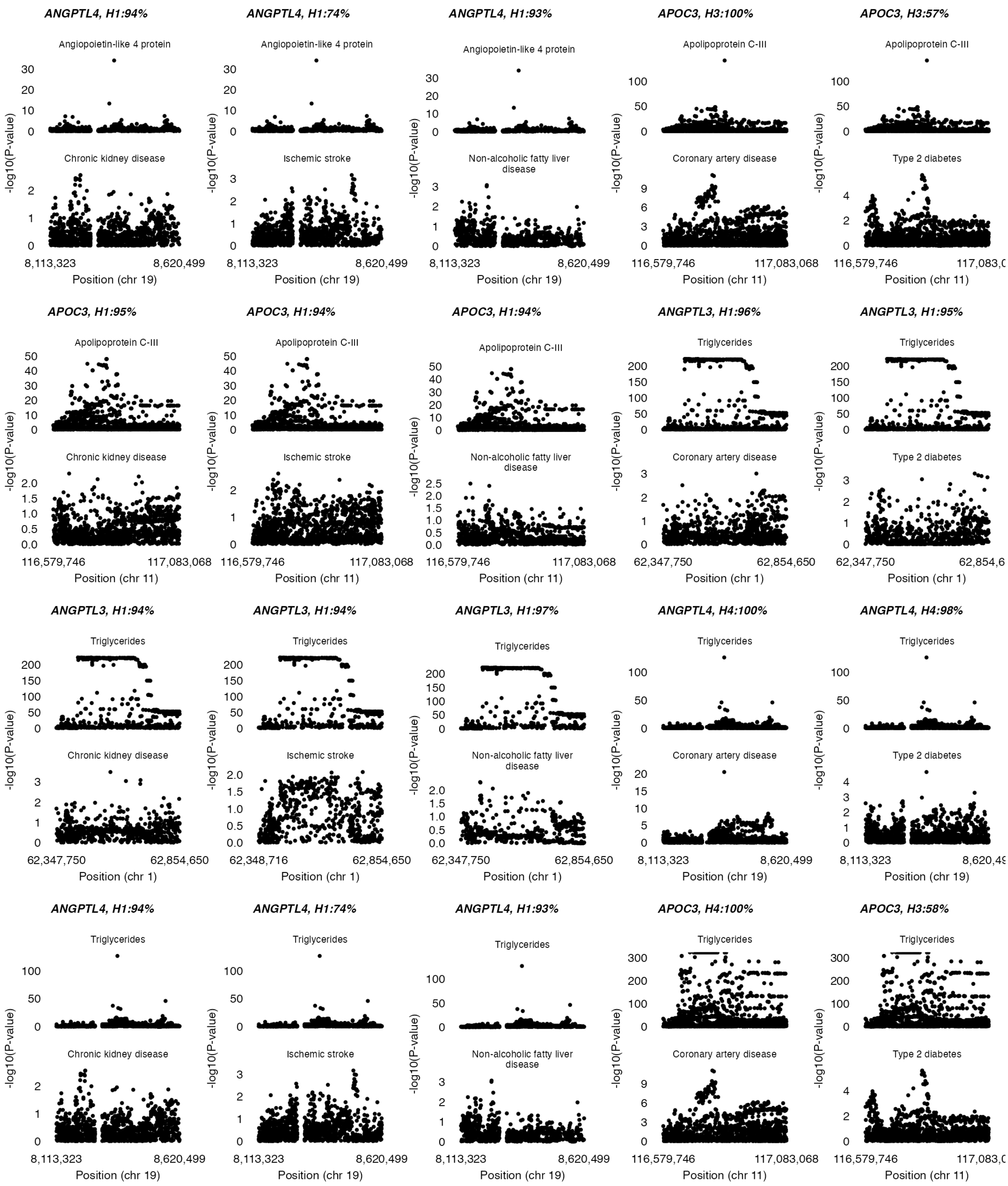
ANGPTL4, H1:94%
ANGPTL4, H1:74%
ANGPTL4, H1:93%
APOC3, H3:100%
APOC3, H3:57%
Angiopoietin-like 4 protein
Apolipoprotein C-III
Chronic kidney disease
Ischemic stroke
Non-alcoholic fatty liver disease
Coronary artery disease
Type 2 diabetes
-log10(P-value)
8,113,323
8,620,499
Position (chr 19)
116,579,746
117,083,068
Position (chr 11)
APOC3, H1:95%
APOC3, H1:94%
APOC3, H1:94%
ANGPTL3, H1:96%
ANGPTL3, H1:95%
Triglycerides
62,347,750
62,854,650
Position (chr 1)
ANGPTL3, H1:94%
ANGPTL3, H1:94%
ANGPTL3, H1:97%
ANGPTL4, H4:100%
ANGPTL4, H4:98%
62,348,716
ANGPTL4, H1:94%
ANGPTL4, H1:74%
ANGPTL4, H1:93%
APOC3, H4:100%
APOC3, H3:58%

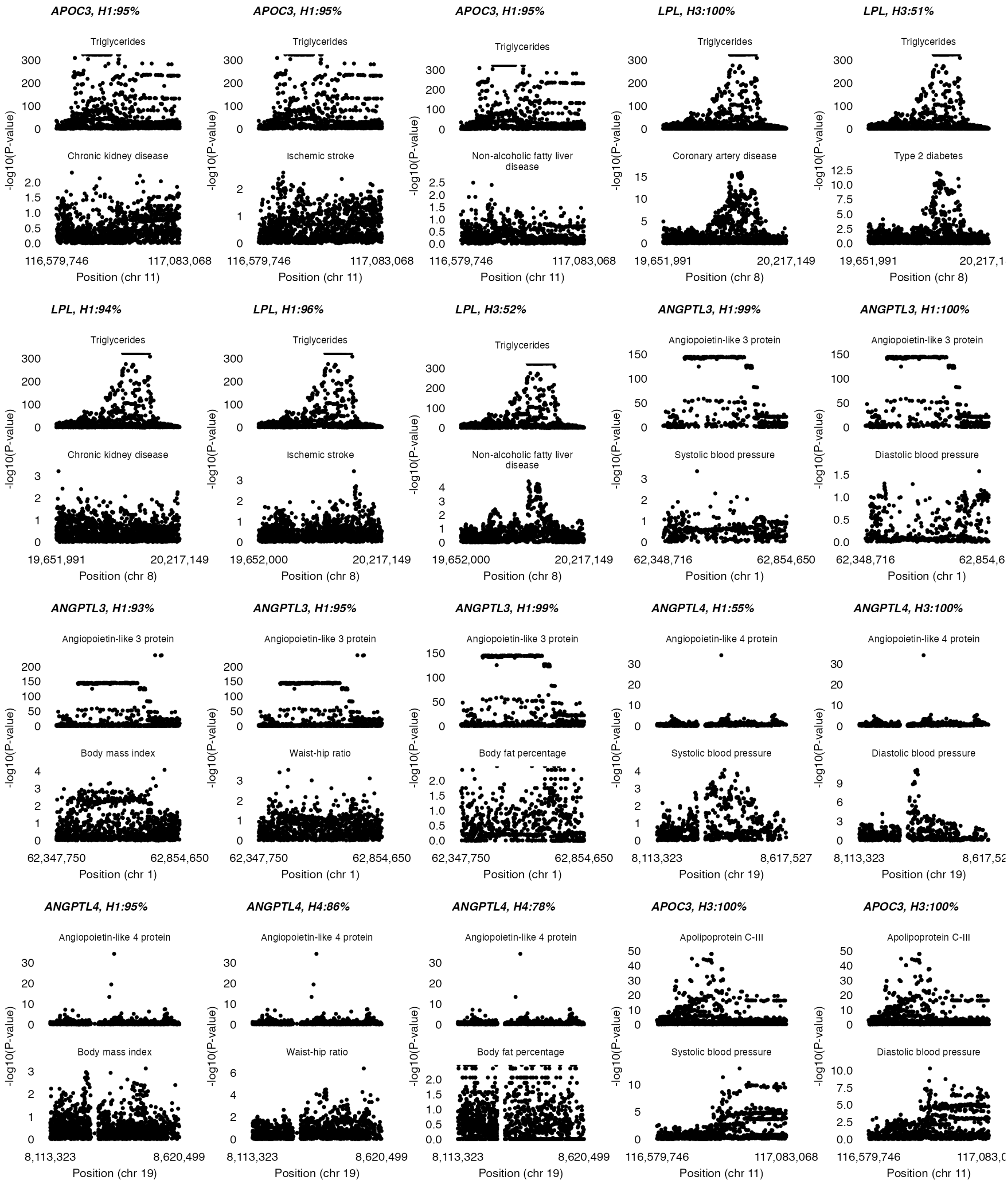
APOC3, H1:95%
Triglycerides
Chronic kidney disease
116,579,746
117,083,068
Position (chr 11)
APOC3, H1:95%
Triglycerides
Ischemic stroke
116,579,746
117,083,068
Position (chr 11)
APOC3, H1:95%
Triglycerides
Non-alcoholic fatty liver disease
116,579,746
117,083,068
Position (chr 11)
LPL, H3:100%
Triglycerides
Coronary artery disease
19,651,991
20,217,149
Position (chr 8)
LPL, H3:51%
Triglycerides
Type 2 diabetes
19,651,991
Position (chr 8)
LPL, H1:94%
Triglycerides
Chronic kidney disease
19,651,991
20,217,149
Position (chr 8)
LPL, H1:96%
Triglycerides
Ischemic stroke
19,652,000
20,217,149
Position (chr 8)
LPL, H3:52%
Triglycerides
Non-alcoholic fatty liver disease
19,652,000
20,217,149
Position (chr 8)
ANGPTL3, H1:99%
Angiopoietin-like 3 protein
Systolic blood pressure
62,348,716
62,854,650
Position (chr 1)
ANGPTL3, H1:100%
Angiopoietin-like 3 protein
Diastolic blood pressure
62,348,716
Position (chr 1)
ANGPTL3, H1:93%
Angiopoietin-like 3 protein
Body mass index
62,347,750
62,854,650
Position (chr 1)
ANGPTL3, H1:95%
Angiopoietin-like 3 protein
Waist-hip ratio
62,347,750
62,854,650
Position (chr 1)
ANGPTL3, H1:99%
Angiopoietin-like 3 protein
Body fat percentage
62,347,750
62,854,650
Position (chr 1)
ANGPTL4, H1:55%
Angiopoietin-like 4 protein
Systolic blood pressure
8,113,323
8,617,527
Position (chr 19)
ANGPTL4, H3:100%
Angiopoietin-like 4 protein
Diastolic blood pressure
8,113,323
Position (chr 19)
ANGPTL4, H1:95%
Angiopoietin-like 4 protein
Body mass index
8,113,323
8,620,499
Position (chr 19)
ANGPTL4, H4:86%
Angiopoietin-like 4 protein
Waist-hip ratio
8,113,323
8,620,499
Position (chr 19)
ANGPTL4, H4:78%
Angiopoietin-like 4 protein
Body fat percentage
8,113,323
8,620,499
Position (chr 19)
APOC3, H3:100%
Apolipoprotein C-III
Systolic blood pressure
116,579,746
117,083,068
Position (chr 11)
APOC3, H3:100%
Apolipoprotein C-III
Diastolic blood pressure
116,579,746
Position (chr 11)
-log10(P-value)

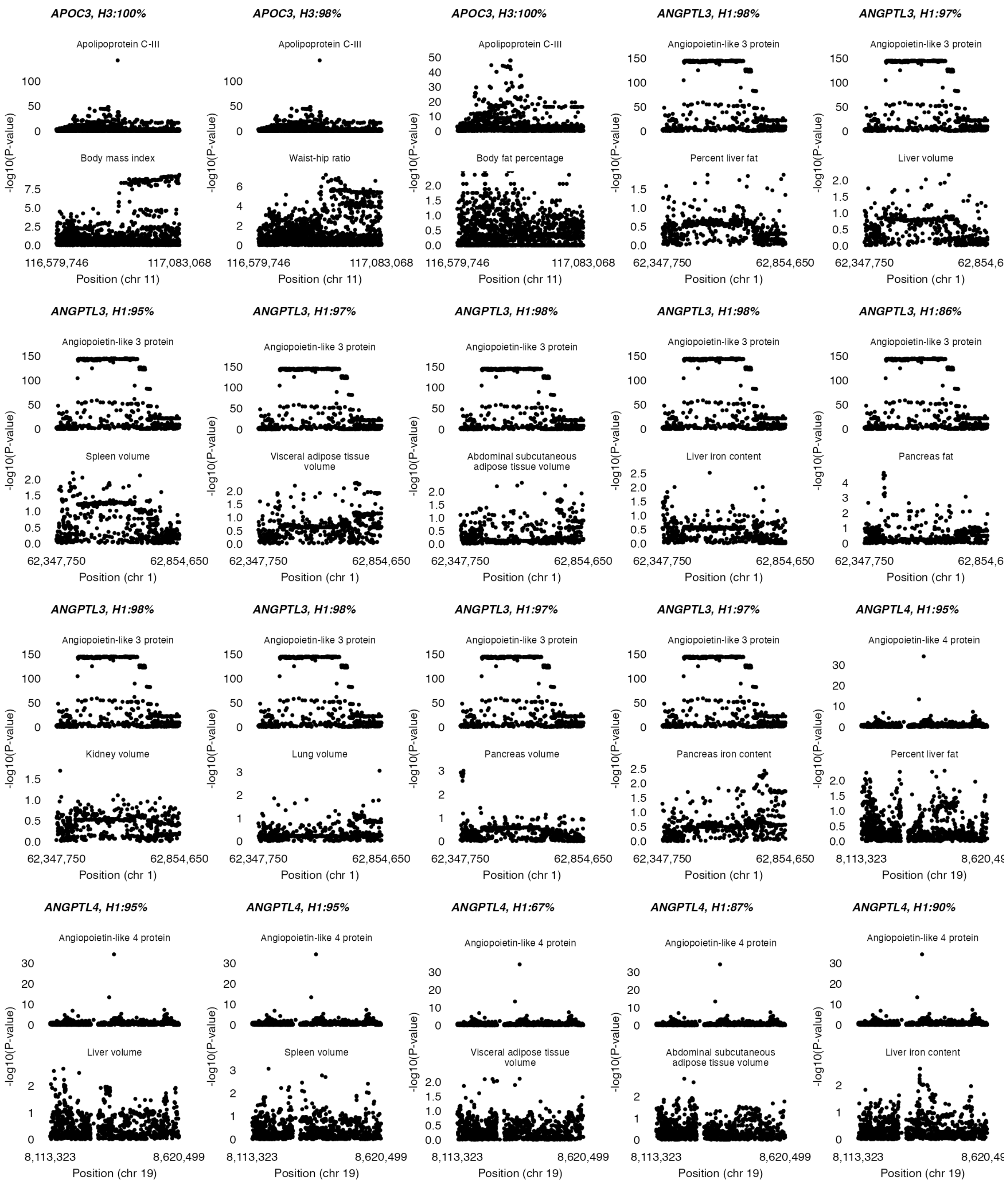
APOC3, H3:100%
Apolipoprotein C-III
Body mass index
APOC3, H3:98%
Apolipoprotein C-III
Waist-hip ratio
APOC3, H3:100%
Apolipoprotein C-III
Body fat percentage
Position (chr 11)
ANGPTL3, H1:98%
Angiopoietin-like 3 protein
Percent liver fat
ANGPTL3, H1:97%
Angiopoietin-like 3 protein
Liver volume
Position (chr 1)
ANGPTL3, H1:95%
Angiopoietin-like 3 protein
Spleen volume
ANGPTL3, H1:97%
Angiopoietin-like 3 protein
Visceral adipose tissue volume
ANGPTL3, H1:98%
Angiopoietin-like 3 protein
Abdominal subcutaneous adipose tissue volume
ANGPTL3, H1:98%
Angiopoietin-like 3 protein
Liver iron content
ANGPTL3, H1:86%
Angiopoietin-like 3 protein
Pancreas fat
ANGPTL3, H1:98%
Angiopoietin-like 3 protein
Kidney volume
ANGPTL3, H1:98%
Angiopoietin-like 3 protein
Lung volume
ANGPTL3, H1:97%
Angiopoietin-like 3 protein
Pancreas volume
ANGPTL3, H1:97%
Angiopoietin-like 3 protein
Pancreas iron content
ANGPTL4, H1:95%
Angiopoietin-like 4 protein
Percent liver fat
Position (chr 19)
ANGPTL4, H1:95%
Angiopoietin-like 4 protein
Liver volume
ANGPTL4, H1:95%
Angiopoietin-like 4 protein
Spleen volume
ANGPTL4, H1:67%
Angiopoietin-like 4 protein
Visceral adipose tissue volume
ANGPTL4, H1:87%
Angiopoietin-like 4 protein
Abdominal subcutaneous adipose tissue volume
ANGPTL4, H1:90%
Angiopoietin-like 4 protein
Liver iron content
-log10(P-value)

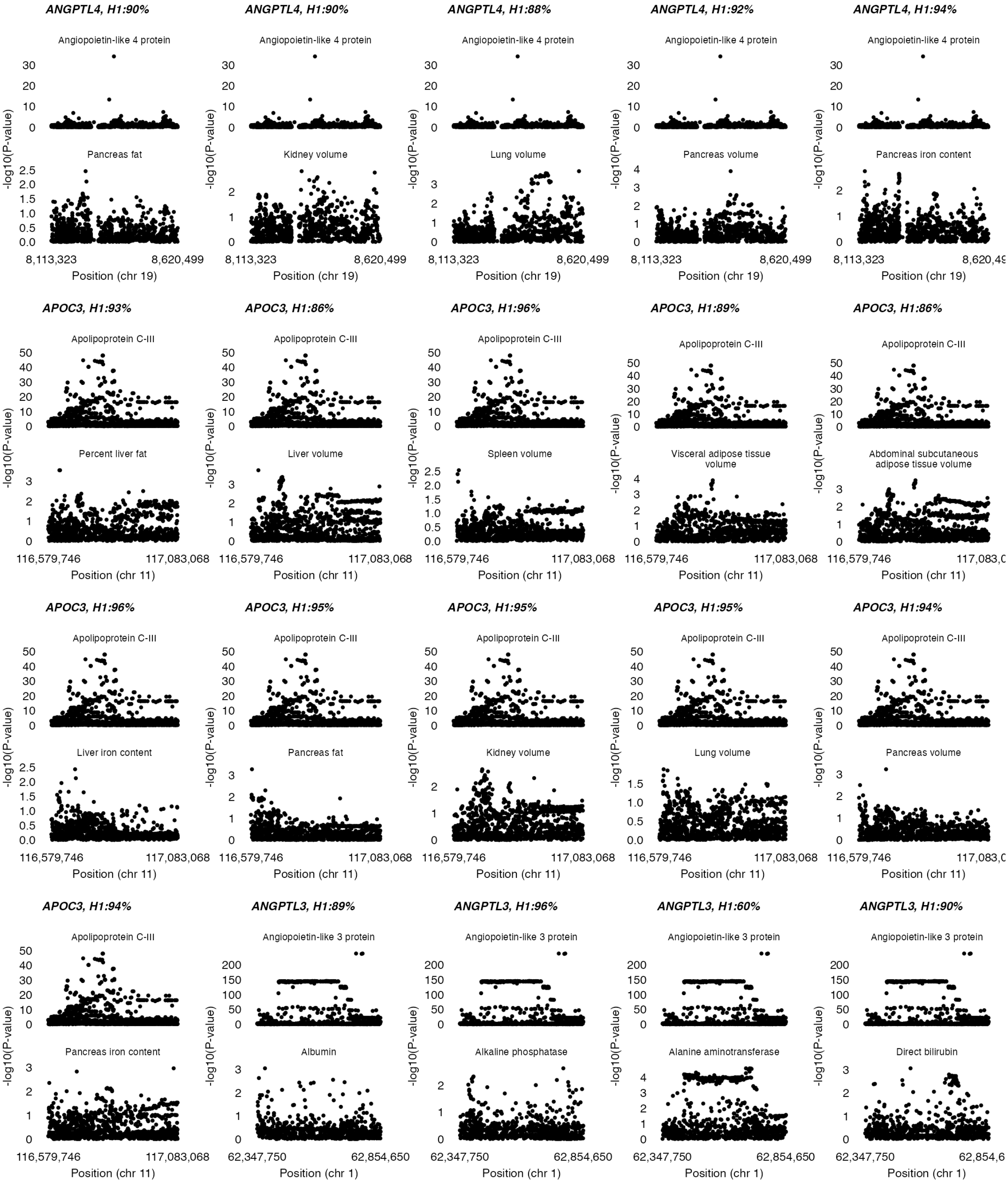
ANGPTL4, H1:90%
Angiopoietin-like 4 protein
Pancreas fat
ANGPTL4, H1:90%
Angiopoietin-like 4 protein
Kidney volume
ANGPTL4, H1:88%
Angiopoietin-like 4 protein
Lung volume
ANGPTL4, H1:92%
Angiopoietin-like 4 protein
Pancreas volume
ANGPTL4, H1:94%
Angiopoietin-like 4 protein
Pancreas iron content
-log10(P-value)
8,113,323
8,620,499
Position (chr 19)
APOC3, H1:93%
Apolipoprotein C-III
Percent liver fat
APOC3, H1:86%
Apolipoprotein C-III
Liver volume
APOC3, H1:96%
Apolipoprotein C-III
Spleen volume
APOC3, H1:89%
Apolipoprotein C-III
Visceral adipose tissue volume
APOC3, H1:86%
Apolipoprotein C-III
Abdominal subcutaneous adipose tissue volume
APOC3, H1:96%
Apolipoprotein C-III
Liver iron content
APOC3, H1:95%
Apolipoprotein C-III
Pancreas fat
APOC3, H1:95%
Apolipoprotein C-III
Kidney volume
APOC3, H1:95%
Apolipoprotein C-III
Lung volume
APOC3, H1:94%
Apolipoprotein C-III
Pancreas volume
116,579,746
117,083,068
Position (chr 11)
APOC3, H1:94%
Apolipoprotein C-III
Pancreas iron content
ANGPTL3, H1:89%
Angiopoietin-like 3 protein
Albumin
ANGPTL3, H1:96%
Angiopoietin-like 3 protein
Alkaline phosphatase
ANGPTL3, H1:60%
Angiopoietin-like 3 protein
Alanine aminotransferase
ANGPTL3, H1:90%
Angiopoietin-like 3 protein
Direct bilirubin
62,347,750
62,854,650
Position (chr 1)

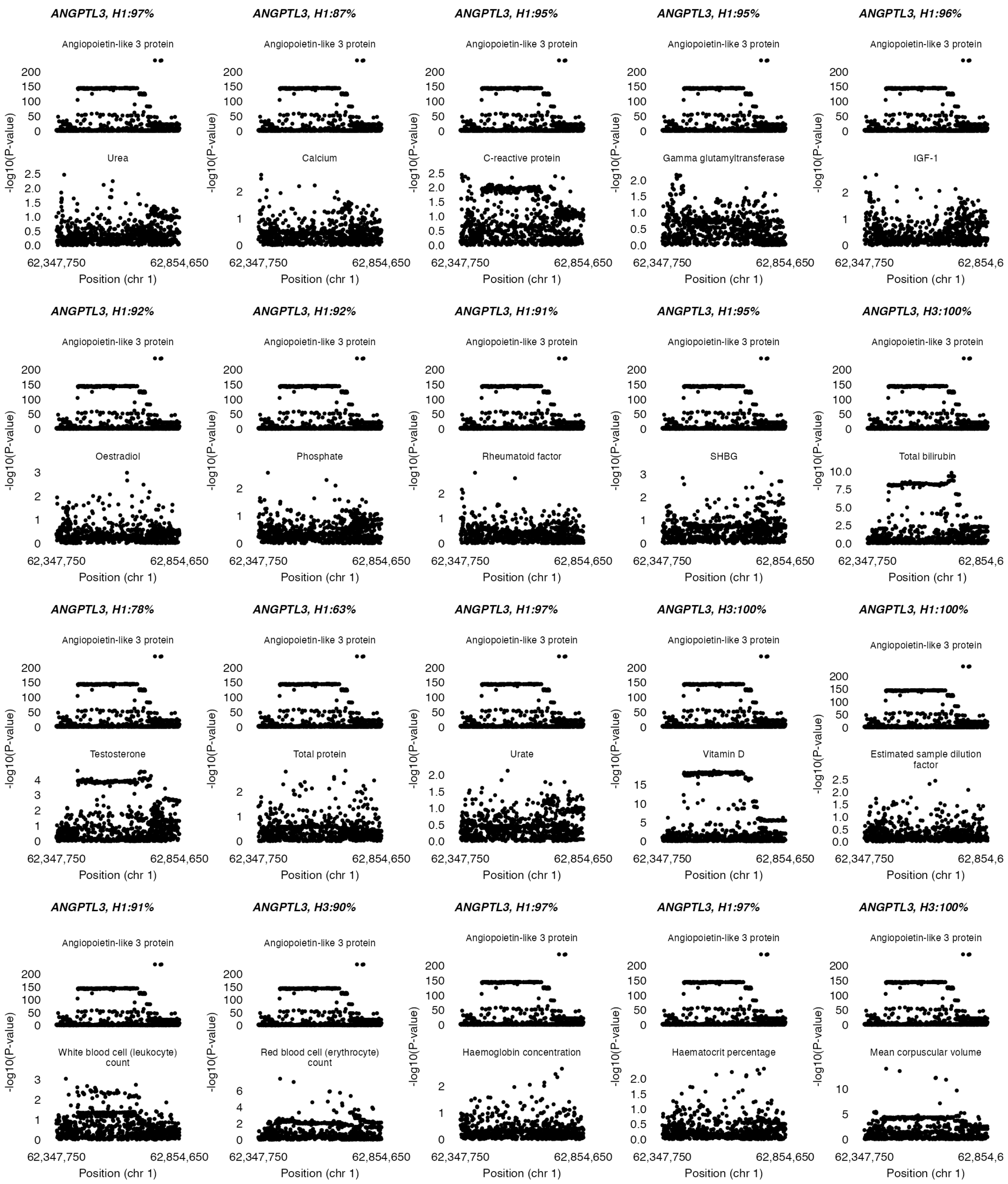
ANGPTL3, H1:97%
ANGPTL3, H1:87%
ANGPTL3, H1:95%
ANGPTL3, H1:95%
ANGPTL3, H1:96%
Angiopoietin-like 3 protein
Urea
Calcium
C-reactive protein
Gamma glutamyltransferase
IGF-1
ANGPTL3, H1:92%
ANGPTL3, H1:92%
ANGPTL3, H1:91%
ANGPTL3, H1:95%
ANGPTL3, H3:100%
Oestradiol
Phosphate
Rheumatoid factor
SHBG
Total bilirubin
ANGPTL3, H1:78%
ANGPTL3, H1:63%
ANGPTL3, H1:97%
ANGPTL3, H3:100%
ANGPTL3, H1:100%
Testosterone
Total protein
Urate
Vitamin D
Estimated sample dilution factor
ANGPTL3, H1:91%
ANGPTL3, H3:90%
ANGPTL3, H1:97%
ANGPTL3, H1:97%
ANGPTL3, H3:100%
White blood cell (leukocyte) count
Red blood cell (erythrocyte) count
Haemoglobin concentration
Haematocrit percentage
Mean corpuscular volume
-log10(P-value)
62,347,750
62,854,650
Position (chr 1)

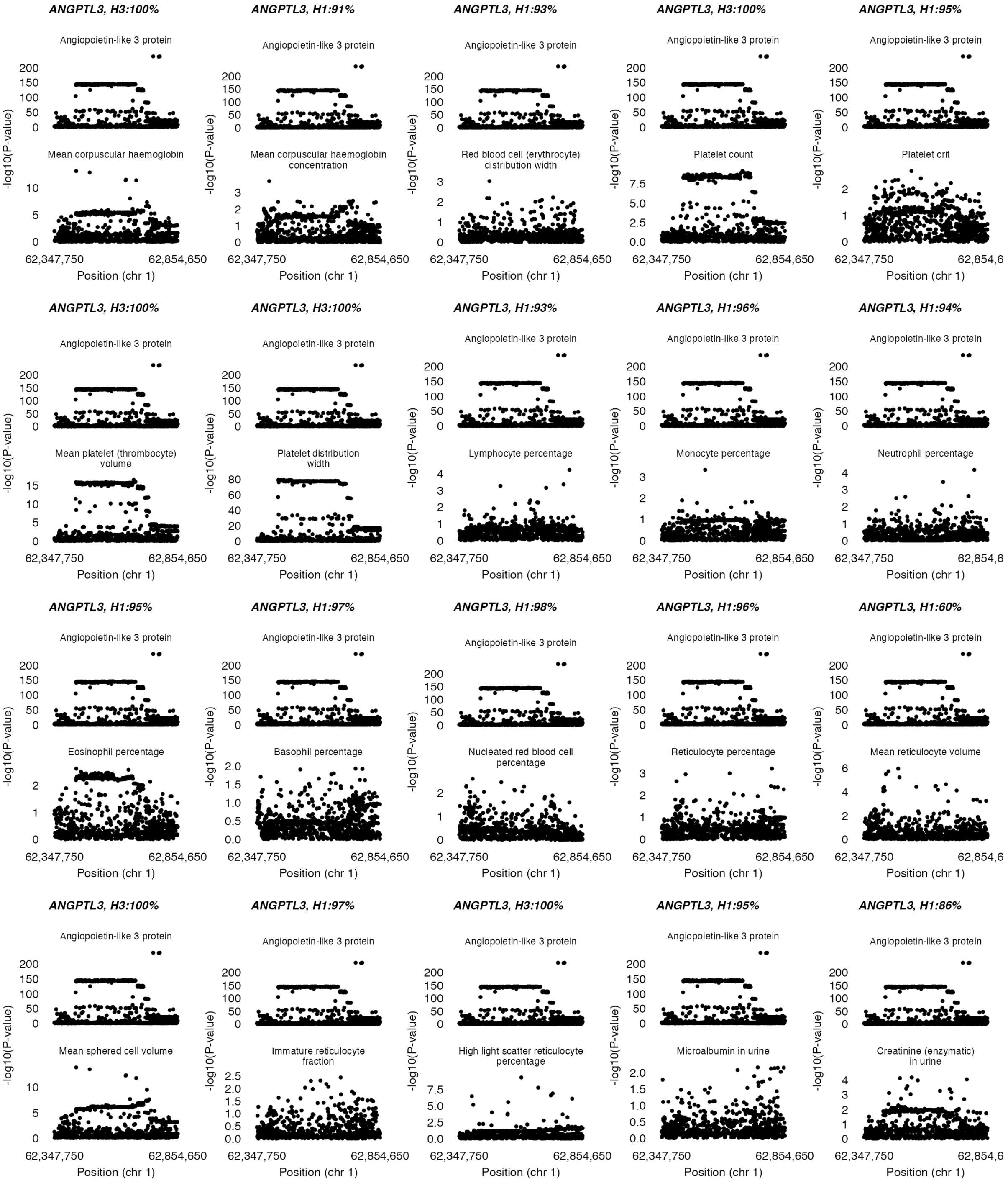
ANGPTL3, H3:100%
Angiopoietin-like 3 protein
Mean corpuscular haemoglobin
ANGPTL3, H1:91%
Angiopoietin-like 3 protein
Mean corpuscular haemoglobin concentration
ANGPTL3, H1:93%
Angiopoietin-like 3 protein
Red blood cell (erythrocyte) distribution width
ANGPTL3, H3:100%
Angiopoietin-like 3 protein
Platelet count
ANGPTL3, H1:95%
Angiopoietin-like 3 protein
Platelet crit
ANGPTL3, H3:100%
Angiopoietin-like 3 protein
Mean platelet (thrombocyte) volume
ANGPTL3, H3:100%
Angiopoietin-like 3 protein
Platelet distribution width
ANGPTL3, H1:93%
Angiopoietin-like 3 protein
Lymphocyte percentage
ANGPTL3, H1:96%
Angiopoietin-like 3 protein
Monocyte percentage
ANGPTL3, H1:94%
Angiopoietin-like 3 protein
Neutrophil percentage
ANGPTL3, H1:95%
Angiopoietin-like 3 protein
Eosinophil percentage
ANGPTL3, H1:97%
Angiopoietin-like 3 protein
Basophil percentage
ANGPTL3, H1:98%
Angiopoietin-like 3 protein
Nucleated red blood cell percentage
ANGPTL3, H1:96%
Angiopoietin-like 3 protein
Reticulocyte percentage
ANGPTL3, H1:60%
Angiopoietin-like 3 protein
Mean reticulocyte volume
ANGPTL3, H3:100%
Angiopoietin-like 3 protein
Mean sphered cell volume
ANGPTL3, H1:97%
Angiopoietin-like 3 protein
Immature reticulocyte fraction
ANGPTL3, H3:100%
Angiopoietin-like 3 protein
High light scatter reticulocyte percentage
ANGPTL3, H1:95%
Angiopoietin-like 3 protein
Microalbumin in urine
ANGPTL3, H1:86%
Angiopoietin-like 3 protein
Creatinine (enzymatic) in urine
-log10(P-value)
62,347,750
62,854,650
Position (chr 1)

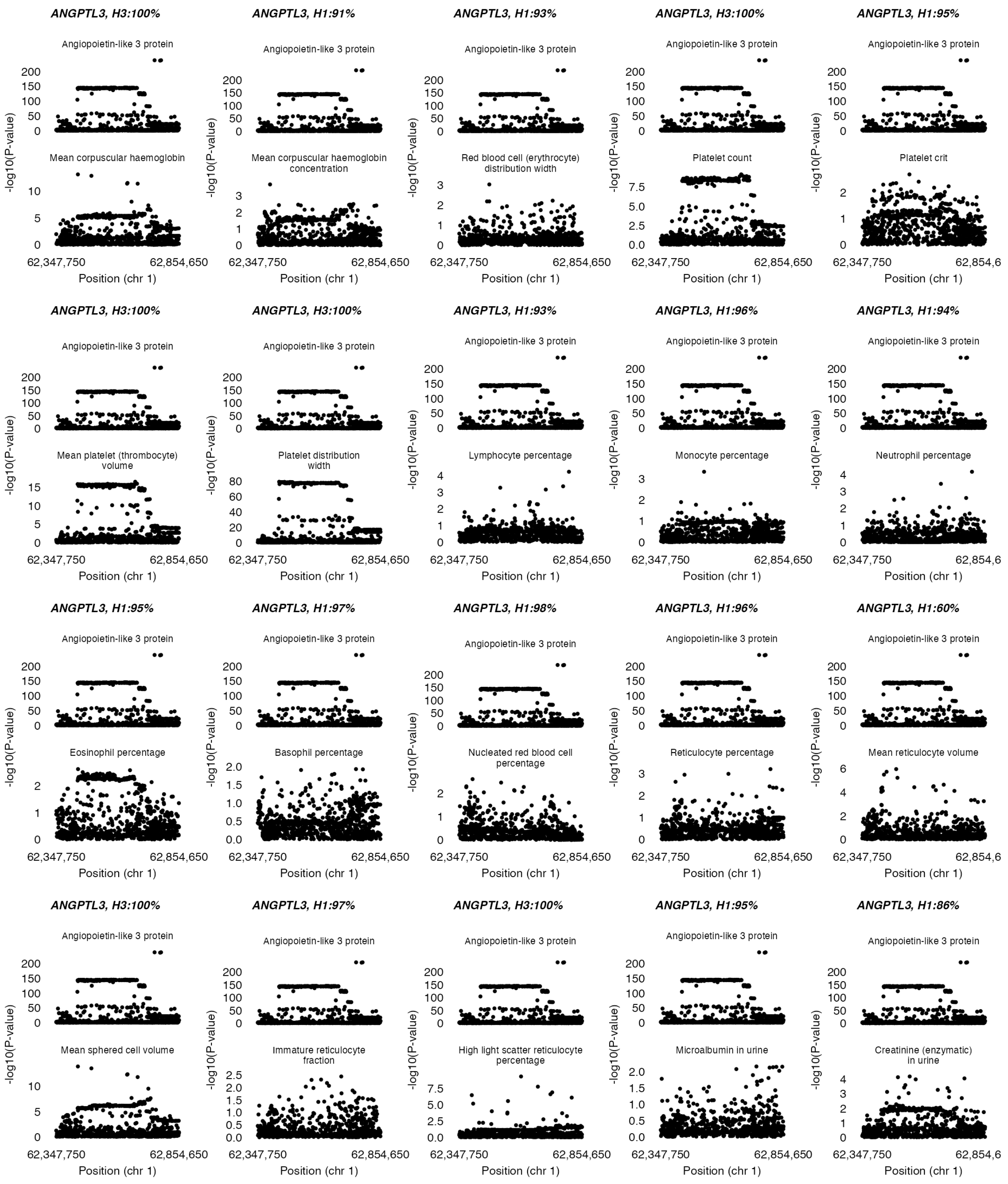
ANGPTL3, H3:100%
Angiopoietin-like 3 protein
Mean corpuscular haemoglobin
ANGPTL3, H1:91%
Angiopoietin-like 3 protein
Mean corpuscular haemoglobin concentration
ANGPTL3, H1:93%
Angiopoietin-like 3 protein
Red blood cell (erythrocyte) distribution width
ANGPTL3, H3:100%
Angiopoietin-like 3 protein
Platelet count
ANGPTL3, H1:95%
Angiopoietin-like 3 protein
Platelet crit
ANGPTL3, H3:100%
Angiopoietin-like 3 protein
Mean platelet (thrombocyte) volume
ANGPTL3, H3:100%
Angiopoietin-like 3 protein
Platelet distribution width
ANGPTL3, H1:93%
Angiopoietin-like 3 protein
Lymphocyte percentage
ANGPTL3, H1:96%
Angiopoietin-like 3 protein
Monocyte percentage
ANGPTL3, H1:94%
Angiopoietin-like 3 protein
Neutrophil percentage
ANGPTL3, H1:95%
Angiopoietin-like 3 protein
Eosinophil percentage
ANGPTL3, H1:97%
Angiopoietin-like 3 protein
Basophil percentage
ANGPTL3, H1:98%
Angiopoietin-like 3 protein
Nucleated red blood cell percentage
ANGPTL3, H1:96%
Angiopoietin-like 3 protein
Reticulocyte percentage
ANGPTL3, H1:60%
Angiopoietin-like 3 protein
Mean reticulocyte volume
ANGPTL3, H3:100%
Angiopoietin-like 3 protein
Mean sphered cell volume
ANGPTL3, H1:97%
Angiopoietin-like 3 protein
Immature reticulocyte fraction
ANGPTL3, H3:100%
Angiopoietin-like 3 protein
High light scatter reticulocyte percentage
ANGPTL3, H1:95%
Angiopoietin-like 3 protein
Microalbumin in urine
ANGPTL3, H1:86%
Angiopoietin-like 3 protein
Creatinine (enzymatic) in urine
-log10(P-value)
62,347,750
62,854,650
Position (chr 1)

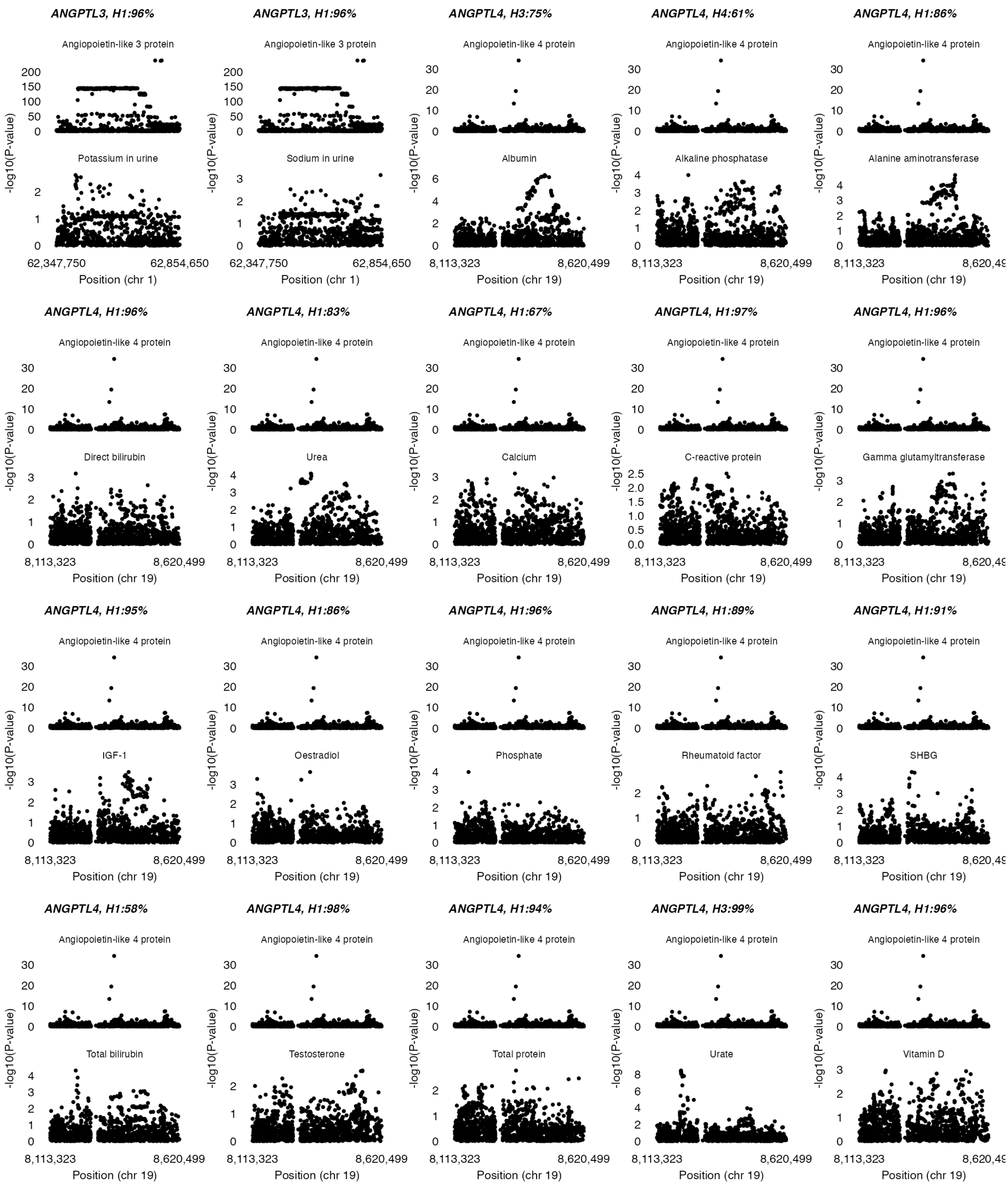
ANGPTL3, H1:96%
Angiopoietin-like 3 protein
Potassium in urine
ANGPTL3, H1:96%
Angiopoietin-like 3 protein
Sodium in urine
62,347,750
62,854,650
Position (chr 1)
ANGPTL4, H3:75%
Angiopoietin-like 4 protein
Albumin
ANGPTL4, H4:61%
Angiopoietin-like 4 protein
Alkaline phosphatase
ANGPTL4, H1:86%
Angiopoietin-like 4 protein
Alanine aminotransferase
ANGPTL4, H1:96%
Angiopoietin-like 4 protein
Direct bilirubin
ANGPTL4, H1:83%
Angiopoietin-like 4 protein
Urea
ANGPTL4, H1:67%
Angiopoietin-like 4 protein
Calcium
ANGPTL4, H1:97%
Angiopoietin-like 4 protein
C-reactive protein
ANGPTL4, H1:96%
Angiopoietin-like 4 protein
Gamma glutamyltransferase
ANGPTL4, H1:95%
Angiopoietin-like 4 protein
IGF-1
ANGPTL4, H1:86%
Angiopoietin-like 4 protein
Oestradiol
ANGPTL4, H1:96%
Angiopoietin-like 4 protein
Phosphate
ANGPTL4, H1:89%
Angiopoietin-like 4 protein
Rheumatoid factor
ANGPTL4, H1:91%
Angiopoietin-like 4 protein
SHBG
ANGPTL4, H1:58%
Angiopoietin-like 4 protein
Total bilirubin
ANGPTL4, H1:98%
Angiopoietin-like 4 protein
Testosterone
ANGPTL4, H1:94%
Angiopoietin-like 4 protein
Total protein
ANGPTL4, H3:99%
Angiopoietin-like 4 protein
Urate
ANGPTL4, H1:96%
Angiopoietin-like 4 protein
Vitamin D
-log10(P-value)
8,113,323
8,620,499
Position (chr 19)

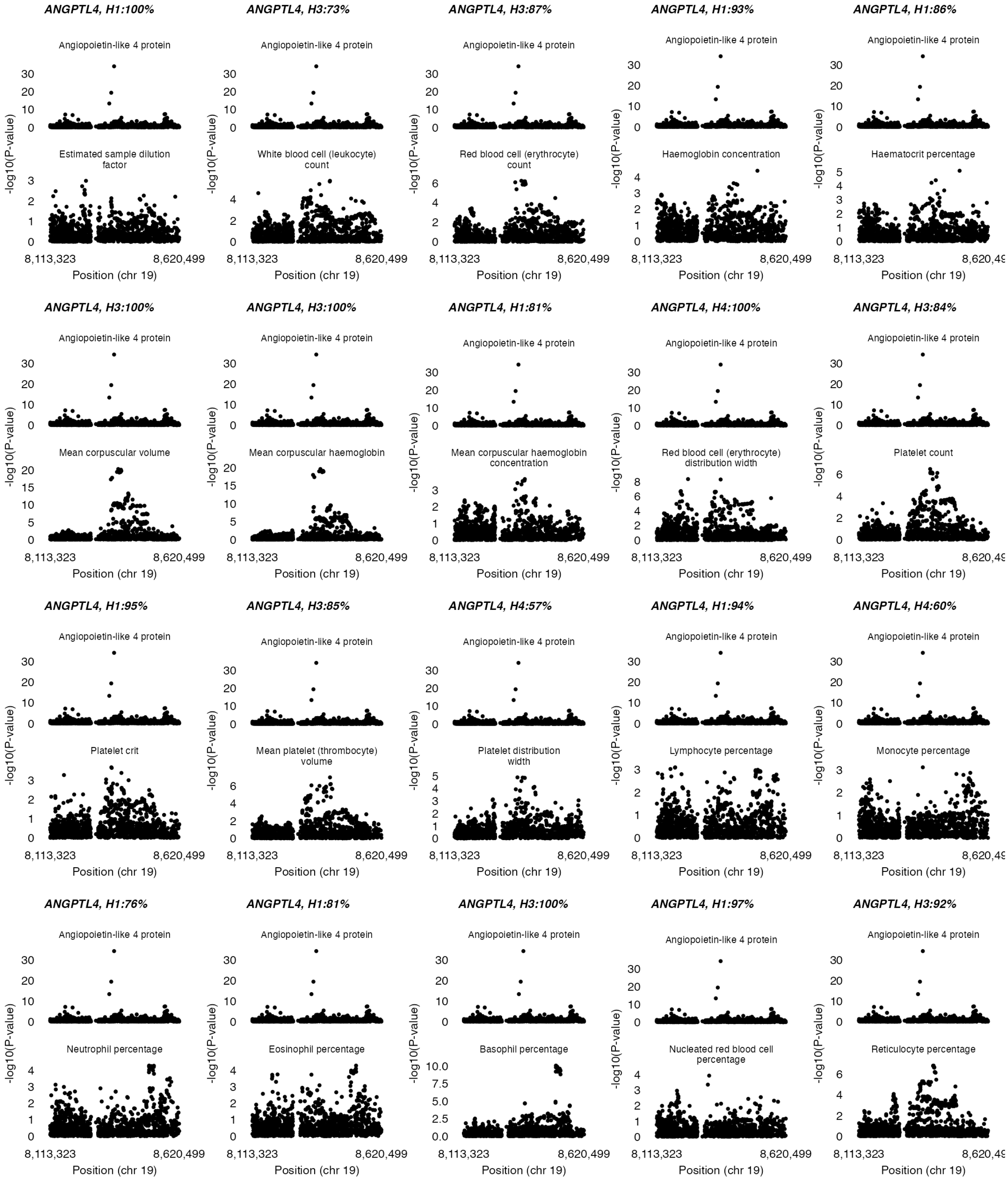
ANGPTL4, H1:100%
Angiopoietin-like 4 protein
Estimated sample dilution factor
ANGPTL4, H3:73%
Angiopoietin-like 4 protein
White blood cell (leukocyte) count
ANGPTL4, H3:87%
Angiopoietin-like 4 protein
Red blood cell (erythrocyte) count
ANGPTL4, H1:93%
Angiopoietin-like 4 protein
Haemoglobin concentration
ANGPTL4, H1:86%
Angiopoietin-like 4 protein
Haematocrit percentage
ANGPTL4, H3:100%
Angiopoietin-like 4 protein
Mean corpuscular volume
ANGPTL4, H3:100%
Angiopoietin-like 4 protein
Mean corpuscular haemoglobin
ANGPTL4, H1:81%
Angiopoietin-like 4 protein
Mean corpuscular haemoglobin concentration
ANGPTL4, H4:100%
Angiopoietin-like 4 protein
Red blood cell (erythrocyte) distribution width
ANGPTL4, H3:84%
Angiopoietin-like 4 protein
Platelet count
ANGPTL4, H1:95%
Angiopoietin-like 4 protein
Platelet crit
ANGPTL4, H3:85%
Angiopoietin-like 4 protein
Mean platelet (thrombocyte) volume
ANGPTL4, H4:57%
Angiopoietin-like 4 protein
Platelet distribution width
ANGPTL4, H1:94%
Angiopoietin-like 4 protein
Lymphocyte percentage
ANGPTL4, H4:60%
Angiopoietin-like 4 protein
Monocyte percentage
ANGPTL4, H1:76%
Angiopoietin-like 4 protein
Neutrophil percentage
ANGPTL4, H1:81%
Angiopoietin-like 4 protein
Eosinophil percentage
ANGPTL4, H3:100%
Angiopoietin-like 4 protein
Basophil percentage
ANGPTL4, H1:97%
Angiopoietin-like 4 protein
Nucleated red blood cell percentage
ANGPTL4, H3:92%
Angiopoietin-like 4 protein
Reticulocyte percentage
-log10(P-value)
8,113,323
8,620,499
8,620,4
Position (chr 19)

25

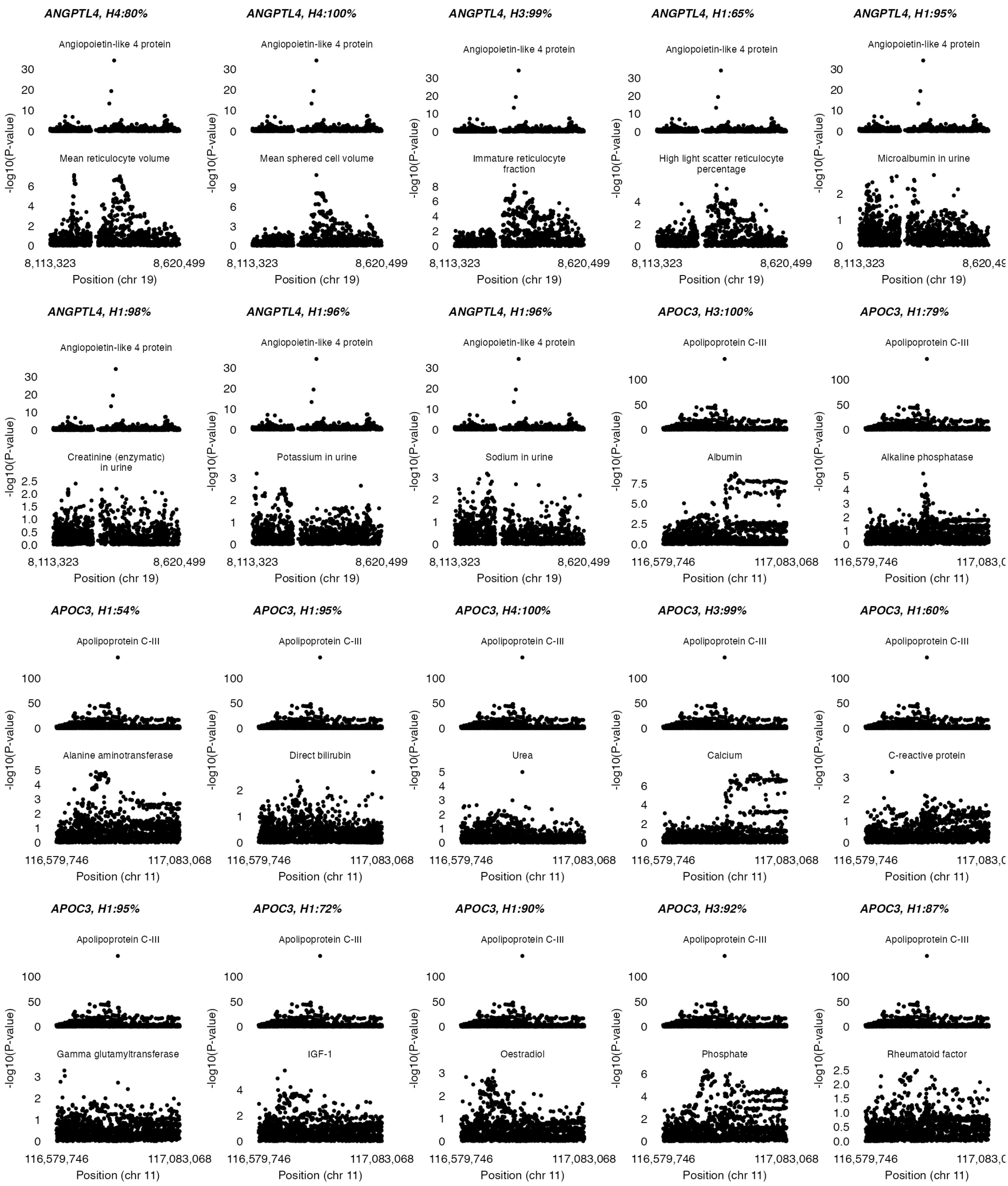
ANGPTL4, H4:80%
Angiopoietin-like 4 protein
Mean reticulocyte volume
ANGPTL4, H4:100%
Angiopoietin-like 4 protein
Mean sphered cell volume
ANGPTL4, H3:99%
Angiopoietin-like 4 protein
Immature reticulocyte fraction
ANGPTL4, H1:65%
Angiopoietin-like 4 protein
High light scatter reticulocyte percentage
ANGPTL4, H1:95%
Angiopoietin-like 4 protein
Microalbumin in urine
ANGPTL4, H1:98%
Angiopoietin-like 4 protein
Creatinine (enzymatic) in urine
ANGPTL4, H1:96%
Angiopoietin-like 4 protein
Potassium in urine
ANGPTL4, H1:96%
Angiopoietin-like 4 protein
Sodium in urine
APOC3, H3:100%
Apolipoprotein C-III
Albumin
APOC3, H1:79%
Apolipoprotein C-III
Alkaline phosphatase
APOC3, H1:54%
Apolipoprotein C-III
Alanine aminotransferase
APOC3, H1:95%
Apolipoprotein C-III
Direct bilirubin
APOC3, H4:100%
Apolipoprotein C-III
Urea
APOC3, H3:99%
Apolipoprotein C-III
Calcium
APOC3, H1:60%
Apolipoprotein C-III
C-reactive protein
APOC3, H1:95%
Apolipoprotein C-III
Gamma glutamyltransferase
APOC3, H1:72%
Apolipoprotein C-III
IGF-1
APOC3, H1:90%
Apolipoprotein C-III
Oestradiol
APOC3, H3:92%
Apolipoprotein C-III
Phosphate
APOC3, H1:87%
Apolipoprotein C-III
Rheumatoid factor
-log10(P-value)
8,113,323
8,620,499
Position (chr 19)
116,579,746
117,083,068
Position (chr 11)

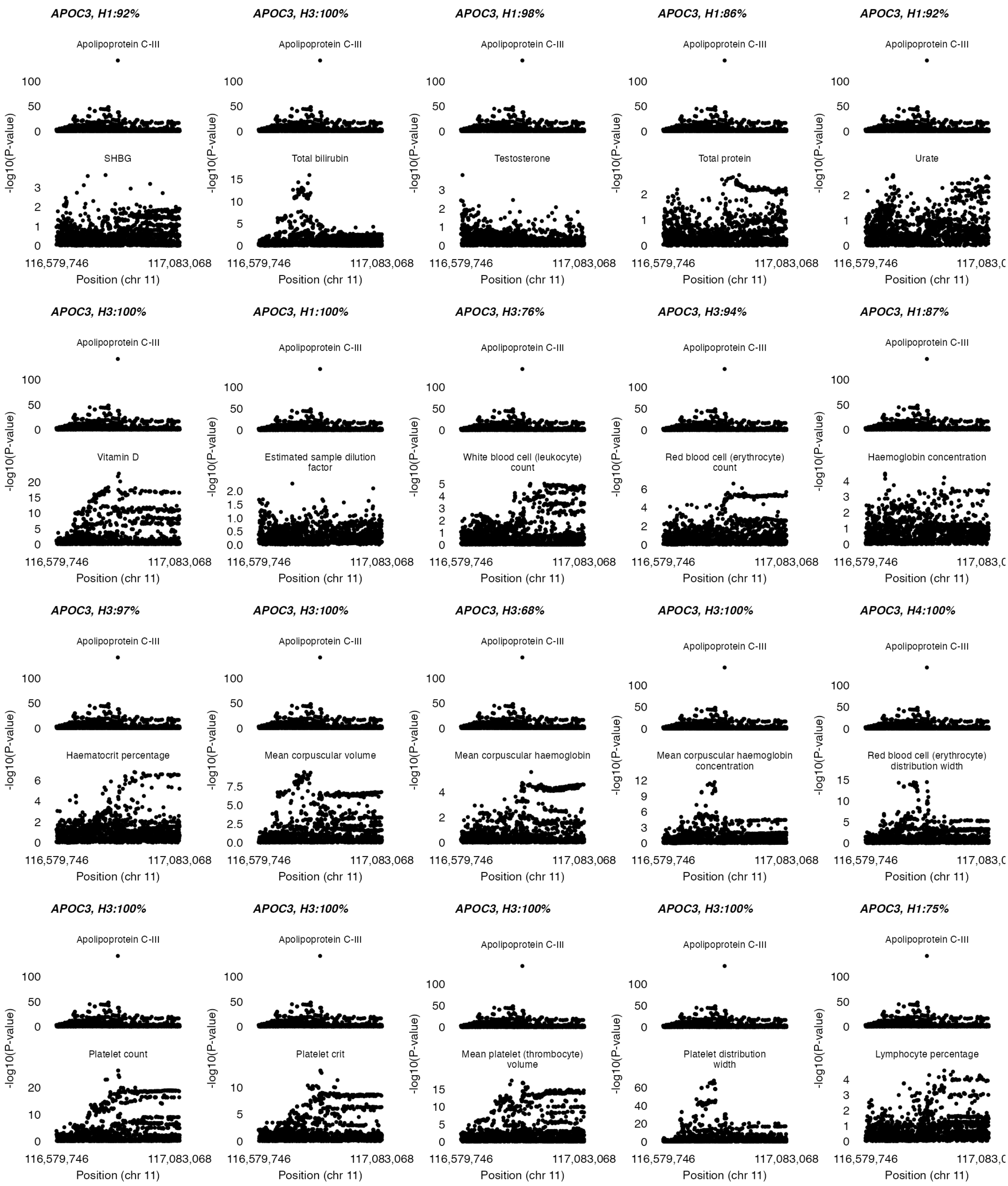
APOC3, H1:92%
Apolipoprotein C-III
SHBG
APOC3, H3:100%
Apolipoprotein C-III
Total bilirubin
APOC3, H1:98%
Apolipoprotein C-III
Testosterone
APOC3, H1:86%
Apolipoprotein C-III
Total protein
APOC3, H1:92%
Apolipoprotein C-III
Urate
APOC3, H3:100%
Apolipoprotein C-III
Vitamin D
APOC3, H1:100%
Apolipoprotein C-III
Estimated sample dilution factor
APOC3, H3:76%
Apolipoprotein C-III
White blood cell (leukocyte) count
APOC3, H3:94%
Apolipoprotein C-III
Red blood cell (erythrocyte) count
APOC3, H1:87%
Apolipoprotein C-III
Haemoglobin concentration
APOC3, H3:97%
Apolipoprotein C-III
Haematocrit percentage
APOC3, H3:100%
Apolipoprotein C-III
Mean corpuscular volume
APOC3, H3:68%
Apolipoprotein C-III
Mean corpuscular haemoglobin
APOC3, H3:100%
Apolipoprotein C-III
Mean corpuscular haemoglobin concentration
APOC3, H4:100%
Apolipoprotein C-III
Red blood cell (erythrocyte) distribution width
APOC3, H3:100%
Apolipoprotein C-III
Platelet count
APOC3, H3:100%
Apolipoprotein C-III
Platelet crit
APOC3, H3:100%
Apolipoprotein C-III
Mean platelet (thrombocyte) volume
APOC3, H3:100%
Apolipoprotein C-III
Platelet distribution width
APOC3, H1:75%
Apolipoprotein C-III
Lymphocyte percentage
-log10(P-value)
116,579,746
117,083,068
Position (chr 11)

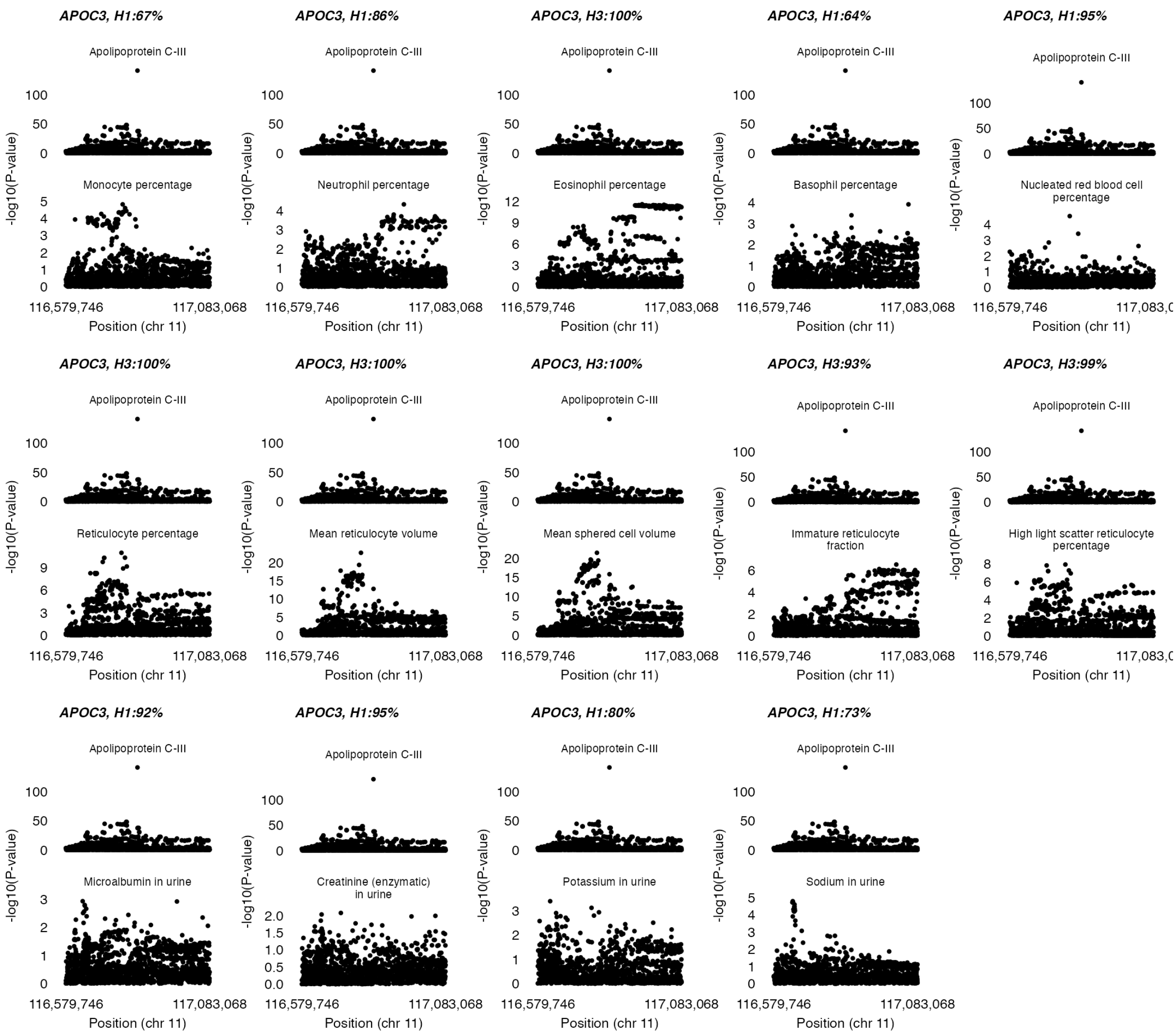
APOC3, H1:67%
Apolipoprotein C-III
Monocyte percentage
APOC3, H1:86%
Apolipoprotein C-III
Neutrophil percentage
APOC3, H3:100%
Apolipoprotein C-III
Eosinophil percentage
APOC3, H1:64%
Apolipoprotein C-III
Basophil percentage
APOC3, H1:95%
Apolipoprotein C-III
Nucleated red blood cell percentage
APOC3, H3:100%
Apolipoprotein C-III
Reticulocyte percentage
APOC3, H3:100%
Apolipoprotein C-III
Mean reticulocyte volume
APOC3, H3:100%
Apolipoprotein C-III
Mean sphered cell volume
APOC3, H3:93%
Apolipoprotein C-III
Immature reticulocyte fraction
APOC3, H3:99%
Apolipoprotein C-III
High light scatter reticulocyte percentage
APOC3, H1:92%
Apolipoprotein C-III
Microalbumin in urine
APOC3, H1:95%
Apolipoprotein C-III
Creatinine (enzymatic) in urine
APOC3, H1:80%
Apolipoprotein C-III
Potassium in urine
APOC3, H1:73%
Apolipoprotein C-III
Sodium in urine
-log10(P-value)
116,579,746
117,083,068
Position (chr 11)

### SUPPLEMENTAL FIGURE 3

**Figure S3. Results of cis-pQTL MR analyses of the estimated glomerular filtration rate by (eGFR) by Cystatin C and plasma Creatinine.** The findings are displayed in bar graphs, illustrating the level of the effect per s.d. decrease in protein abundance. The red lines represent the 95% CI.

**Cardiometabolic risk factors**

**Exposure: Target protein abundance**

| Outcome | Sample size | Gene | No. SNPs | Effect size (95% CI) | P-value | Coloc. |
|---|---|---|---|---|---|---|
| Est. glomerular filtration rate (cystatin c) | 1,201,909 | *ANGPTL3* | 3 | + 0.00 (0.00;0.01) | 1.000 | H1: 100% |
| | | *ANGPTL4* | 1 | 0.00 (−0.01;0.02) | 1.000 | H1: 100% |
| | | *APOC3* | 1 | 0.00 (−0.02;0.01) | 1.000 | H1: 100% |
| | | *APOC3 c.55+1G>A* | 0 | | | |
| Est. glomerular filtration rate (creatinine) | 1,004,040 | *ANGPTL3* | 3 | + 0.00 (0.00;0.01) | 0.148 | H1: 100% |
| | | *ANGPTL4* | 1 | + 0.01 (0.00;0.02) | 1.000 | H1: 100% |
| | | *APOC3* | 1 | 0.00 (−0.01;0.01) | 1.000 | H1: 100% |
| | | *APOC3 c.55+1G>A* | 0 | | | |

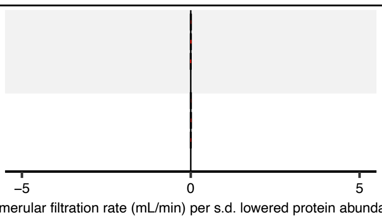

### SUPPLEMENTAL FIGURE 4

**Figure S4. Results of phenome-wide MR analysis on FinnGen outcomes using *ANGPTL3 rs34483103-1:62604866:AGTTAATGTG>A* [3 prime UTR, c.*52_*60del] and *APOC3 rs138326449-11:116830638:G>A* [donor loss, c.55+1G>A] variants. A**: Volcano plot displaying the results of ANGPTL3 cis-pQTL phenome-wide MR. **B**: APOC3 cis-pQTL phenome-wide MR volcano plot. The y-axis solid straight lines indicate the phenome-wide significance threshold. 'OR' indicates the odds ratio with 95% confidence intervals with Bonferroni correction for the 694 FinnGen outcomes.

**A. *ANGPTL3 rs34483103*-1:62604866:AGTTAATGTG>A [3 prime UTR, c.*52_*60del]**

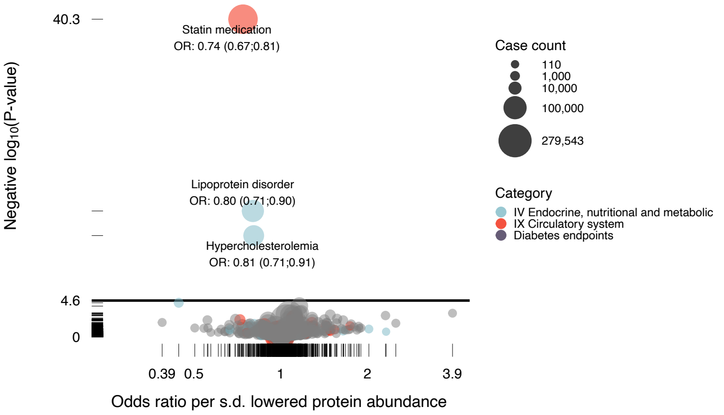

**B. *APOC3 rs138326449*-11:116830638:G>A [donor loss, c.55+1G>A]**

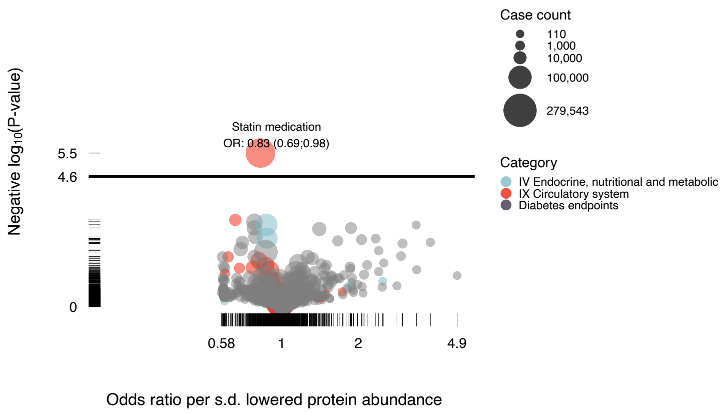

### SUPPLEMENTAL FIGURE 5

**Figure S5. Cis-pQTL MR of the clinical laboratory outcomes, showing the results on a 1-s.d. scale.** The red bars indicate the 95 % CI. The black dots indicate the effect point estimate. '*' indicate P < 0.05. '**' indicates P < 0.05 with a shared causal variant ($H_4$). A list explaining the abbreviations is provided in the supplemental material (**Supplemental Table 1**).

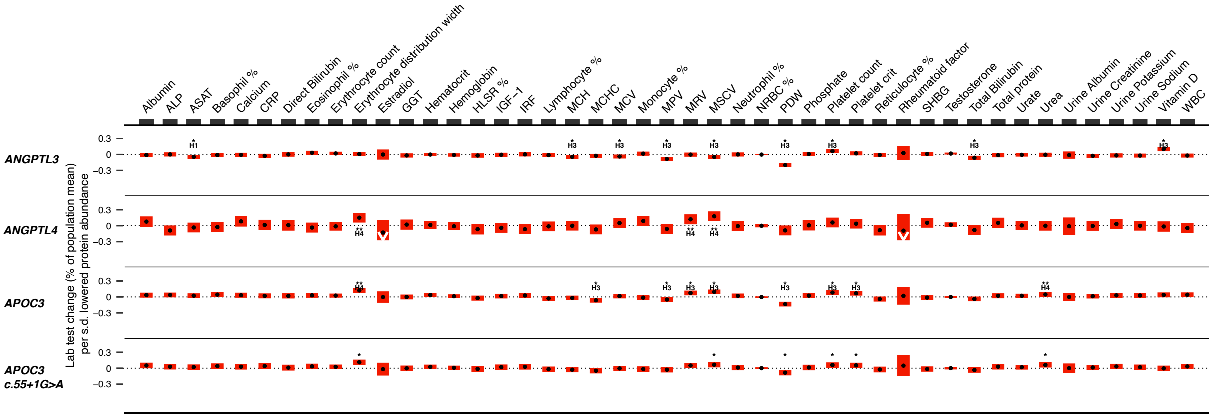

### SUPPLEMENTAL FIGURE 6

**Figure S6. Wildtype human ANGPTL4 protein and the ANGPTL4 E40K variant protein are detected to a similar extent by the antibody used in the human ANGPTL4 ELISA.** Western blot of the precipitated medium of HEK293 cells transfected with expression vectors for different mutant forms of human ANGPTL4 fused to a V5-tag (34). CC/AA is an oligomerization defective ANGPTL4 variant. SDS-PAGE was performed using a loading buffer without DTT or other disulfide-reducing agent. Equal amounts of medium were loaded. Membranes were blotted with an antibody from R&D Systems that is used for the human ANGPTL4 Elisa (AF3485, 1:2500). Secondary antibody was goat anti-rabbit IgG conjugated to HRP (1:5000).

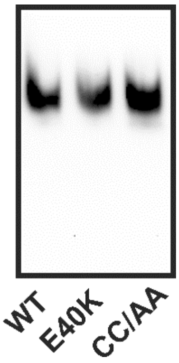
